# Optimising social mixing strategies achieving COVID-19 herd immunity while minimising mortality in six European countries

**DOI:** 10.1101/2020.08.25.20182162

**Authors:** Romain Ragonnet, Guillaume Briffoteaux, Bridget M. Williams, Julian Savulescu, Matthew Segal, Milinda Abayawardana, Rosalind M. Eggo, Daniel Tuyttens, Nouredine Melab, Ben J. Marais, Emma S. McBryde, James M. Trauer

**Author notes:** Corresponding author: Dr Romain Ragonnet, Monash School of Public Health and Preventive Medicine, 553 St Kilda Road Melbourne VIC 3004.

## Abstract

Strategies are needed to minimise the impact of COVID-19 in the medium-to-long term, until safe and effective vaccines can be used. Using a mathematical model in a formal optimisation framework, we identified contact mitigation strategies that minimised COVID-19-related mortality over a time-horizon of 15 months while achieving herd immunity in six or 12 months, in Belgium, France, Italy, Spain, Sweden, and the UK. We show that manipulation of social contacts by age can reduce the impact of COVID-19 considerably in the presence of intense transmission. If immunity was persistent, the optimised scenarios would result in herd immunity while causing a number of deaths considerably lower than that observed during the March-April European wave in Belgium, France, Spain and Sweden, whereas the numbers of deaths required to achieve herd immunity would be comparable to somewhat larger that the past epidemics in Italy and the UK. Our results also suggest that countries’ herd immunity thresholds may be considerably lower than first estimated for SARS-CoV-2. If post-infection immunity was short-lived, ongoing contact mitigation would be required to prevent major epidemic resurgence.

## Introduction

The rapid spread of SARS-CoV-2 has resulted in millions of cases of COVID-19 and hundreds of thousands of deaths. This has resulted in a global crisis that has overwhelmed health care systems and induced economic hardship in many settings.

In an effort to control the epidemic, many governments have implemented severe restrictions on population movement and social mixing. These have varied in scope and stringency^1^, and have included ‘stay at home’ orders, travel restrictions, and school and business closures. Although these measures, combined with extensive testing, quarantine, and contact tracing with isolation, have been successful in reducing transmission in many countries, the adverse community-wide effects of these restrictions have been severe. Evidence from the United Kingdom (UK) suggests restrictions have had negative effects on mental health^2^, and non-COVID-19 health through delays in diagnostic services^3^.

In the absence of a vaccine, there are few alternative approaches to combating the pandemic; each associated with major drawbacks which should be objectively quantified. Although elimination of infection has been successful in some settings, attempts to ease restrictions have often resulted in epidemic recurrence. As long as a large proportion of the population remains susceptible to SARS-CoV-2 infection, populations will remain at high risk of resurgences of transmission. Reaching a level of post-infection immunity in the population that results in the effective reproduction number remaining below 1 (also called “herd immunity”) can be part of a strategy to minimise population health impacts over the medium-to-long term^4^.

As countries may not be able to sustain strict movement and contact restrictions in the medium-to-long term, it is important to identify which restrictions can be lifted with lesser impacts than a relaxation of measures across all ages and locations.

Countries such as Sweden have tried to minimise the impact of the disease with less restrictive measures, to slow transmission while shielding those at greatest risk^5^. However, Swedish authorities have acknowledged errors in implementation, particularly around infection prevention in aged residential care facilities^6^. The UK also initially aimed for a limited lockdown with shielding of at-risk groups, but changed course after modelling suggested that without drastic measures, hundreds of thousands of deaths would be expected^7^.

Permitting SARS-CoV-2 transmission in the community is clearly associated with negative consequences, including the potential long-term effects of COVID-19 that are still poorly understood and the potential overwhelming of health systems. However, until safe and effective vaccines can be deployed at scale, all strategies are associated with serious risks and adverse effects. It is therefore necessary to evaluate the merits and risks of each approach.

We present an optimisation analysis that aims to identify strategies of social contact restrictions resulting in non-vaccine herd immunity within six or 12 months while minimising the number of COVID-19-related deaths or years of life lost (YLLs) over a time-horizon of 15 months in six highly-affected countries: Belgium, France, Italy, Spain, Sweden, and the UK. We consider strategies that either altered age- or location-specific contact patterns.

## Results

### Model calibration

Model fits to local data on COVID-19 cases, hospitalisations, deaths and seroprevalence are shown in Figure 1. Our model was able to replicate the observed dynamics of the different disease indicators in the six countries. Our approach of allowing for time-variant case detection successfully captured the differences observed between the profiles of confirmed cases and those of hospitalisations or deaths. For example, in Sweden the model was able to replicate the increase observed in the number of confirmed cases in June 2020 while other disease indicators were declining. This is explained by a testing surge that occurred in June in Sweden^8^, and that was automatically captured by our Bayesian model calibration (Figure S8).

**Figure 1.**
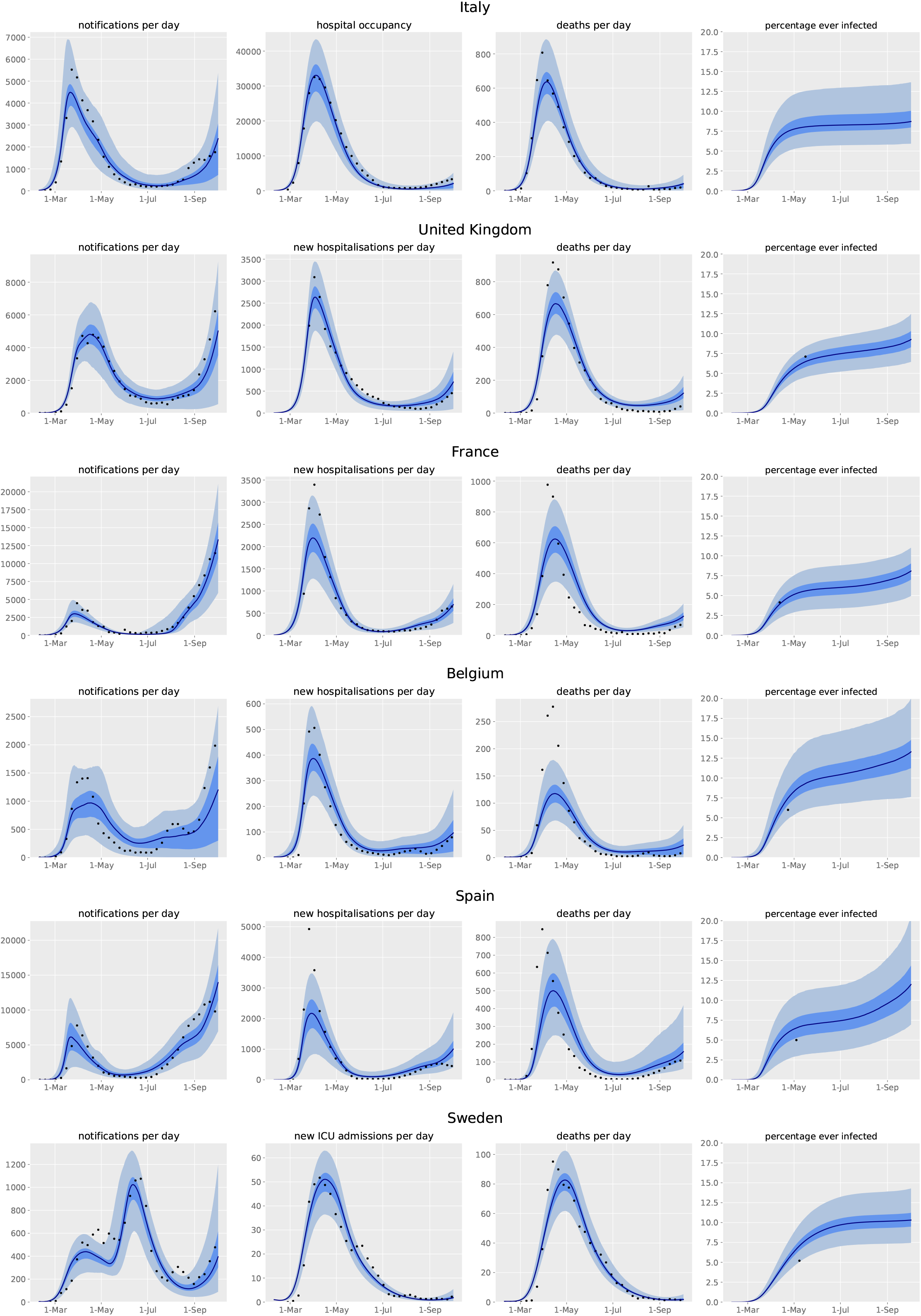
Model projections compared against local data. The figures present the median estimates (dark blue line) and the central 95% credible intervals (light blue shade) against observed numbers of confirmed COVID-19 cases, hospitalisations, deaths and seroprevalence (black dots). The x-axis represents the dates of year 2020. The data points represent the weekly average of the daily counts for cases, hospitalisations and deaths.

The posterior parameter estimates and inferred time-variant profiles of case detection obtained during the model calibration process are presented in Supplementary Section 2.4. While the posterior distributions of most epidemiological parameters were broadly similar across the six countries, we observed significant differences regarding the estimates of mortality rates, detection profiles and the inferred effect of micro-distancing (i.e. the reduction of the per-contact risk of transmission attributable to preventive measures such as mask wearing or physical distancing). The modelled infection fatality rates were found to be higher in the UK and Belgium compared to the other countries. This finding has also been reported in previous studies and may be explained by differences in the criteria used to determine whether deaths are COVID-19-related^9,10^. We also found that higher levels of case detection had to be modelled in France and Sweden compared to the other countries in order to capture the different disease indicators accurately. We noted that the dynamics of the Swedish epidemic were best captured when micro-distancing was applied sooner and at a higher level compared to the other countries. Finally, small variations were also observed in the risk of transmission per contact between countries, which can be explained by the fact that the contact matrices used to inform the model only capture the average number of contacts per day but fail to account for other contact characteristics such as contact duration or intensity.

In order to validate the age-specific dynamics of the models, we compared the modelled age-specific proportions of recovered individuals with estimates of seroprevalence by age obtained from 24 serosurveys (Supplementary Section 5.1), covering all countries except Italy for which age-specific estimates were not available. These comparisons demonstrated that the model estimates were consistent with serosurvey measures, especially when surveys were conducted shortly after the first epidemic waves. In contrast, the proportions of recovered individuals predicted by the model tended to overestimate observed seroprevalence for the surveys that were conducted at later times, which is consistent with the known decline in antibody prevalence over time since infection^10^.

### Optimisation results

In all countries, age-specific mixing restrictions resulted in fewer deaths and YLLs than location-specific mixing reductions (Table 1), although both generated considerably fewer deaths and YLLs compared to an unmitigated scenario. The optimisations also led to reductions of up to 50% in the final proportion of ever-infected individuals, compared to the unmitigated scenario (Table 2). The total number of deaths occurring after the start of the optimised intervention on 1^st^ October 2020 was significantly lower than the number of deaths that had occurred before this time in Belgium, France, Spain and Sweden when considering optimisation by age. In contrast, achieving herd immunity while optimising mixing by location would lead to a greater number of deaths compared to those that had occurred before the 1^st^ October 2020 in all countries.

**Table 1.**
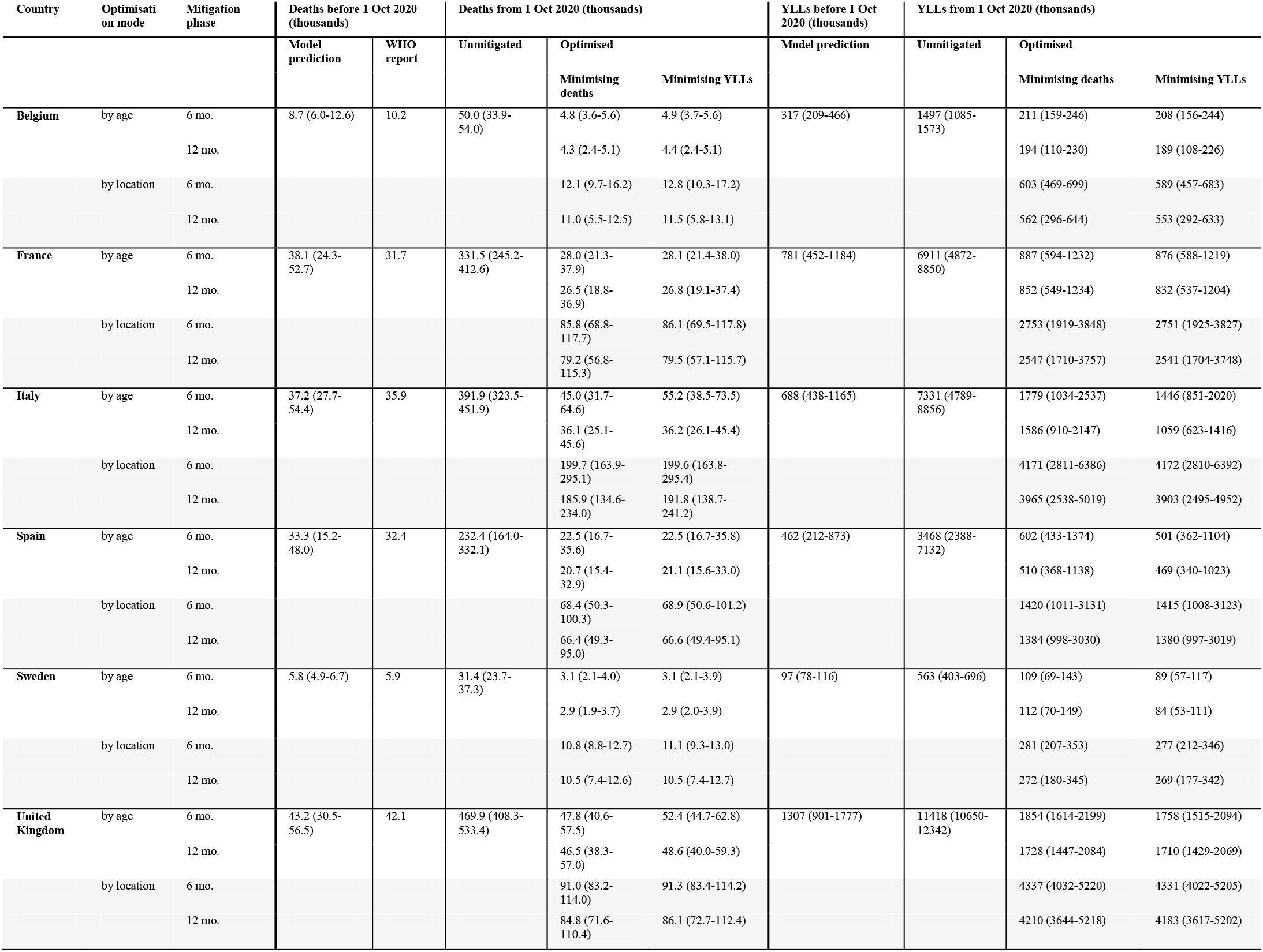
Predicted numbers of deaths and years of life lost. Optimisation realised under the assumption of persistent immunity. Numbers are presented in thousands of deaths and thousands of YLLs as median and central 95% credible intervals. YLLs: Years of life lost.

**Table 2.** Proportions of recovered individuals at the start of the mitigation phase and at the end of the simulation. Optimisations realised under the assumption of persistent immunity. Numbers are presented as median and central 95% credible intervals. Herd immunity was reached by the end of the mitigation phase. YLLs: Years of life lost.

| Country | Optimisation mode | Mitigation phase | Proportion recovered on 1 Oct 2020 (%) | Final proportion recovered (%) |  |  |
| --- | --- | --- | --- | --- | --- | --- |
|  |  |  |  | Unmitigated | Optimised Minimising deaths | Optimised Minimising YLLs |
| Belgium | by age | 6 months | 13 (9-20) | 64 (58-68) | 36 (32-40) | 36 (31-40) |
|  |  | 12 months |  |  | 35 (29-39) | 34 (29-39) |
|  | by location | 6 months |  |  | 38 (35-44) | 39 (35-44) |
|  |  | 12 months |  |  | 36 (29-42) | 37 (29-42) |
| France | by age | 6 months | 7 (5-11) | 65 (62-71) | 31 (28-35) | 32 (29-36) |
|  |  | 12 months |  |  | 30 (27-35) | 30 (27-34) |
|  | by location | 6 months |  |  | 37 (35-43) | 37 (35-43) |
|  |  | 12 months |  |  | 35 (31-43) | 35 (31-42) |
| Italy | by age | 6 months | 7 (5-10) | 73 (68-78) | 43 (40-47) | 42 (39-46) |
|  |  | 12 months |  |  | 39 (36-43) | 39 (36-43) |
|  | by location | 6 months |  |  | 50 (47-60) | 50 (47-60) |
|  |  | 12 months |  |  | 46 (39-53) | 46 (39-53) |
| Spain | by age | 6 months | 9 (5-13) | 79 (76-82) | 41 (37-45) | 41 (37-45) |
|  |  | 12 months |  |  | 39 (34-42) | 39 (35-43) |
|  | by location | 6 months |  |  | 46 (42-51) | 46 (42-52) |
|  |  | 12 months |  |  | 46 (40-51) | 46 (39-51) |
| Sweden | by age | 6 months | 11 (9-14) | 82 (79-83) | 46 (44-47) | 46 (44-47) |
|  |  | 12 months |  |  | 43 (41-44) | 45 (43-46) |
|  | by location | 6 months |  |  | 52 (51-54) | 52 (51-55) |
|  |  | 12 months |  |  | 51 (47-53) | 51 (47-53) |
| UK | by age | 6 months | 9 (6-12) | 75 (72-80) | 41 (37-46) | 41 (37-45) |
|  |  | 12 months |  |  | 40 (35-44) | 39 (35-44) |
|  | by location | 6 months |  |  | 43 (40-50) | 43 (40-50) |
|  |  | 12 months |  |  | 42 (37-49) | 42 (37-49) |

We observed broadly consistent mixing patterns across all the countries considered under the optimised mitigation strategies. Namely, contacts of older adults were restricted most, while contacts of individuals aged between 15 and 49 years old were maintained near pre-COVID-19 levels for optimisation of deaths or YLLs (Figure 2). Contacts involving children and adolescents were also maintained at or near 100% in most countries under the optimised scenarios. The mortality indicators (deaths and YLLs) were highly sensitive to small perturbations in the mixing contributions of young-to-middle-age adults considered in the sensitivity analyses. In contrast, the contact rates involving children and adolescents could deviate significantly from the optimal plan without substantially compromising outcomes.

**Figure 2.**
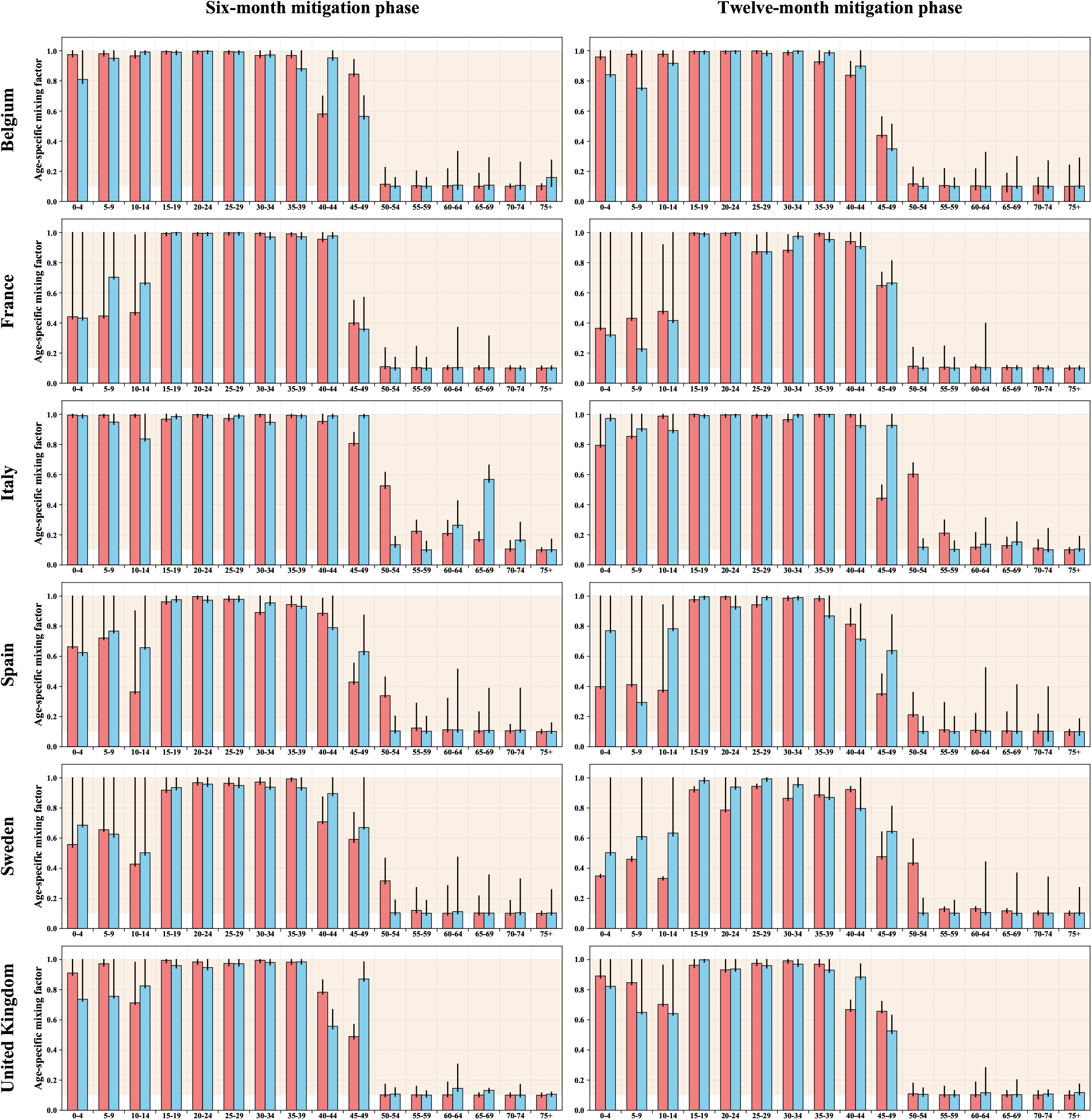
Optimal mixing pattern with contact mitigation by age. The red bars and the blue bars represent the optimised age-specific mixing factors when minimising the number of deaths and years of life lost, respectively. The mixing factors are presented as relative values compared to the pre-COVID-19 era for each age group. A value of 1 means that individuals have the same opportunity of effective contact as before the pandemic, whereas a value of 0.1 indicates a 90% reduction in effective contact opportunity. To determine the relative effective contact rate of one age category with another, the relative mixing values of each of the two categories must be multiplied together to reflect both the contactor’s and the contactee’s reduction in contact opportunity. The coloured background represents the acceptable region for the mixing factors (i.e. the interval [0.1 - 1]). The thin black bars represent the maximum change in individual age-group contributions that would cause an excess of no more than 20 deaths per million people (red bars) or 1000 YLLs per million people (blue bars) as compared to the optimal plan, while still reaching herd immunity by the end of the mitigation phase. The left and right panels show the result obtained when assuming that the mitigation phase lasts 6 and 12 months, respectively. The optimisations were performed based on the countries’ maximum a posteriori parameter sets.

Optimising for YLLs, rather than deaths, resulted in a larger decrease in contacts needed in the 50-54-year-old age-group across most scenarios. These reductions were compensated by contact increases in younger age groups, as these groups are associated with a lower mortality risk. The patterns of optimal mixing by age were similar between the six-month and 12-month mitigation strategies, although the longer scenario necessitated slightly greater contact reductions.

While the base-case analyses assumed a minimum threshold of 10% for the mixing factors, we considered alternate lower bounds in a sensitivity analysis (Supplement, Section 5.1). We observed that the two mortality indicators increased roughly linearly, as the minimum mixing threshold was raised (Figure S15). Considering a threshold of 20%, we predicted that the number of deaths required to reach herd immunity would increase by 30 to 65% compared to the base-case analyses assuming a 10% minimum threshold.

Figure 3 shows the optimised mixing patterns by location..

**Figure 3.**
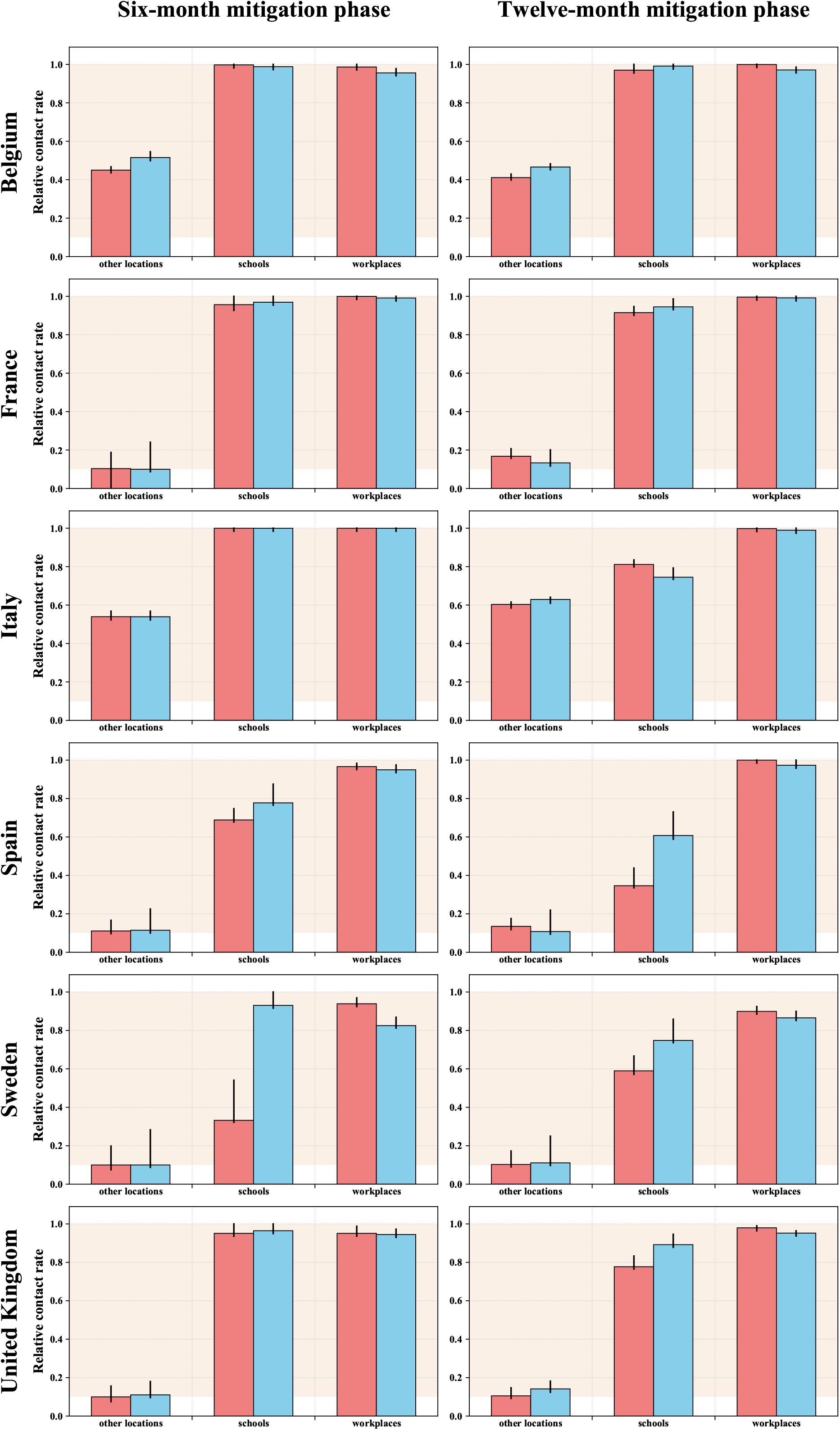
Optimal mixing pattern with contact mitigation by location. Red and blue bars represent the optimised relative contact rates by location when minimising the number of deaths and years of life lost, respectively. The mixing variables are presented as relative values compared to the pre-COVID-19 era for each location. A value of 1 represents unchanged location-specific contact rates compared to before the pandemic, whereas a value of 0.1 indicates a 90% reduction in contact rates. The tan-coloured background represents the acceptable region for the mixing factors (i.e. the interval [0.1 - 1]). The thin black bars represent the maximum change in individual age-group contributions that would cause an excess of no more than 20 deaths per million people (red bars) or 1000 YLLs per million people (blue bars) as compared to the optimal plan, while still reaching herd immunity by the end of the mitigation phase. The left panels show the result obtained when assuming that the mitigation phase lasts 6 months. In the right panels, a longer duration of 12 months was allowed to achieve herd immunity. The optimisations were performed based on the countries’ maximum a posteriori parameter sets.

### Projected epidemics

The trajectories of the optimised epidemics are shown in Figure 4. The younger populations were considerably more affected in the optimised strategies, resulting in a much lower ratio of deaths to incident disease episodes compared with the first wave. The median percentage of the population ever-infected with SARS-CoV-2 at the end of the simulation on 31 December 2021 ranged between 30% and 52%, depending on the country and the scenario considered (Table 2 and Figures S18-19). These percentages represent overestimates of herd immunity thresholds, since achieving herd immunity by the end of the intervention was a constraint of the optimisation algorithm. The percentages of the population ever-infected with SARS-CoV-2 are also presented by age-group in Figures S20-21.

**Figure 4.**
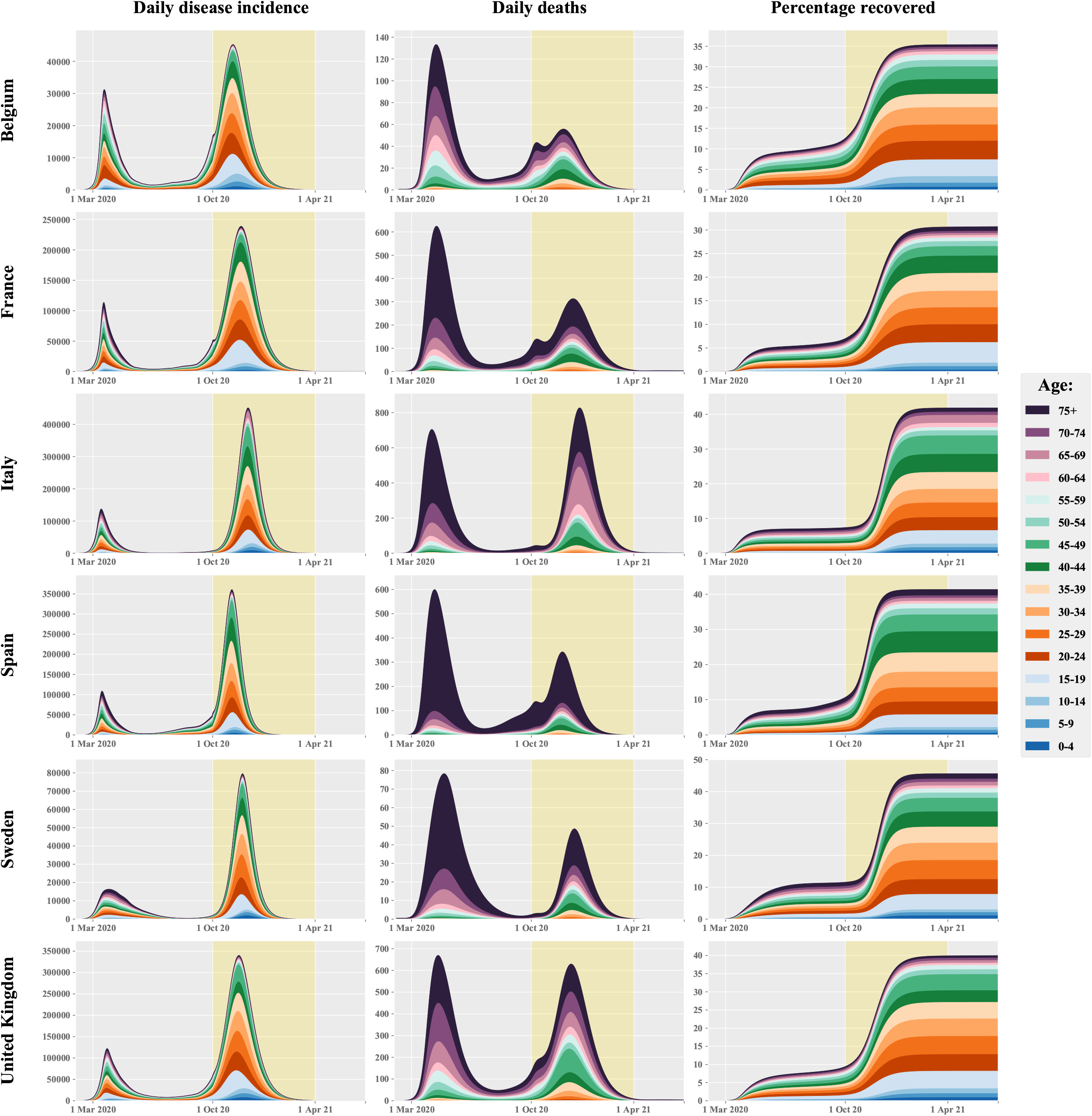
Age-specific profile of disease incidence, COVID-19-related deaths and proportion recovered over time optimised for life-years lost (6-month mitigation by age) The yellow background indicates the 6-month mitigation phase during which age-specific contacts were optimised. These projections were produced assuming that recovered individuals have persistent immunity against SARS-CoV-2 reinfection and using the maximum a posteriori parameter sets.

Our model projected that optimising social contacts by age could achieve herd immunity with lower or similar hospital occupancies to those observed in March and April in Belgium and France, whereas more hospital beds would be required in the other four countries (Figure 5). Our results also suggest that a longer mitigation phase could reduce the peak and total hospital burden, although the differences between the two durations in terms of COVID-19-related mortality and final epidemic size were minor (Tables 1 and 2).

**Figure 5.**
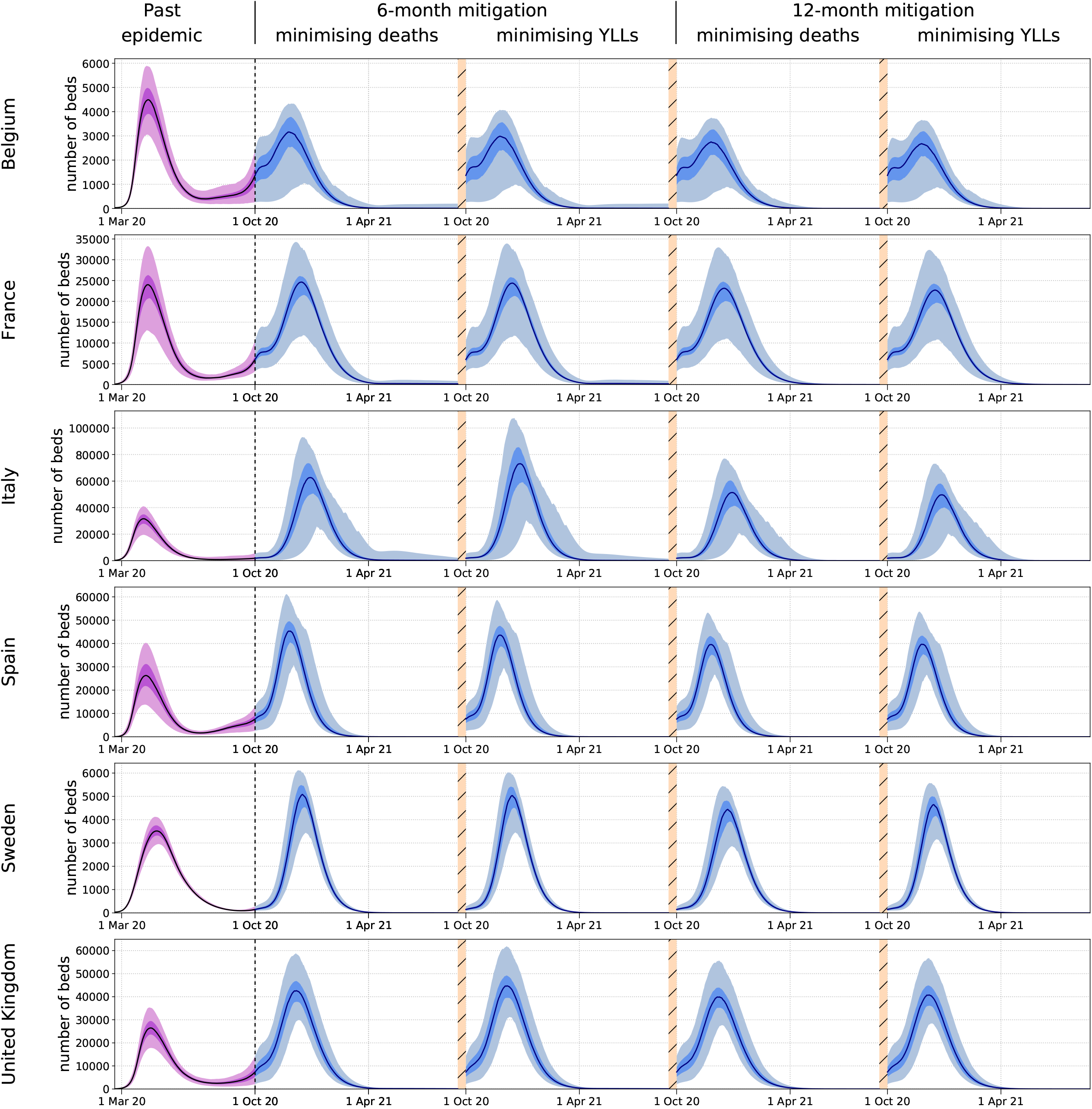
Projected hospital occupancy during the first wave compared to a mitigated wave that would achieve herd immunity (optimised mitigation by age, persistent immunity assumed) The modelled past epidemics are represented in purple while the projections of the mitigated epidemics are represented in blue. The future epidemics are those associated with the four different optimisation configurations: six- or 12-month mitigation minimising total number of deaths or years of life lost (YLLs). The light shades show the central 95% credible intervals, the dark shades show the central 50% credible intervals and the solid lines represent the median estimates.

### Effect of waning immunity

The four simulated scenarios of waning immunity considered different durations of post-infection immunity, as well as different assumptions regarding the effect of previous infection on disease severity for repeated infections. Using the six-month age-specific mitigation scenario, we found that under the four tested scenarios of waning immunity a third epidemic wave would occur by the end of 2021 in the absence of further intervention in all countries (Figure 6). The duration of post-infection immunity affected the future epidemics much more than the extent of protection against severe disease. Under the assumption of a six-month immunity duration, the predicted peak deaths and hospitalisations during epidemic resurgence were more than double those observed during the first wave or the mitigation phase in all countries.

**Figure 6.**
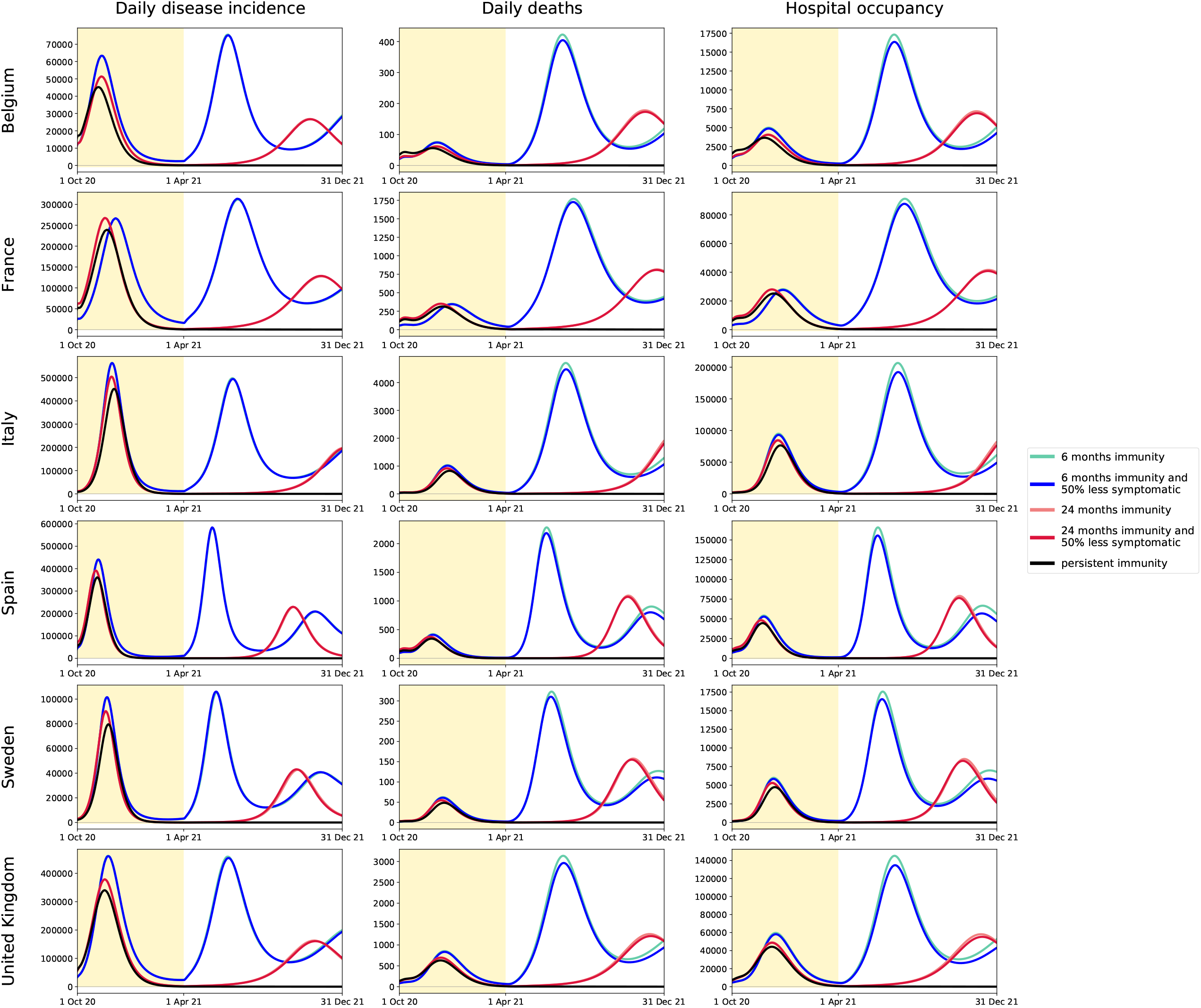
Projected COVID-19 incidence, mortality and hospital occupancy over time under various assumptions of waning immunity. Projections were obtained using the maximum a posteriori parameter sets and based on the 6-month contact mitigation by age minimising years of life lost (YLLs). The yellow background indicates the mitigation phase during which age-specific contacts were optimised. Five different assumptions were used to project the disease indicators: persistent immunity (black), 24-month average duration of immunity with and without 50% reduction in risk of symptoms for repeat infections (red and coral, respectively), 6-month average duration of immunity with and without 50% reduction in risk of symptoms for repeat infections (blue and turquoise, respectively).

Finally, we simulated scenarios considering our most pessimistic assumption of waning immunity but applying mild mixing restrictions after the mitigation phase (Figure 7). Our results suggest that relative mixing reductions of 30% would be required to maintain the epidemics at low levels until the end of 2021 in all countries except France and Sweden, where a 20% reduction may suffice to prevent significant resurgence.. These mixing reductions were defined as universal reductions relative to the pre-COVID-19 era.

**Figure 7.**
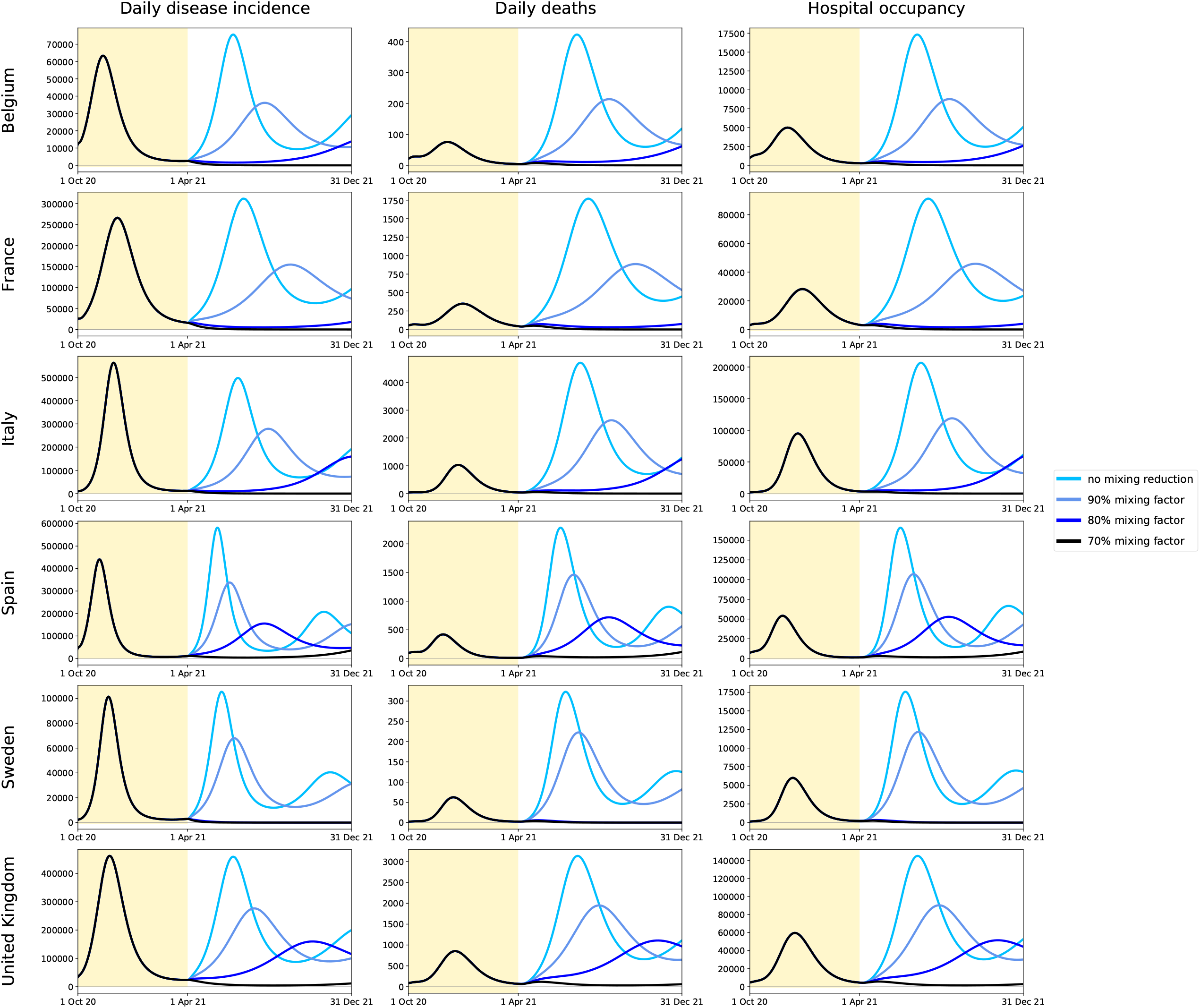
Projected COVID-19 incidence, mortality and hospital occupancy over time with short-lived post-infection immunity and applying mild mixing reductions after the optimised phase. The predictions were obtained using the maximum a posteriori parameter sets and based on the 6-month contact mitigation by age minimising years of life lost (YLLs). The yellow background indicates the mitigation phase during which age-specific contacts were optimised. These predictions were obtained assuming 6-month average duration of immunity with no effect on the severity of repeat SARS-CoV-2 infections. The mixing factors were defined in the same way as during optimisation except that the same factor was applied to all age-groups. That is, a 90% mixing factor corresponds to a situation where every individual reduces their opportunity of contact by 10%.

## Discussion

Our model suggests that altering age-specific mixing patterns can dramatically reduce the mortality-related impacts of COVID-19 over the medium-to-long term. We estimate that strategies based on contact mitigation by age could achieve non-vaccine herd immunity with mortality that is considerably lower than has been previously observed in Belgium, France, Spain and Sweden, whereas the numbers of deaths required to achieve herd immunity would be comparable to somewhat larger than the first wave in Italy and the UK. We also highlight the critical need for improved knowledge around post-infection immunity duration if such strategies are to be considered.

We quantify the contact patterns that are most likely to result in decreased mortality or YLLs, providing guidance on targeted release strategies. While many governments have used mitigation strategies based on location-specific restrictions, such as school or business closures, we demonstrate that strategies considering age-selective restrictions would have a greater impact on the countries’ epidemics, as the predicted number of future deaths was two-to-four times lower when optimising by age compared to optimising by location. Across all six countries included in our analysis, our model suggested that over 1 million deaths could potentially be averted, and over 20 million life-years saved, by employing age-specific mitigation compared to an unmitigated scenario. Such outcomes were obtained by imposing highly stringent contact reductions on individuals aged over 50, while returning interactions involving children and young-to-middle-aged adults to pre-COVID-19 levels in most countries. This age cut-off is lower than considered in previous studies investigating age-based shielding strategies^11,12^.

The stringency of the optimal restrictions on social contacts of people aged 50 years and over raises concerns about the feasibility of achieving the required age-differential mixing. For example, the optimised results we present rely on the assumption that the opportunity of effective contact could be reduced by up to 90%. It is to be noted that such reductions may be achieved by combining both mobility and gathering restrictions with reductions in the per-contact risk of transmission through preventive measures (i.e. micro-distancing, including masks, improved hygiene, physical distancing), such that interpersonal contacts would not need to be reduced to such an extreme degree. Nevertheless, even with micro-distancing, it may be impossible to achieve such reductions in many settings, including multigenerational households or residential aged care. Furthermore, we observed that the numbers of deaths and YLLs were highly sensitive to perturbations in some of the mixing variables, indicating that the strategies may rapidly become suboptimal if the targeted contact mitigation plan could not be implemented precisely. This indicates that the optimised number of deaths and YLLs should be interpreted as a representation of what could ideally be achieved under a best-case scenario. Practical implementation would require further analyses and critical consideration of tailored strategies to specific settings. Finally, our analyses considering less extreme restrictions showed that mortality would increase significantly compared to the optimised scenarios.

Our analyses showed that a longer duration of the optimised mitigation phase was associated with slightly improved outcomes compared to the shorter scenario. We note that this finding is intuitive, since the ensemble of acceptable mixing factor combinations associated with the shorter scenario (i.e. all mitigation strategies leading to herd immunity in 6 months) is necessarily included in that associated with the longer scenario (i.e. all mitigation strategies leading to herd immunity in 12 months). More practically, this is explained by the fact that more intense social mixing would be required if herd immunity had to be reached in a shorter period of time, which implicitly induced higher minimal levels on the mixing factors for the shorter scenario compared to the longer scenario. However, the benefits of increasing mitigation duration in terms of averted COVID-19 deaths should be weighed against the risks associated with extending the duration of social mixing manipulation.

It is notable that the final proportions of ever-infected individuals for each of the modelled scenarios in the six countries were between 30 and 52% in all six countries. Academic and public discussion has largely referred to a herd immunity threshold of between 60 and 70%^13,14^, a proportion that can be readily estimated from the basic reproductive number under the assumption of homogenous mixing. In reality, individuals differ as to how likely they are to contract and transmit SARS-CoV-2. Several other modelling incorporating heterogeneity have emerged and suggested lower estimates of the herd immunity threshold^15–18^. In particular, young-to-middle-age individuals, who have higher contact rates compared to older adults, contribute disproportionately to transmission, such that removing them from the susceptible pool would be disproportionately effective. Our findings also have important implications for vaccination strategies, as we estimated the age-specific proportions of recovered individuals after herd immunity was reached, providing examples of age-specific vaccination coverage that could result in herd immunity.

Our main projections were obtained assuming persistent post-infection immunity. Early studies have shown that most people infected with SARS-CoV-2 generate both humoral and cellular immune responses^19,20^. Some studies have shown antibody levels waning over the first three months post-infection, suggesting short-lived immunity^21^. However, other studies have shown neutralising antibodies to persist at protective levels three^22^, and six months post-infection^23^, as well as SARS-CoV-2-specific memory lymphocytes with characteristics suggestive of protective immunity up to three months post-infection^24^. A large-scale longitudinal seroprevalence study demonstrated robust humoral immune response, as antiviral antibodies against SARS-CoV-2 did not decline within four months after diagnosis^25^. Infection has been shown to offer protection against reinfection in non-human primates^26^, and recent evidence suggests similar protection in humans^27^. Although we considered waning immunity under only a limited number of configurations, these highlight the importance of immune effects and persistence to public health strategies.

We demonstrated that the selected countries’ health systems would be overwhelmed if immunity were to wane rapidly in the absence of any mitigation after the optimised phase. Long-term restrictions would be required under such scenarios, although such restrictions may consist of mild continuous contact mitigations following the optimised phase. For example, 30% continued reduction in effective contacts would be needed after the optimised phase in Belgium, Spain, Italy and the UK to maintain sufficient epidemic control until the end of 2021.

Our analysis raises the question of whether it would be ethical to restrict the freedom of a subset of the population, and to do so on the basis of age. Savulescu and Cameron argue that age-selective lockdowns would not constitute unjust discrimination, as it involves treating people differently due to a morally relevant difference: their susceptibility to severe infection^28^. They suggest that restrictions on personal freedoms would be most justified if they bring about benefit to the group whose freedoms are restricted. It is also possible that restriction of freedom of individuals could be reduced through the use of immunity passports, though these raise further ethical and practical issues^29^. Factors other than age also impact COVID-19 risk^30^. These factors are also important to consider when designing policies, although we do not explicitly account for these other risk factors.

The results for optimising for deaths or for YLLs were slightly different. This raises the question of what the optimisation target should be, and a welfare-adjusted life year, such as the quality-adjusted life year, may be preferable. Our analysis did not include the morbidity of illness or possible long-term sequelae of infection. Data on longer term outcomes suggest that most people who experience mild to moderate infections recover within two to three weeks^31^, although some experience prolonged symptoms or long-term sequelae^32^. These data will be essential to optimise for morbidity. It must be noted that our analysis was based on epidemiological indicators only and the optimisation trade-off was introduced by the competition between the infections required to increase population immunity and mortality that were minimised. Previous works also considered optimisation of COVID-19 control in a mathematical modelling framework. Perkins and España chose to minimise a single objective combining the number of deaths and the level of control by non-pharmaceutical interventions^33^. In another analysis, age-specific interventions were considered in an optimisation exercise based on a single objective function combining COVID-19-related mortality and the cost of the interventions and highlighted the benefit of age-targeted control^34^. However, the interpretation of these analyses is strongly dependent upon the definition of the intervention cost, which remains to be adequately quantified. Indeed, combining deaths and control cost into a single objective implies that these two components can be measured in the same unit, while their respective contributions to the objective remain extremely difficult to characterise.

Our approach has some technical limitations. We used previously published synthetic contact matrices, which allowed a consistent approach across the six countries and incorporation of location-specific contact rates^35^. The model assumes that mixing patterns in countries are well represented by these matrices and does not capture repeated contacts between the same individuals. We chose to mitigate the age-specific contact rates by applying a single multiplier to each age-group, whereas more flexibility could be introduced by allowing mixing between particular pairs of age-groups. Given the important uncertainties in the current epidemiological knowledge of SARS-CoV-2, we chose broad ranges of parameter values to inform the most critical aspects of the model, which translated into moderate uncertainty ranges for several epidemiological indicators. However, we believe our approach to handling uncertainty is appropriate to the current stage of the pandemic. Finally, the optimised strategies identified in this study may become suboptimal if the background epidemiological conditions changed significantly. In particular, we anticipate that as the epidemics progress, population immunity will naturally increase such that reduced levels of transmission may be required during the optimised phase to achieve herd immunity. However, the fact that we observed very similar optimal mixing patterns across the six countries, whereas the level of population immunity at the start of the mitigation phase ranged between 7% in Italy and 13% in Belgium, suggests stability in our findings about optimal strategies.

The present work could be refined as further knowledge arises about SARS-CoV-2 epidemiology, especially around the nature and magnitude of post-infection immunity. In addition, alternative optimisation frameworks to the one used in this study could be assessed in an attempt to further improve population outcomes. Finally, future work could include the negative effects of population restrictions more explicitly in order better to address the trade-off between restriction stringency and uncontrolled viral transmission.

Caution is also required in interpretation of our projections. It must be noted that the strategies we present would undoubtedly result in a greater number of COVID-19-specific deaths compared to approaches based on universal stringent restrictions to force the reproduction number below one. Accordingly, the risks and benefits of the strategies presented must be carefully weighed against those associated with universal lockdowns, which also have serious negative effects^2,3^. The right balance between these approaches will likely depend on how long we will have to wait until long-term solutions such as vaccines can be deployed and is a societal choice that should be informed by epidemiological analysis.

In conclusion, we found that strategies can minimise deaths or YLLs over the medium-to-long term while allowing an increase in population mixing if interpersonal contact patterns can be manipulated to prevent transmission to older adults. In particular, modification of contact rates by age is the key factor, although age-independent vulnerabilities also require consideration. We show the cut-off for contact restriction - analogous to shielding or cocooning - may occur at a younger age than previously assumed. Finally, our findings suggest that strategies combining a phase of age-selective contact restrictions designed to increase population immunity followed by ongoing but mild contact mitigation could maintain transmission at low levels even with short-lived post-infection immunity.

## Methods

### General approach

We used a compartmental model to simulate SARS-CoV-2 transmission in the six countries analysed. These were the six highest ranked countries in COVID-19 deaths per capita as reported by the World Health Organization on 15th July 2020, excluding countries of less than one million people. After calibrating the model using local data, we manipulated social mixing patterns for an intervention period of six or 12 months starting from 1^st^ October 2020. During this phase, we identified the changes to contact patterns that would minimise COVID-19-related mortality or YLLs over a time-horizon of 15 months (i.e. ending 31^st^ December 2021), while ensuring all restrictions could be relaxed after the intervention phase without resurgence. We also explored scenarios of waning immunity to project the future epidemics under the identified optimal plans.

### Transmission model

We explicitly simulated six infection states using a susceptible compartment, two pre-disease compartments (including one presymptomatic infectious), two disease states (early and late stages) and a recovered compartment (Supplementary Section 1.2). Infectious states were stratified according to severity, as well as estimated detection and hospitalisation fractions.

We employed age-specific parameter values to characterise susceptibility to infection, disease severity and risk of death (Table S3). We used previously published age-specific contact matrices by location (home, schools, workplace, other locations) to inform heterogeneous mixing by age^35^.

We modelled physical distancing by reducing the location-specific contact rates in the three non-household locations. We also included micro-distancing by reducing the transmission probability in non-household contacts, reflecting preventive measures taken to reduce the per-contact transmission probability, such as keeping a greater distance, hygiene measures, and wearing masks.

Under the base-case assumption of persistent immunity, recovered individuals were assumed to be permanently protected against future infection. Four scenarios of waning immunity were also considered by assuming that recovered individuals became susceptible to reinfection after an average duration of six or 24 months, with or without reduced disease severity during repeat SARS-CoV-2 infections (Supplementary Sections 1.2, 1.4). The model code is publicly available on Github^36^.

### Model fitting and simulation phases

We fitted the model to observed numbers of confirmed cases, hospitalisations and deaths over time. Seroprevalence data were also included as calibration targets when available (Supplement Section 2.2). Fitted parameters included those governing transmission, disease severity and the time-variant profiles of case detection and micro-distancing. Our simulations were divided into three successive phases (Figure 8). In Phase 1 we modelled the preceding SARS-CoV-2 epidemics and included social distancing measures in place in each country until 31st September 2020 (Figure S4). In Phase 2 the model was run using the same epidemiological parameters and detection profile as during Phase 1, but social mixing interventions were optimised for six or 12 months, before being lifted in Phase 3. We assumed that mild micro-distancing was maintained during Phases 2 and 3 to capture the likely long-term changes in individuals’ behaviours and the preventive measures undertaken in the future, as public awareness of the modes of transmission of SARS-CoV-2 increases relative to the early stages of the epidemics (Supplement Sections 3.2 and 3.3).

**Figure 8.**
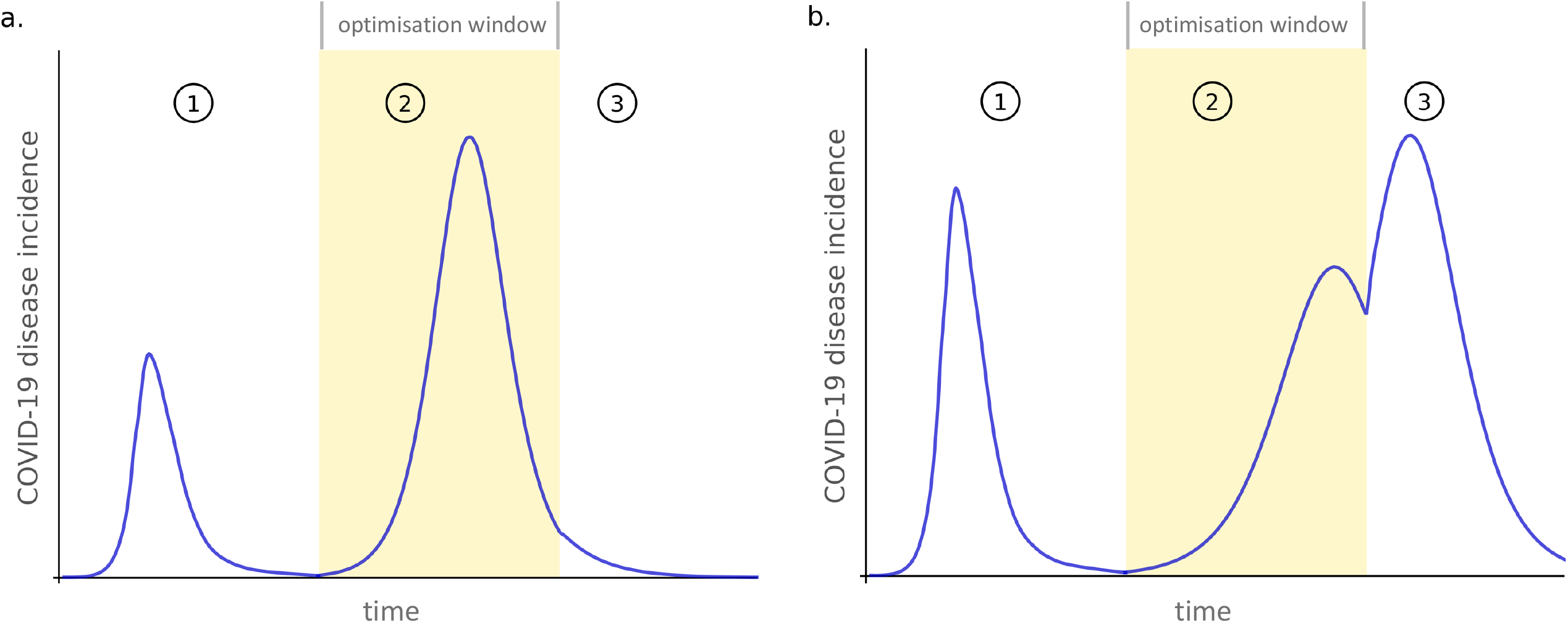
Illustration of the three simulation phases. Numbered circles indicate the different phases: capturing past dynamics (1), manipulating social mixing to achieve herd immunity with minimum COVID-19 impacts (2, highlighted with yellow background), testing for epidemic resurgence (3). Panel a. shows an example simulation where herd immunity was reached by the end of Phase 2, whereas Panel b. shows a configuration that failed to achieve herd immunity.

### Optimising contact patterns

We used two different indicators to represent the disease impact in separate optimisations: the number of COVID-19-related deaths and the number of YLLs due to COVID-19-related deaths. The number of YLLs was estimated using the country-specific life-expectancy values by age reported by the United Nations. The two objective functions were calculated over a time-horizon of 15 months covering Phases 2 and 3. Two types of mitigation strategies were explored. First, we allowed contact rates to vary by age by applying age-specific mixing factors to the original contact matrix (Supplementary Section 3.2). A mixing factor is defined as the relative opportunity of social contact that an individual of a given age has, compared to the pre-COVID-19 era. Therefore, the relative contact rate of one age category with respect to another is calculated as the product of the mixing factors of the age-groups of the infectious and susceptible individuals. In separate analyses, we considered reductions in social mixing by location where the decision variables were scaling factors applying to the location-specific contact rates.

All decision variables were assumed to be bounded between 0.1 and one and the optimisation was also constrained to solutions in which the number of incident cases did not increase after the mitigation phase. Because of this constraint, significant transmission was necessary during the optimised mitigation phase in order to increase the level of population immunity, introducing a trade-off between the required SARS-CoV-2 infections and the COVID-19-related deaths to be minimised. As the optimisation tasks were computationally-expensive, the searches were performed using a Genetic Algorithm where the newly generated candidate strategies were evaluated in parallel on multiple CPUs^37^.

### Sensitivity analyses

In order to test the sensitivity of the objective functions to alterations of each optimised variable, we calculated the marginal variable deviation from the optimum that would cause an excess of 20 deaths per million people (or 1000 YLLs per million people when minimising YLLs) as compared to the optimum (Supplementary Section 4.3). An additional sensitivity analysis was performed considering values greater than 0.1 for the lower bound of the mixing factors (Supplementary Section 5.1).

## Data Availability

The study was conducted using publically available data only. The references are presented in the manuscript.

## Contributors

RR, ESM and JMT conceived the study with input from BMW, BJM and RME. RR conducted the analyses. RR, JMT, MS and MA developed the code. GB, NM and DT designed the optimisations and GB led their implementation. RR, BMW and JMT wrote the first draft and all authors contributed to the final draft.

## Declaration of interests

The authors declare no competing interests.

## Data availability

No individual data were used in this study. The sources of the country-level data used to calibrate our models are indicated in the Supplement.

## Code availability

The computer code used in this study is available in a GitHub repository: https://github.com/monash-emu/AuTuMN

## Acknowledgements

The optimisation experiments were performed using the Grid’5000 testbed, supported by a scientific interest group hosted by Inria and including CNRS, RENATER and several Universities as well as other organisations (see https://www.grid5000.fr).

## Notes

### Competing Interest Statement

The authors have declared no competing interest.

### Funding Statement

RR was supported by a Project Grant (APP1144570) from the Australian National Health and Medical Research Council. JMT was supported by an Early Career Fellowship (APP1142638) from the Australian National Health and Medical Research Council.

### Summary of Updates

Revised version of the manuscript after addressing reviewers' comments.

