## Supplement for "Optimising social mixing strategies achieving COVID-19 herd immunity while minimising mortality in six European countries"

### Optimising social mixing strategies to mitigate the impact of COVID-19 in six European countries

### 1 Model description

#### 1.1 Base platform

We developed a deterministic compartmental model of SARS-CoV-2 transmission using the AuTuMN platform, publicly available at github.com/monash-emu/AuTuMN [1]. This repository allows for the rapid and robust creation and stratification of models of infectious disease epidemiology and includes pluggable modules to simulate heterogeneous population mixing, demographic processes, multiple circulating pathogen strains, repeated stratification and other modelling features relevant to infectious disease transmission.

#### 1.2 Base model

For this application, we first created a model with sequential compartments representing susceptible ($S$), latently infected ($E$), infectious pre-symptomatic ($P$), early disease ($I$), late disease ($L$) and recovered ($R$) persons (although patients in the early and late disease compartments are not considered symptomatic if assigned to the first "clinical" stratification, as described in Section 1.4). The latently infected and infectious pre-symptomatic periods together comprise the incubation period, with the incubation period and the proportion of this period for which patients are infectious defined by parameters described below. The transition from early disease to late disease is intended to represent the point at which patients are detected (in the event that detection does eventually occur) and isolation then occurs from this point forward (i.e. applies during the late disease phase only). This transition point is also intended to represent the point of admission to hospital or to intensive care for patients for whom this occurs (again see Section 1.4).

When waning immunity was assumed, individuals transitioned back to a susceptible compartment at a rate that was defined as the reciprocal of the assumed immunity duration. The compartments $S$ and $E$ were stratified by infection history, such that differential risks of severe disease could be considered for individuals who have experienced SARS-CoV-2 infection before, compared to infection-naive individuals.

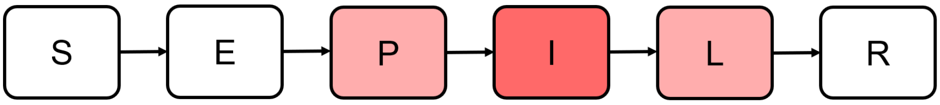

**Figure S1. Compartmental structure of the base model.**
Red shading indicates infectiousness.

#### 1.3 Age stratification

All compartments of this base compartmental structure were stratified by age into five-year bands from 0-4 years of age through to 70-74 years of age, with the final age group being those aged 75 years and older.

We used the age-specific contact matrices by location (home, schools, workplace and other locations) reported by Prem *et al.* for the five investigated countries to inform heterogeneous mixing in our models [2]. The model age groups were chosen to match these mixing matrices. We did not implement births, ageing and non-COVID-19-related deaths given that the current study pertains to the short to medium-term and the immediate implementation of non-pharmaceutical interventions, for which population demographics are less relevant.

#### 1.4 Clinical stratification

The age-stratified pre-symptomatic and disease compartments $P$, $I$ and $L$ were further stratified into five categories: 1) asymptomatic, 2) symptomatic ambulatory never detected, 3) symptomatic ambulatory ever detected, 4) hospitalised never critical and 5) ever critically unwell. The proportion of new infectious persons entering stratum 1 (asymptomatic) is age-dependent as described in Table S3. When waning immunity was assumed, these proportions also depended on the individuals' infection history, such that a smaller proportion of symptomatic infections may be considered for repeat episodes of SARS-CoV-2 infection. The proportion of symptomatic patients (strata 2 to 5) ever detected (strata 3 to 5) is set through a time-variant parameter that represents the proportion of all symptomatic patients who are ever detected. Of those ever symptomatic (strata 2 to 5), an age-specific proportion was considered to be hospitalised (entering strata 4 or 5). Of those hospitalised (entering strata 4 or 5), a fixed proportion was considered to be critically unwell (entering stratum 5). The figures below illustrate this conceptual approach.

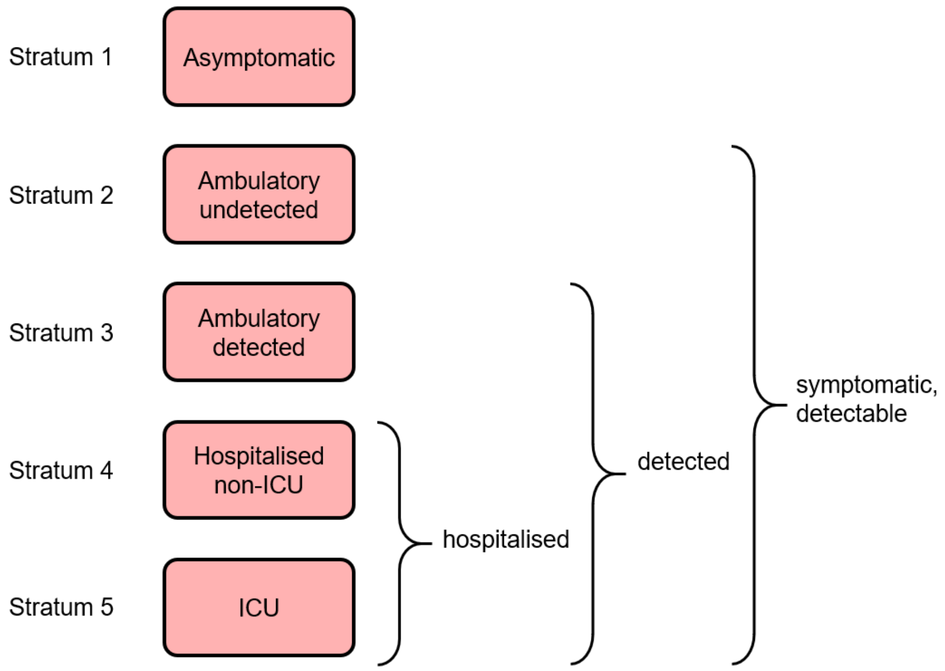

**Figure S2. Clinical stratification applying to compartments $P$, $I$ and $L$.**

#### 1.5 COVID-19-related death

We used age-specific infection fatality rates (IFRs) to model COVID-19 deaths. These rates were previously estimated using age-specific death data from 45 countries and results from national-level seroprevalence surveys [3,4]. In our models, we used the reported point estimates to which we applied an uncertainty multiplier in order to reflect the fact that IFRs may vary by country and because COVID-19 deaths may be reported differently in different countries. The prior distribution range associated with this multiplier in our uncertainty analysis was obtained by considering the amplitude of between-country variations reported in a previous analysis [5].

Age-specific IFRs were applied and distributed across strata 4 and 5, with no deaths

| Clinical stratum | Pre-symptomatic | Early disease | Late disease |
|---|---|---|---|
| Asymptomatic | 0.5 | 0.7 | 0.7 |
| Ambulatory undetected | 0.7 | 1 | 1 |
| Ambulatory detected | 0.7 | 1 | 0.2 |
| Hospitalised non-ICU | 0.7 | 1 | 0.2 |
| ICU | 0.7 | 1 | 0.2 |

**Table S1. Illustration of the relative infectiousness of disease compartments by clinical stratification and stage of infection.**
Darker shades of red indicate higher levels of infectiousness.

typically applied to the first three strata. If the IFR was greater than half of the absolute proportion of persons critically unwell (entering stratum 5), the proportion of critically unwell persons dying was set to one half, with the remainder of the IFR then applied to the hospitalised proportion. Otherwise, if the IFR is less than half of the absolute proportion of persons critically unwell, the IFR is applied entirely through stratum 5 (such that the proportion of critically unwell persons dying in that age group becomes <50% and the proportion of stratum 4 dying is set to zero). In the event that the IFR for an age group is greater than the total proportion hospitalised (which is unusual, but could occur for the oldest age group under certain parameter configurations), the remaining deaths are assigned to the asymptomatic stratum. This approach was chosen because this stratum has a fixed value over time and the dynamics are equivalent to assigning the deaths to any of the first three strata, because the sojourn times in the infected compartments are the same for each of these groups.

#### 1.6 Infectiousness

For patients with disease who were admitted to hospital, the sojourn time in the early and late infectious compartments was modified, as indicated in Table S2. The point of admission to hospital was considered to be the transition from early to late infectious disease, such that the sojourn time in late disease was the period of time admitted to hospital. For patients admitted to ICU, admission to ICU occurs at this same transition point. For this group, the period of time hospitalised outside of ICU is estimated as a proportion of the early active period, such that the early active period represents both the period ambulatory in the community and the period in hospital prior to ICU admission. Infectiousness declined at the point of transition from early to late disease for all patients admitted to hospital (both ICU and non-ICU) and for patients who were effectively detected and so underwent isolation.

The relative infectiousness of both early and late compartments within the asymptomatic stratum, as well as the late disease compartment of the symptomatic ambulatory detected late disease were modified. This was intended to reflect lower infectiousness per unit time of asymptomatic persons and of detected persons who were assumed to self-isolate from the point of entering the late disease compartment. Pre-symptomatic individuals were assumed to have the same level of infectiousness as asymptomatic diseased individuals, although pre-symptomatic individuals of the asymptomatic stratum were assumed less infectious. Persons with late stage disease in the hospital and critical strata also had their infectiousness modified. Table S1 illustrates the different levels of infectiousness associated with the different clinical strata and the different infection stages.

#### 1.7 Implementation of non-pharmaceutical interventions

For this study, it was critical to simulate the past dynamics of the epidemics accurately in order to capture the level of immunity acquired in the past. For this reason, we needed to capture the past impact of non-pharmaceutical interventions (NPIs) such as social distancing or school closures.

##### 1.7.1 Isolation and quarantine

For persons who are identified with symptomatic disease and enter clinical stratum 3, self-isolation is assumed to occur. The proportion of ambulatory symptomatic persons effectively identified through the public health response by any means is set by the time-variant process described in Section 1.7.2. On moving to stratum 3, infectiousness declines according to the "relative infectiousness of identified persons undergoing quarantine and isolation" parameter indicated above.

##### 1.7.2 Modelling time-variant detection

The proportion of symptomatic individuals detected was varied over time in order to account for increasing detection during the course of the epidemic. We used the following translated and rescaled logistic function to model this increase:

$$prop_{sympt\_detected}(t) = prop_{start} + \frac{prop_{final} - prop_{start}}{2} \left( tanh\left( b\left( t - c \right) \right) + 1 \right),$$

where $prop_{start}$ is the proportion of symptomatic individuals that are detected at the start of 2020, $prop_{final}$ is the maximum asymptotic proportion of symptomatic individuals that are detected, $b$ is a shape parameter defining how rapidly scaling occurs and $c$ is the time when inflection occurs in the scale-up curve. The parameters $prop_{final}$, $b$ and $c$ are automatically estimated during model calibration (see Section 2). Figure S3 illustrates two examples of functions describing the proportion of symptomatic individuals that are detected over time. The six countries' detection profiles inferred from the detection parameters' posterior distributions are presented in Section 2.4.

##### 1.7.3 Other non-pharmaceutical interventions

For all NPIs relating to reduction of human mobility or "lockdown", these interventions are implemented through dynamic adjustments to the age-assortative mixing matrix. The mixing matrices of Prem et al. allow for disaggregation of total contact rates by location, i.e. home, work, school and other locations. This disaggregation allows for the simulation of various NPIs in a local context by dynamically varying the contribution of each location to reflect the historical implementation of the interventions. The corresponding mixing matrix (denoted $C_0$) is presented using the standard convention that a row represents the average numbers of age-specific contacts per day for a contact recipient of a given age-group. In other words, the element $C_0[i, j]$ is the average number of contacts per day that an individual of age-group $i$ has with individuals of age-group $j$. This matrix results from the summation of the four context-specific contact matrices: $C_0 = C_H + C_S + C_W + C_L$, where $C_H$, $C_S$, $C_W$ and $C_L$ are the age-specific contact matrices associated with households, schools, workplaces and other locations, respectively. In our model, the

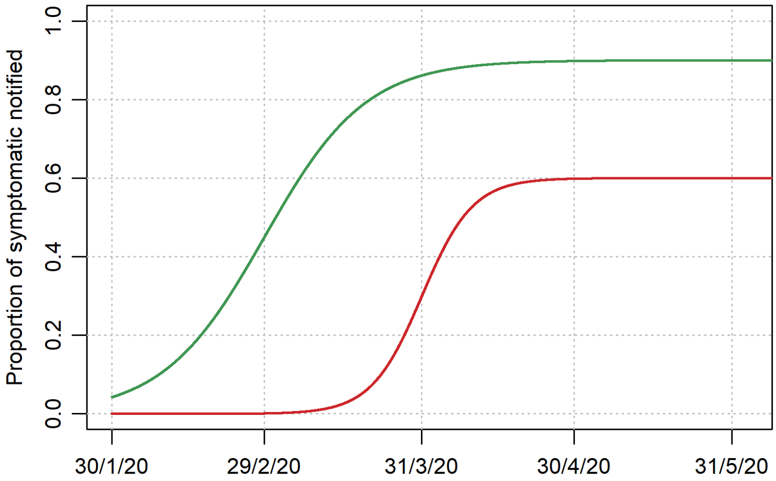

**Figure S3. Examples of modelled time-variant proportions of symptomatic individuals detected over time.**
Two parameter sets are illustrated: ($prop_{start} = 0$, $prop_{final} = 0.6$, $b = 0.1$, $c = 31/3/2020$) in red; ($prop_{start} = 0$, $prop_{final} = 0.9$, $b = 0.05$, $c = 29/2/2020$) in green.

contributions of the matrices $C_S$, $C_W$ and $C_L$ are time-variant, such that the input contact matrix can be written:

$$C(t) = C_H + s(t)^2 C_S + w(t)^2 C_W + l(t)^2 C_L, \tag{1}$$

where $s$, $w$ and $l$ are location-specific time-variant multipliers. Note that these multipliers were squared to capture the fact that both individuals involved in a contact are affected by the relative probability of being in a given location. The following three sections describe how these multipliers are set and Figure S4 shows their respective profiles for the six countries.

It is important to note that $s$, $w$, $l$ are all set back to 1 when predicting the future epidemic during Phase 3 as well as during Phase 2 when mixing is optimised by age (see Section 3 for description of the different phases).

##### School closures

School closures were represented by decreasing the contribution of the school-based contacts to the mixing matrix by 90% at the time school closures occurred in the country considered. The closure and reopening times were obtained from the UNESCO website [6]. The reason why the school contacts were not set to zero after closure is that many schools continued to accept some children whose parents worked in priority sectors.

##### Workplace closures

Workplace closures were represented by reducing the contribution of workplace contacts to the total mixing matrix over time. We used Google mobility data to inform this time-variant reduction after applying a 7-day moving average to the raw data [7].

##### Community-wide movement restriction

This was simulated by reducing the contribution of the "other location" contacts to the overall mixing matrix. The functional form of this reduction was set using Google mobility data and obtained by combining the proportional reductions reported by Google for the three categories "Retail and recreation", "Supermarket and pharmacy" and "Public transport" [7]. We did not include the relative changes of the "Parks" category, as their

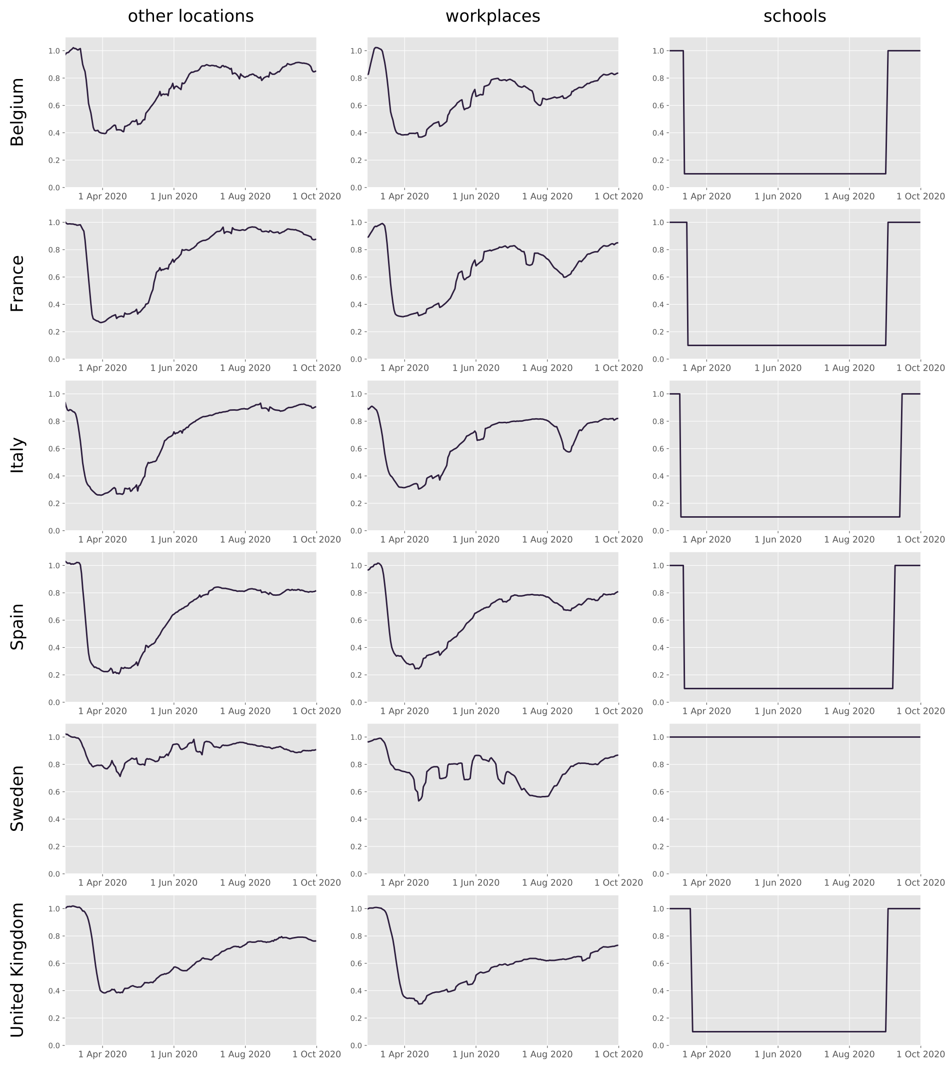

**Figure S4. Time-variant contact rate multipliers used to model mobility restrictions.** "Other locations" refers to all places other than schools, workplaces or households.

contribution to transmission is expected to be minimal. The reduction applied to the "other locations" contact rates $\rho := 1 - l$, was obtained from:

$$\rho(t) = \frac{\rho_R(t) + \rho_S(t) + \rho_P(t)}{3},$$

where $\rho_R$, $\rho_S$ and $\rho_P$ are the proportional reductions reported by Google for "Retail and recreation", "Supermarket and pharmacy" and "Public transport", respectively. We applied a 7-day moving average to the raw data for smoothing.

**Micro-distancing**

The previous adjustments to social mixing reflected reductions in people's mobility that resulted in lower rates of contact. In addition to this, we apply a time-variant reduction to the per-contact probability of transmission in locations other than households. This adjustment is referred to as micro-distancing and is intended to capture physical distancing between individuals, mask wearing and other preventive measures that individuals may take to reduce the per-contact transmission risk. Micro-distancing was modelled using the combination of an increasing function of time that reflects the progressive emergence of micro-distancing, and a decreasing function that captures potential waning effects such as decreasing compliance over time. The increasing component was implemented as follows:

$$micro_{emergence}(t) = \frac{micro_{final}}{2}\left(tanh\left(0.05\left(t - micro_{inflection}\right)\right) + 1\right),$$

where $micro_{final}$ represents the final value of the emergence component and $micro_{inflection}$ is the time when inflection occurs in the scaling curve. The waning component was modelled as follows:

$$micro_{wane}(t) = wane_{final} + \frac{1 - wane_{final}}{2}\left(tanh\left(-0.05\left(t - wane_{inflection}\right)\right) + 1\right),$$

where $wane_{final}$ represents the final value of the emergence component and $wane_{inflection}$ is the time when inflection occurs in the decreasing curve.

Finally, the multiplier used in the model to adjust the per-contact risk of transmission was obtained by combining the two components as follows:

$$m_d(t) = 1 - micro_{emergence}(t) \times micro_{wane}(t).$$

The parameters $micro_{final}$, $micro_{inflection}$, $wane_{final}$ and $wane_{inflection}$ were automatically inferred during calibration. Figure S5 illustrates an example of micro-distancing profile.

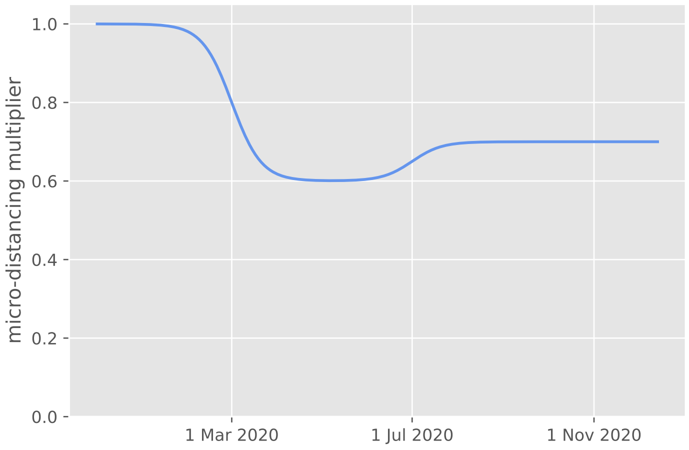

**Figure S5. Example of time-variant micro-distancing profiles.**
The following parameter set was used: $micro_{final} = 0.4$, $micro_{inflection} = 1/3/2020$, $wane_{final} = 0.75$, $wane_{inflection} = 1/7/2020$.

##### Age-specific behavioural changes

Our early attempts at fitting the model to data on both the first epidemic wave and the second epidemic take-off demonstrated that the dramatic change observed in the ratio between COVID-19 deaths and notifications after the first wave could not be explained by improvements in case detection alone. In order to capture these significant changes, we assumed that there may have been some age-specific behavioural changes after the first wave, whereby older individuals were more likely to reduce their social interactions compared to younger individuals.

To implement these age-specific variations, we allowed the contact rates of the elderly population (60 years old and over) to be reduced by up to 50%, with the reduction occurring progressively between 1 April 2020 and 31 May 2020. The magnitude of this reduction was automatically inferred during calibration (see Table S2).

#### 1.8 Model equations

The dynamics of the model are governed by a set of ordinary differential equations. We use the subscripts $a$, $h$ and $c$ to denote the different age-groups, the two infection history statuses and the clinical strata, respectively. That is, $a$ is an element of {"0-4", "5-9", ..., "70-74", "75+"}, $h$ is an element of {"infection-naive", "previously infected") } and $c$ is one of {"asymptomatic", "ambulatory undetected", "ambulatory detected", "hospitalised non-ICU", "ICU"}.

Using the compartment notation introduced in Section 1.2 and the parameter notation presented in Table S4, we have:

$$\begin{cases} \dot{S}_{a,h} & = -\lambda_a(t)\sigma_a S_{a,h} + \omega R_a \mathbb{1}_{\{h=\text{"}previously\, infected\text{"}\}} \\ \dot{E}_{a,h} & = \lambda_a(t)\sigma_a S_{a,h} - \alpha E_{a,h} \\ \dot{P}_{a,c} & = \sum_h p_{a,h,c}(t)\, \alpha E_{a,h} - \nu P_{a,c} \\ \dot{I}_{a,c} & = \nu P_{a,c} - \gamma_c I_{a,c} \\ \dot{L}_{a,c} & = \gamma_c I_{a,c} - \delta_{a,c} L_{a,c} - \mu_{a,c} L_{a,c} \\ \dot{R}_a & = \sum_c \delta_{a,c} L_{a,c} - \omega R_a \ , \end{cases} \tag{2}$$

where:

$$\begin{cases} \lambda_a(t) & = \beta \left[ \sum_j \frac{\epsilon \times P_j}{N_j} K_{a,j}(t) \ + \ \sum_{j,c} \frac{\iota_c \times I_{j,c} \, + \, \kappa_c \times L_{j,c}}{N_j} K_{a,j}(t) \right], \\ K_{a,j}(t) & = C_{H\,a,j} + (m_d(t)s(t))^2 C_{S\,a,j} + (m_d(t)w(t))^2 C_{W\,a,j} + (m_d(t)l(t))^2 C_{L\,a,j}, \\ \sum_c p_{a,h,c}(t) & = 1, \quad \forall t \in \mathbb{R}, \ \forall h \in \{\text{"}infection{-}naive\text{"}, \text{"}previously\, infected\text{"}\}. \end{cases} \tag{3}$$

| Parameter | Definition | Value/Prior distribution | Source |
|---|---|---|---|
| Epidemic seeding time[*,c] | Time when first infectious individuals were introduced in the model | Uniform on [0, 40] | Assumption |
| Transmission probability per contact[c] | Probability of transmission per contact between a fully-infectious and a fully-susceptible individual | Uniform on [0.02 - 0.06] | Early model explorations |
| Incubation time[c] | Average total duration spent in pre-disease infection compartments ($E$ and $P$) | Normal(5.5, 1) lower bound: 1 day | [8–11] |
| Incubation infectious proportion | Proportion of incubation period infectious | 0.5 | [12] |
| Disease duration[c] | Average total duration spent in the disease compartments ($I$ and $L$). Does not apply to hospitalised patients. | Normal(6.5, 0.8) lower bound: 1 day | [11–13] |
| Early disease proportion (non-hospital) | Proportion of disease period before isolation can occur for individuals who are never hospitalised | 0.33 | Assumed (i.e. that identification and isolation can only occur after the first third of the disease episode has elapsed) |
| Early disease duration (hospital non-ICU) | Disease duration prior to admission for hospitalised patients not critically unwell | 7.7 days | Expected mean from ISARIC cohort, as reported on 4th Oct 2020 [14] |
| Early disease duration (ICU) | Disease duration prior to admission for ICU patients | 10.5 days | Calculated as 7.7 days prior to hospital admission plus 2.8 days in hospital prior to ICU admission. Former value being as for disease duration prior to admission for hospitalised patients not critically unwell. Latter value being expected mean of period from admission to ICU entry from ISARIC cohort, as reported on 4th Oct 2020 [14]. |
| Late disease duration (hospital) | Average hospitalisation duration, excluding ICU | 12.8 days | Expected mean from ISARIC cohort, as reported on 4th Oct 2020 [14] |
| Late disease duration (ICU) | Average duration in intensive care unit | 13.3 days | Mean duration of stay in ICU/HDU from ISARIC cohort, as reported on 4th Oct 2020 [14] |
| Symptomatic proportions uncertainty[c] | Multiplier applying to the age-specific proportions of symptomatic presented in Table S3 | Uniform on [0.6 - 1.4] | Assumption |
| Hospital proportions uncertainty[c] | Multiplier applying to the age-specific proportions of hospitalised presented in Table S3 | Uniform on [0.5 - 1.5] | Assumption |
| ICU proportion | Proportion of hospitalised individuals admitted to intensive care | 0.17 | [14] |
| Infection fatality rates | Age-specific proportions of death among infected individuals | See Table S3 | [3, 4] |
| Infection fatality rate multiplier[c] | Uncertainty multiplier applied to the age-specific infection fatality rates presented in Table S3 | Uniform on [0.5 - 3.8] | [5] |
| Elderly protection after the first wave[c] | Relative contact reduction scaling progressively between 1 April and 31 May 2020 | Uniform on [0 - 0.5] | Assumption |
| Detection profile:<br>→ $prop_{start}$[c]<br>→ $prop_{final}$[c]<br>→ $b$[c]<br>→ $c$[*,c] | See Section 1.7.2. | Uniform on<br>[0 - 0.3]<br>[0.10 - 0.99]<br>[0.03 - 0.15]<br>[100 - 250] | Assumption |
| Micro-distancing:<br>→ $micro_{final}$[c]<br>→ $micro_{inflection}$[*,c]<br>→ $wane_{final}$[c]<br>→ $wane_{inflection}$[*,c] | See Section 1.7.3 | Uniform on<br>[0.25 - 0.8]<br>[60 - 130]<br>[0.4 - 1]<br>[130 - 250] | Assumption |

**Table S2. Model parameters.**
*times are expressed as number of days since 31 December 2019. [c]Parameter included in the Bayesian calibration.

<table>
<tr><th>Age group</th><th>Proportion symptomatic$^{(u)}$</th><th>Relative susceptibility to infection</th><th>Proportion hospitalised among symptomatic$^{(u)}$</th><th>Infection fatality rate$^{(u)}$</th></tr>
<tr><td>0-4</td><td rowspan="2">0.29</td><td rowspan="3">0.36</td><td>0.087</td><td>$2e^{-5}$</td></tr>
<tr><td>5-9</td><td>0.009</td><td>$5e^{-6}$</td></tr>
<tr><td>10-14</td><td rowspan="2">0.21</td><td>0.005</td><td>$5e^{-6}$</td></tr>
<tr><td>15-19</td><td rowspan="10">1</td><td>0.008</td><td>$1.5e^{-5}$</td></tr>
<tr><td>20-24</td><td rowspan="2">0.27</td><td>0.011</td><td>$4e^{-5}$</td></tr>
<tr><td>25-29</td><td>0.011</td><td>$9e^{-5}$</td></tr>
<tr><td>30-34</td><td rowspan="2">0.33</td><td>0.013</td><td>$1.7e^{-4}$</td></tr>
<tr><td>35-39</td><td>0.015</td><td>$2.9e^{-4}$</td></tr>
<tr><td>40-44</td><td rowspan="2">0.40</td><td>0.018</td><td>$5.3e^{-4}$</td></tr>
<tr><td>45-49</td><td>0.040</td><td>$8.6e^{-4}$</td></tr>
<tr><td>50-54</td><td rowspan="2">0.49</td><td>0.047</td><td>$1.5e^{-3}$</td></tr>
<tr><td>55-59</td><td>0.070</td><td>$2.4e^{-3}$</td></tr>
<tr><td>60-64</td><td rowspan="2">0.63</td><td>0.071</td><td>$3.6e^{-3}$</td></tr>
<tr><td>65-69</td><td rowspan="4">1.41</td><td>0.114</td><td>$6.4e^{-3}$</td></tr>
<tr><td>70-74</td><td rowspan="3">0.69</td><td>0.113</td><td>0.01</td></tr>
<tr><td>75-79</td><td rowspan="2">0.071</td><td>0.02</td></tr>
<tr><td>80 and above</td><td>0.07</td></tr>
<tr><td>Source</td><td>Davies et al. [15]</td><td>Zhang et al. [16]</td><td>Netherlands RIVM weekly report [17]</td><td>O'Driscoll et al. [3]</td></tr>
</table>

**Table S3. Age-specific parameters.**
$^{(u)}$Uncertainty was incorporated around this parameter by applying a multiplier that was varied in the adaptive Metropolis algorithm (see Table S2).

| parameter | definition |
|---|---|
| $\sigma_a$ | relative susceptibility to infection by age |
| $\alpha$ | rate of progression from latent to pre-symptomatic state |
| $\nu$ | rate of progression from pre-symptomatic to early disease state |
| $p_{a,c}$ | age-specific proportion progressing to each clinical stratification |
| $\gamma_c$ | rate of progression from early active disease to late active disease |
| $\delta_{a,c}$ | rate of progression from late active disease to recovered state |
| $\omega$ | rate of immunity loss |
| $\mu_{a,c}$ | rate of COVID-19-related mortality |
| $\beta$ | probability of infection per contact between a fully-infectious and a fully-susceptible individual |
| $m_d$ | time-variant multiplier used to model micro-distancing (Section 1.7.3) |
| $\epsilon$ | relative infectiousness of pre-symptomatic individuals |
| $\iota_c$ | clinical stratification infectiousness vector for early disease compartments |
| $\kappa_c$ | clinical stratification infectiousness vector for late disease compartments |
| $C_{a,j}$ | contact matrix element $[a, j]$, defined as the average daily number of persons of age $a$ contacted by an individual of age $j$. |

**Table S4. Parameter notation for ordinary differential equations.**

#### 2 Model calibration

The model was calibrated using a Bayesian approach. In particular, we used the adaptive Metropolis algorithm introduced by Haario *et al.* to sample parameters from their posterior distributions [18]. For each country, we ran 7 independent Metropolis chains initialised using Latin Hypercube Sampling based on the parameter priors. We ran simulations for 8 hours per chain in order to achieve at least 5,000 iterations per chain. We discarded the first 2,500 iterations of each chain as burn-in and combined the samples of the 7 chains to project epidemic trajectories over time. The definitions of the prior distributions and the likelihood are detailed in the following sections.

##### 2.1 Parameters varied during calibration

The parameters varied during calibration along with their associated prior distributions are listed in Table S2 and indicated with the superscript $^{c}$. We used uniform prior distributions for the most uncertain parameters and truncated normal distributions for those that have been better characterised in previous works.

##### 2.2 Calibration targets

We used the data reported by the World Health Organization for the daily numbers of COVID-19 confirmed cases and deaths between 1 March 2020 and 30 September 2020 in the six countries considered [19]. We then converted the daily counts into weekly totals in order to remove the effect of weekdays versus weekends in cases reporting. We also used hospitalisation data as calibration targets. We fitted the model to daily numbers of new hospitalisations (7-day average) when these data were available (Belgium, France, Spain, UK). In contrast, the daily number of patients currently hospitalised was used for Italy, and the daily number of new patients in intensive care units were used for Sweden. The sources of the hospitalisation data are summarised in Table S5 and the data points used for calibrations are presented in Figure 1 (main text).

| Country | Hospitalisation indicator | Source |
|---|---|---|
| Belgium | New hospital admissions | COVID-19 Belgium Epidemiological Situation dashboard [20] |
| France | New hospital admissions | data.gouv.fr platform (French Government) [21] |
| Italy | Current total of hospitalisations | National COVID-19 data repository [22] |
| Spain | New hospital admissions | Spanish Ministry of Health [23] |
| Sweden | New ICU admissions | Swedish Intensive Care Registry [24] |
| United Kingdom | New hospital admissions | COVID-19 Government dashboard [25] |

**Table S5. Summary of hospital data used to calibrate the models.**

We also used estimates from seroprevalence surveys for model calibration, assuming that observed seropositive proportions could be compared to the modelled proportions of ever-infected individuals. However, as it was demonstrated that antibody prevalence declines over time [26], we only used seroprevalence data as calibration target for the countries where a nationwide seroprevalence survey was conducted shortly after the first wave's notification peak and where data were reported publicly. This was the case for all countries except Italy. When multiple surveys were available, we used the results of the survey that was conducted the soonest after the first wave's notification peak. Table S6 presents the seroprevalence estimates used for calibration.

| Country | Population | Approach | Date | Result | Source |
|---|---|---|---|---|---|
| Belgium | Nationwide | Residual sera taken outside hospitals by diagnostic laboratories. | 20-26 Apr 2020 | 6.0% (5.1 - 7.1) | [26] |
| France | Nationwide | Residual sera from Biobanks. LuLISAs and PNT assays. | 6-12 Apr 2020 | 4.1% (3.3 5.0) | [27] |
| Spain | Nationwide | Fingerprick RDT, and CLIA on serum | 27 Apr - 11 May 2020 | 5.0% (4.7 - 5.4) | [28] |
| Sweden | 9 regions | Blood samples taken in outpatient care for a medical indication other than COVID-19 | 20 Apr - 26 May 2020 | 5.2% (3.8 - 6.9) | [29] |
| United Kingdom | Nationwide | Biobank participants, self-collected fingerprick sample tested with ELISA | 27 Apr - 3 May 2020 | 7.1% (6.7 - 7.5) | [30] |

**Table S6. Survey estimates of seroprevalence used for model calibration. The numbers in brackets represent 95% confidence intervals.**

#### 2.3 Likelihood calculation

Let $c_i$ denote the average daily number of new confirmed COVID-19 cases in a given country during week $i$, and $\hat{c_i}^\theta$ the associated predicted number according to the model when using the parameter set $\theta$. Similarly, let us denote $h_i$ as the average daily number of new COVID-19 hospitalisations (or average daily hospital occupancy for Italy, or average daily number of new ICU admissions for Sweden) during week $i$, and $\hat{h_i}^\theta$ the associated predicted number according to the model when using the parameter set $\theta$. Finally, let $d_i$ denote the average daily number of COVID-19 deaths during week $i$, and $\hat{d_i}^\theta$ the associated predicted number according to the model with parameter set $\theta$. For the countries where seroprevalence data were available for calibration (i.e. Belgium, France, Spain, Sweden, UK), let denote $s$ the observed proportion of seropositive individuals at the midpoint of the survey period indicated in Table S6 and $\hat{s}^\theta$ the proportion of recovered individuals predicted by the model for the same time point when using the parameter set $\theta$.

When seroprevalence data were not available (Italy), the likelihood was defined as follows:

$$\mathcal{L}(\theta) := \prod_i f(c_i|\,\hat{c_i}^\theta,\,\sigma_c) \times f(h_i|\,\hat{h_i}^\theta,\,\sigma_h) \times f(d_i|\,\hat{d_i}^\theta,\,\sigma_d),$$

where $f(.|\,\mu,\,\sigma)$ is the probability mass function of the normal distribution with mean $\mu$ and standard deviation $\sigma$.

For the five countries where seroprevalence was used as calibraiton target, the likelihood was defined as follows:

$$\mathcal{L}(\theta) := f(s|\,\hat{s}^\theta,\,\sigma_s) \times \prod_i f(c_i|\,\hat{c_i}^\theta,\,\sigma_c) \times f(h_i|\,\hat{h_i}^\theta,\,\sigma_h) \times f(d_i|\,\hat{d_i}^\theta,\,\sigma_d).$$

The parameters $\sigma_c$, $\sigma_h$, $\sigma_d$ and $\sigma_s$ were automatically estimated by the adaptive Metropolis algorithm.

#### 2.4 Parameters' posterior distributions

The posterior distributions of the parameters listed in Table S2 are presented in Figure S6 and the parameter traces after burn-in are shown in Figure S7.

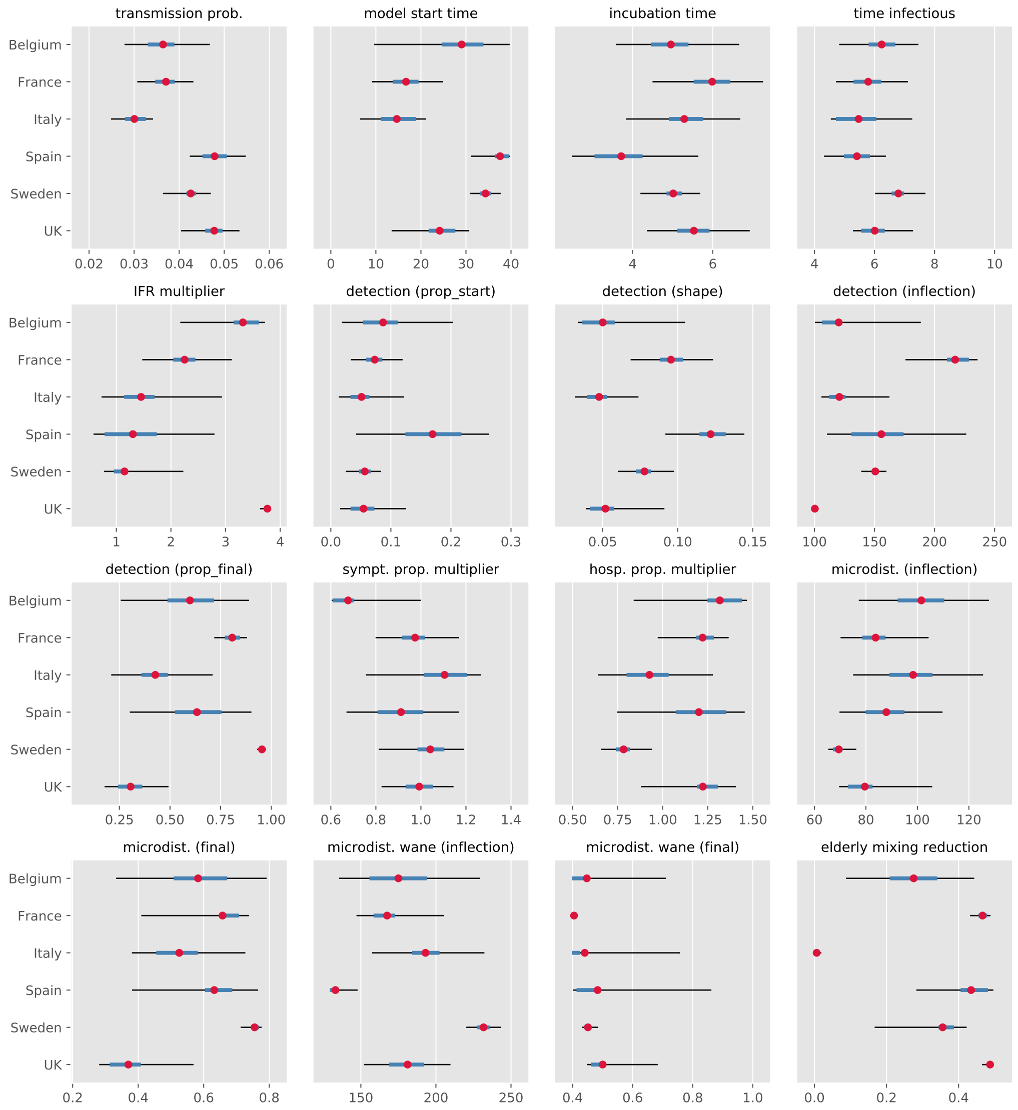

**Figure S6. Posterior estimates of model parameters.**
The mean estimates are represented with a red dot. The central 50% credible intervals are shown in blue and the central 95% credible interval are represented with black bars.

Using the posterior samples of the detection parameters, we computed estimated profiles of the time-variant proportion of symptomatic detected. The six countries' detection profiles are shown in Figure S8.

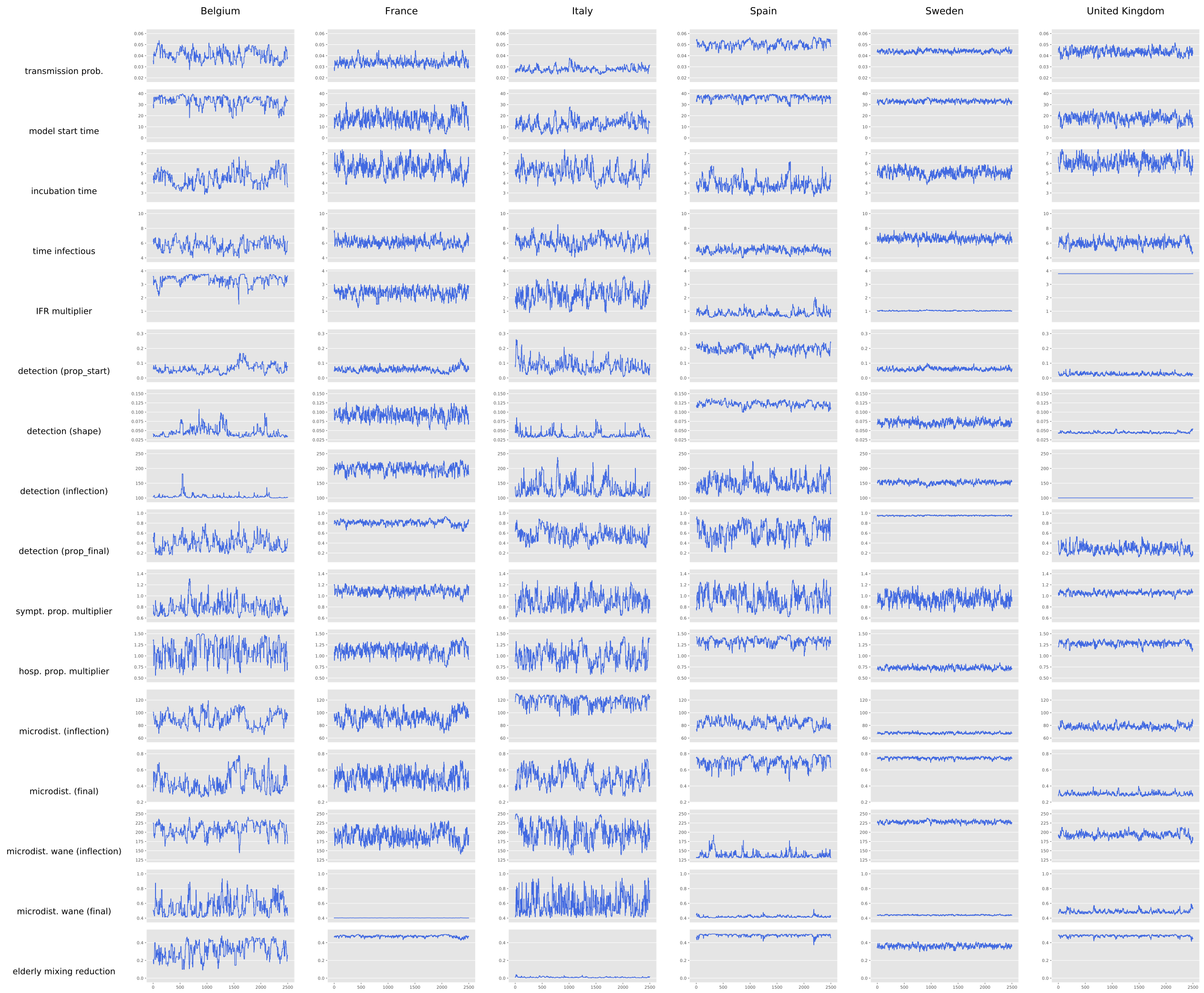

**Figure S7. Parameter traces.**
The 2,500 iterations obtained after burn-in are represented.

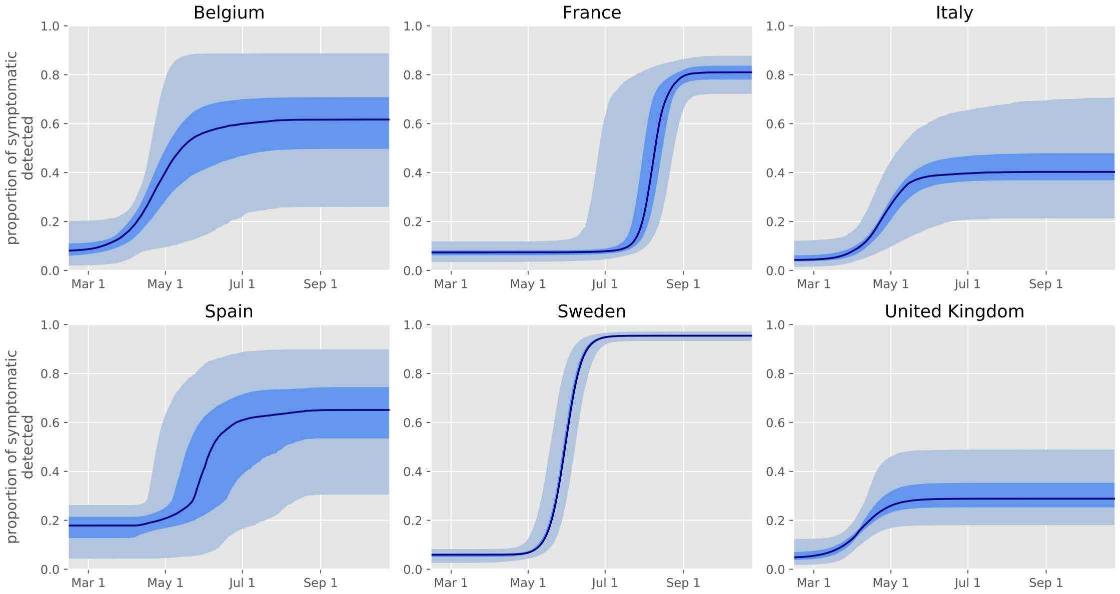

**Figure S8. Posterior estimates of the time-variant proportion of symptomatic persons detected.**
The figures present the median estimate (black line), central 50% credible interval (dark blue) and central 95% credible interval (light blue).

### 3 The three simulation phases

The model is used to simulate three successive phases that have different purposes:

1. Modelling the past (until 30 September 2020),
2. Mitigating the age-specific or location-specific social mixing (during 6 or 12 months from 1 October 2020),
3. Relaxing all restrictions and testing for herd immunity (until 31 December 2021).

Figure 1 (main text) illustrates these three phases, both in the presence and in absence of herd-immunity.

#### 3.1 Phase 1: Modelling the past

This phase aims to simulate the past SARS-CoV-2 epidemic accurately in order to capture the level of immunity acquired by August 2020. During this phase which ends on 30$^{\text{th}}$ September 2020, model parameters were automatically calibrated in order for the model predictions to match the numbers of COVID-19 cases, hospitalisations and deaths, as well as seroprevalence for all countries except Italy (Section 2). Our Bayesian approach to calibration also allowed us to account for uncertainty in the model inputs. This approach is described in Section 2.

#### 3.2 Phase 2: Mitigating the age-specific or location-specific social mixing

The second simulation phase begins on the 1$^{\text{st}}$ of October 2020 and lasts 6 or 12 months depending on the configuration considered. It is during this phase that the social mixing profile was varied and optimised.

**Optimisation performed by adjusting mixing by age**
In a first analysis, we aimed to find the optimal age-specific adjustments to the contact matrix. Under this scenario, the location-specific multipliers $s(t)$, $w(t)$ and $l(t)$ described in Equation 1 were all set back to 1, while mild micro-distancing was assumed to be maintained ($m_d = 0.9$, see Section 1.7.3). All the other parameter values remained the same as those used during Phase 1, including for the parameters that are automatically calibrated (Section 2). In addition, we used age-specific multipliers that were varied during optimisation in order to model various strategies of mitigation by age.

Let us denote $m_i \in [0.1, 1]$ a relative mixing multiplier associated with age group $i \in \{1, ..., 16\}$. During Phase 2, we apply the adjusted contact matrix $A$ defined by:

$$A[i, j] = m_i m_j C_0[i, j],$$

where $C_0$ is the original contact matrix provided by Prem *et al.* and described in Section 1.7.3. Note that during Phase 2, the matrix $A$ replaced the matrix $C$ in the equations presented in Section 1.8. We aimed to identify a combination $\{m_i, i \in \{1, ..., 16\}\}$ that:

1. leads to herd immunity by the end of Phase 2 and
2. minimises the mortality-related objective.

The mixing factors $m_i$ were constrained to stay over 10% in order to allow for a minimum level of mixing. This means that the opportunity of contacts could be reduced by no more than 90%, as compared to the pre-COVID-19 era.

Note that in the context of the optimisation by age, the potential reduction in the contact rates of the elderly inferred during calibration (Section 1.7.3) is not applied during Phase 2. Indeed, the related age-specific parameters are overridden by the optimised age-specific multipliers.

In Figure S9, we show the age-specific contributions in terms of total number of social contacts towards the elderly population. The contribution of age-group $j$ in terms of contacts towards the age-group $i$ was calculated as $C_0[i, j] \times \pi_j$, where $\pi_j$ is the population of age-group $j$. These quantities are presented to help understand the optimised mixing profiles presented in the main text.

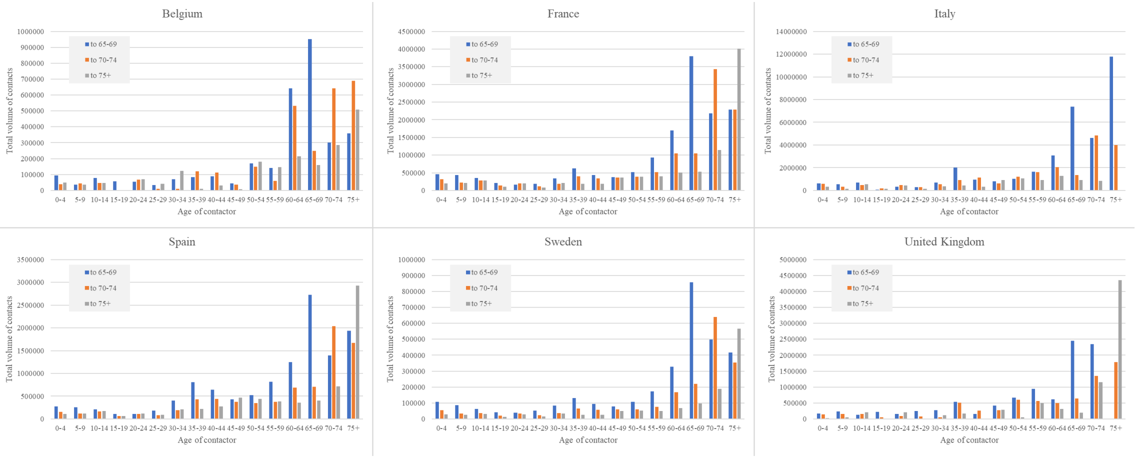

**Figure S9. Age-specific contributions in terms of total number of contacts towards the elderly population.**

##### Optimisation performed by adjusting mixing by location

In a separate exercise, we performed optimisation of social mitigation by location. Namely, we aimed to identify the optimal combinations of contact rate reductions in the three following locations: schools, workplaces and places other than schools, workplaces and homes. The rates of contacts occurring between household members were assumed to remain unchanged during this exercise and mild micro-distancing was assumed to be maintained ($m_d = 0.9$, see Section 1.7.3) during the optimisation phase. We aimed to achieve the same goals as with social mitigation by age. Using the notation introduced in Equation 1, the decision variables of the optimisation problem become the location-specific multipliers $s(t)$, $w(t)$ and $l(t)$ that we consider constant during Phase 2. Note that

##### The mortality-related objectives to minimise

We considered two different mortality-related objectives separately in this study. First, the optimisation exercise aimed to minimise the total number of deaths occurring during Phases 2 and 3. Then, we repeated all analyses by minimising the total number of life-years lost during the same period. The number of life-years lost was estimated by summing the expected number of remaining years that individuals would have lived if they had not died from COVID-19. This process was informed by the country-specific life-expectancy values by age reported by the United Nations.

##### 3.3 Phase 3: Relaxing all restrictions and testing herd immunity

In this analysis, we were only interested in the strategies of mitigation that result in herd immunity by the end of Phase 2. We used a simulation-based approach to test whether herd immunity was reached at that time. To do this, we set all the age-specific mixing multipliers $m_i$ and the location-specific multipliers $s$, $w$ and $l$ back to 1 at the start of Phase 3 in order to simulate an unmitigated epidemic. We assumed that herd immunity was reached by the end of Phase 2 if and only if the number of new diseased individuals was consistently found to decrease during Phase 3.

During Phase 3, we still applied the same level of mild micro-distancing as assumed during Phase 2. This was done to capture the likely long-term changes in individuals' behaviours and the preventive measures that may still be taken in the future, as public awareness of the modes of transmission of SARS-CoV-2 has now increased. Similarly, the inferred contact reduction in the elderly after the first waves due to age-specific behavioural changes (Section 1.7.3) was also applied during Phase 3.

#### 4 Optimisation of the age-specific social mixing

##### 4.1 Problem description

**Optimisation by age**

For each country and for each configuration, we aimed to identify optimal combinations of the age-specific multipliers $m_i$ described in Section 3. Let us denote $\Phi = \{m_1, \ldots, m_{16}\} \in [0.1, 1]^{16}$ a combination of these age-specific multipliers. Let also $\Psi(\Phi)$ be the associated mortality-related objective (either total numbers of deaths or life-years lost during Phases 2 and 3), and $H(\Phi)$ a binary variable indicating whether herd immunity has been achieved by the end of Phase 2 ($H(\Phi) = 1$ if herd immunity, 0 otherwise). We aimed to find $\Phi^*$ such that:

$$\Phi^* = \arg\min_{\Phi} \Psi(\Phi) \; , \tag{4}$$

$$\text{subject to: } H(\Phi) = 1 \; . \tag{5}$$

**Optimisation by location**

The optimisation by location was defined in the same way as the optimisation by age except that the decision variables were the location-specific multipliers $s$, $w$ and $l$ described in Section 1.7.3 instead of the age-specific multipliers $m_i$.

##### 4.2 Technical description of the optimisation algorithm

A total of 48 optimisations searches were performed, as we considered 6 countries, two different Phase 2 durations (6 and 12 months), two different objectives to minimise (deaths and YLLs), and two optimisation modes (by age and by location). The optimisation exercises were treated independently using a parallel mono-objective Evolutionary Algorithm (EA) [31]. In an EA, a population of combinations is evolved by selection and reproduction of promising combinations already simulated (parents). The new combinations (children) generated by the reproduction operators are then simulated and replace the less promising combinations into the population.

The constraint of achieving herd immunity presented in Equation 5 was integrated into the mortality-related objective by setting the objective value to 8,000,000,000. This number is overwhelmingly greater than both the total number of possible deaths and the total number of possible life-years lost for all the countries considered in this study.

The parallel EA is described in Algorithm 1. The population of $N$ combinations was initialized using Latin Hypercube Sampling and was then simulated in parallel (line 1 and 2). Within the generational loop (line 4 to 11), a population of parents was created by selecting $N$ combinations from the population. The tournament selection of size 2 (line 5) repeatedly sampled 2 combinations with replacement from the population and retained the best one as parent. Parent characteristics were mixed by the SBX crossover operator [32] to generate the population of children (line 6). Children are next slightly modified by the polynomial mutation operator [32] and simulated in parallel (line 7 and 8). The best combinations from the population and the children were retained to form the new population (line 9).

**Algorithm 1** Parallel Evolutionary Algorithm

**Input**
$N$: population size
*budget*: budget for the search

1: $\mathcal{P} \leftarrow$ LHS_sampling($N$) ▷ initial population
2: parallel_simulation($\mathcal{P}$)
3: $(\boldsymbol{x}_{min}, y_{min}) \leftarrow$ get_best_cost($\mathcal{P}$)
4: **while** $budget \neq 0$ **do**
5: $\quad \mathcal{P}_p \leftarrow$ tournament(2, $\mathcal{P}$) ▷ population of parents
6: $\quad \mathcal{P}_c \leftarrow$ SBX_crossover($\mathcal{P}_p$) ▷ population of children
7: $\quad \mathcal{P}_c \leftarrow$ polynomial_mutation($\mathcal{P}_c$)
8: $\quad$ parallel_simulation($\mathcal{P}_c$)
9: $\quad \mathcal{P} \leftarrow$ elitist_replacement($\mathcal{P}$, $\mathcal{P}_c$, $N$)
10: $\quad (\boldsymbol{x}_{min}, y_{min}) \leftarrow$ get_best_cost($\mathcal{P}$)
11: **end while**
12: **return** $\boldsymbol{x}_{min}, y_{min}$

Simulations were performed in parallel as they were the most computationally expensive part of the process (6 seconds on 1 computational core). The budget for the search was set to 12 hours on 1 computational node made of 18 Intel Xeon Gold 5220 cores proceeding from Grid5000, a large-scale testbed with a focus on parallel and distributed computing [33]. The population size was set to 126 in order to follow the general guidance given in [31] and to minimise the number of idle cores when simulating a population.

#### 4.3 Sensitivity analysis applying perturbations on the optimal plans

Figures 2 and 3 (main text) present the optimal plans obtained after minimisation by age and by location, respectively. We also measured how variations in the optimised decision variable would affect the objective functions (numbers of deaths and YLLs). To do this, we set a maximum allowed number of additional deaths or YLLs compared the optimum obtained under each configuration ($\Delta obj$). We then determined the alterations of the mixing factors that would:

1. cause an increase of $\Delta obj$ in the objective function and
2. maintain the condition of herd immunity.

The thin black bars on Figures 2 and 3 represent the greatest perturbations allowed. These perturbations were applied to one mixing factor at a time.

### 5 Additional results

#### 5.1 Age-specific infected proportions

While the age-specific seroprevalence data were not used for model calibration, we compare these estimates with the age-specific proportions of recovered individuals obtained from our models for validation purposes.

The following five figures show these comparisons for Belgium, France, Spain, Sweden and the UK, respectively.

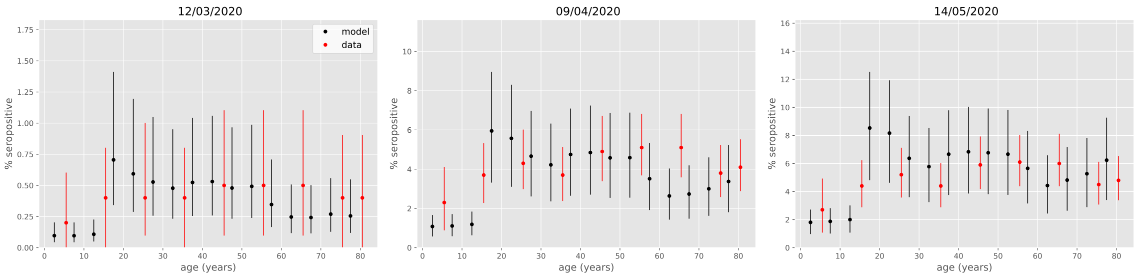

**Figure S10. Age-specific proporitons of recovered individuals against seroprevalence data (France)**

Serosurvey data are shown in red and model estimates are represented in black, both at the midpoints of the reported age groups. The vertical bars show the 95% credible intervals. Serosurvey data were extracted from Levin *et al.* [5].

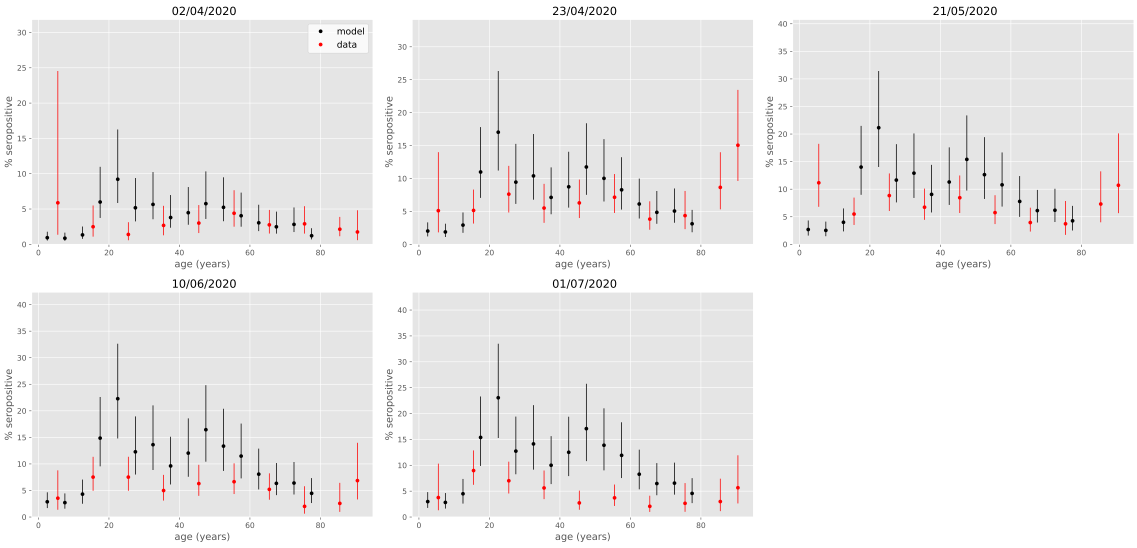

**Figure S11. Age-specific proporitons of recovered individuals against seroprevalence data (Belgium)**

Serosurvey data are shown in red and model estimates are represented in black, both at the midpoints of the reported age groups. The vertical bars show the 95% credible intervals. Serosurvey data were extracted from Herzog *et al.* [26].

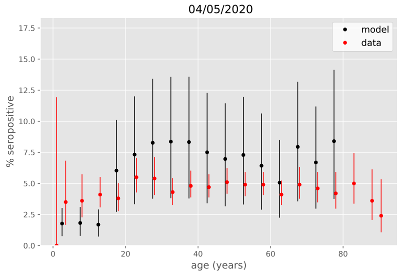

**Figure S12. Age-specific proporitons of recovered individuals against seroprevalence data (Spain)**
Serosurvey data are shown in red and model estimates are represented in black, both at the midpoints of the reported age groups. The vertical bars show the 95% credible intervals. Serosurvey data were extracted from Pollan *et al.* [28].

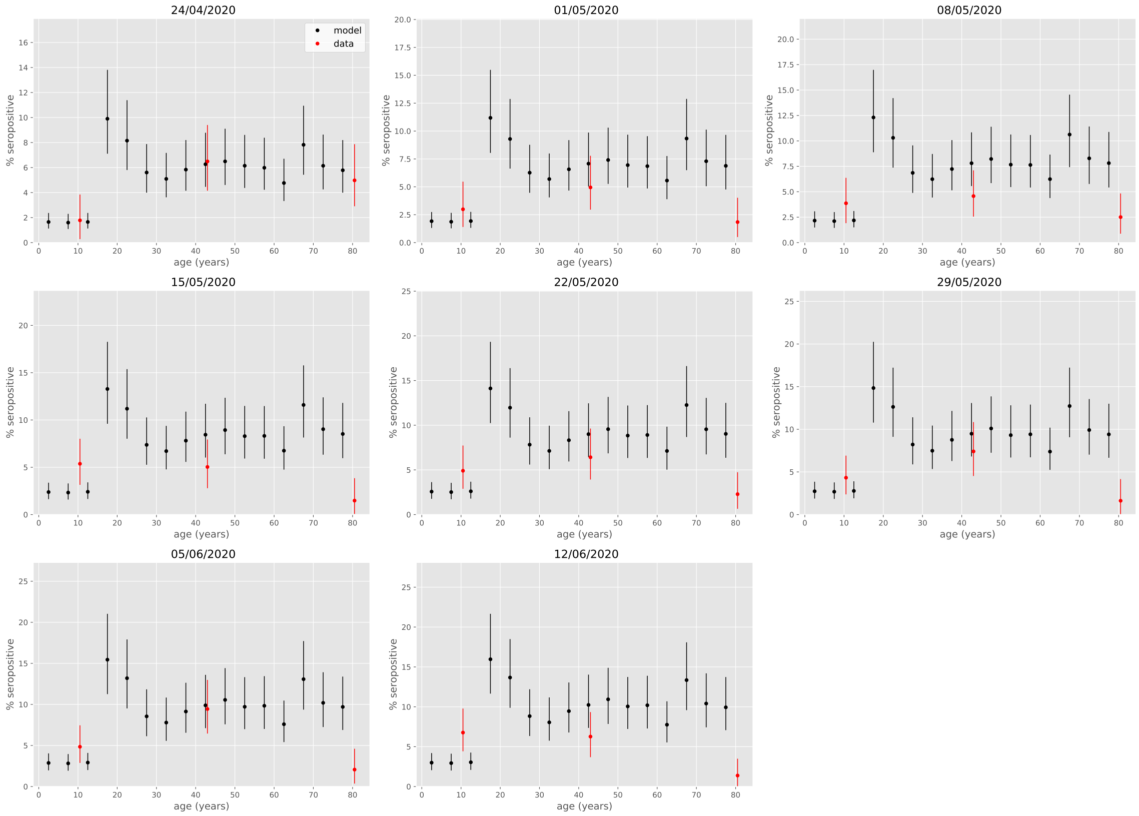

**Figure S13. Age-specific proporitons of recovered individuals against seroprevalence data (Sweden)**
Serosurvey data are shown in red and model estimates are represented in black, both at the midpoints of the reported age groups. The vertical bars show the 95% credible intervals. Serosurvey data were extracted from an interim report of the Swedish Public Health Agency [29].

#### 5.2 Effect of increased mixing on deaths and YLLs

We explored the effect of increasing mixing as compared to the optimal mitigation plans obtained using contact mitigation by age. That is, we applied a minimum bound $b$ to the age-specific mixing factors such that the modified mixing profile could be described as $\{max(m_i^*, b)\}_{i \in \{1,\ldots,16\}}$, where $\{m_i^*\}_{i \in \{1,\ldots,16\}}$ represents the optimal (reference) solution originally obtained in the main analysis using $b = 0.1$. Figure S15 shows the results of

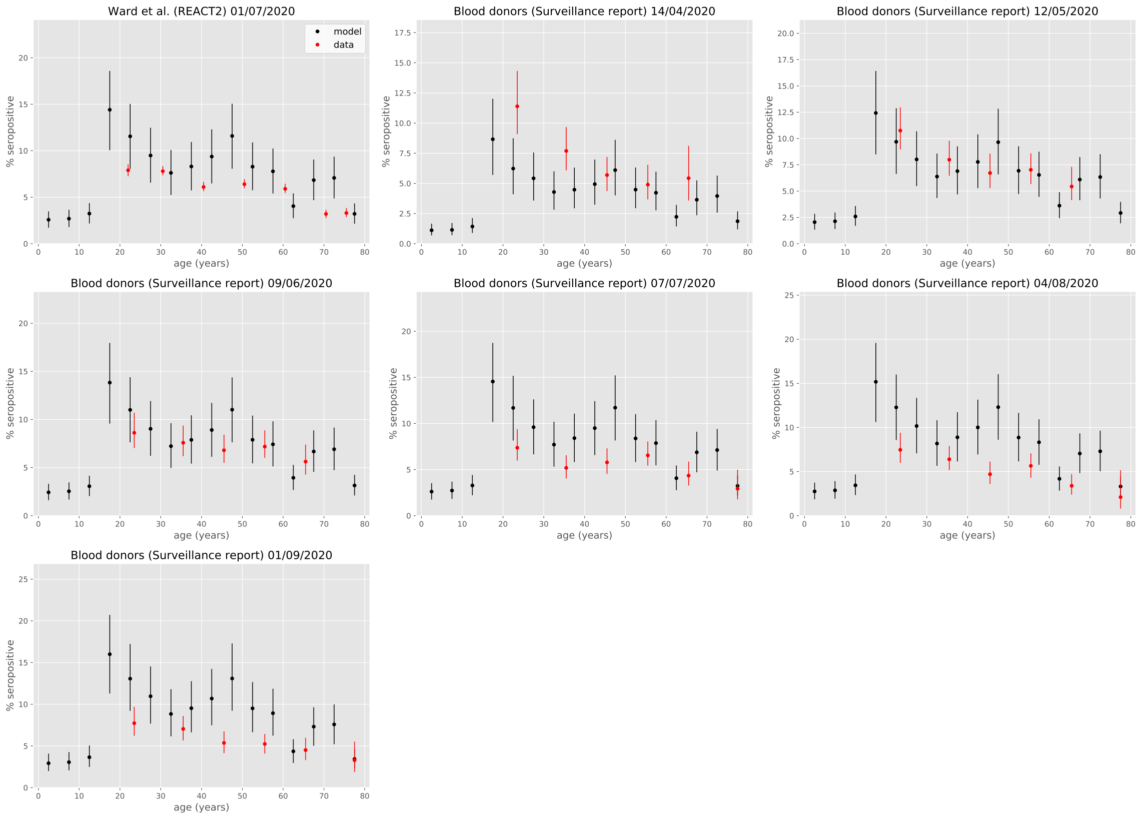

**Figure S14. Age-specific proporitons of recovered individuals against seroprevalence data (UK)**

Serosurvey data are shown in red and model estimates are represented in black, both at the midpoints of the reported age groups. The vertical bars show the 95% credible intervals. Serosurvey data were extracted from Ward *et al.* [34] and The Public Health England weekly surveillance report (week 40) .

this analysis in terms of predicted total numbers of COVID-19-related deaths and YLLs occurring after 1 October 2020 when considering $0.1 \leq b \leq .5$. Note that no additional optimisation was run for this exercise such that the predictions presented here were not optimal solutions. Recovered individuals were assumed to have persistent immunity in this analysis.

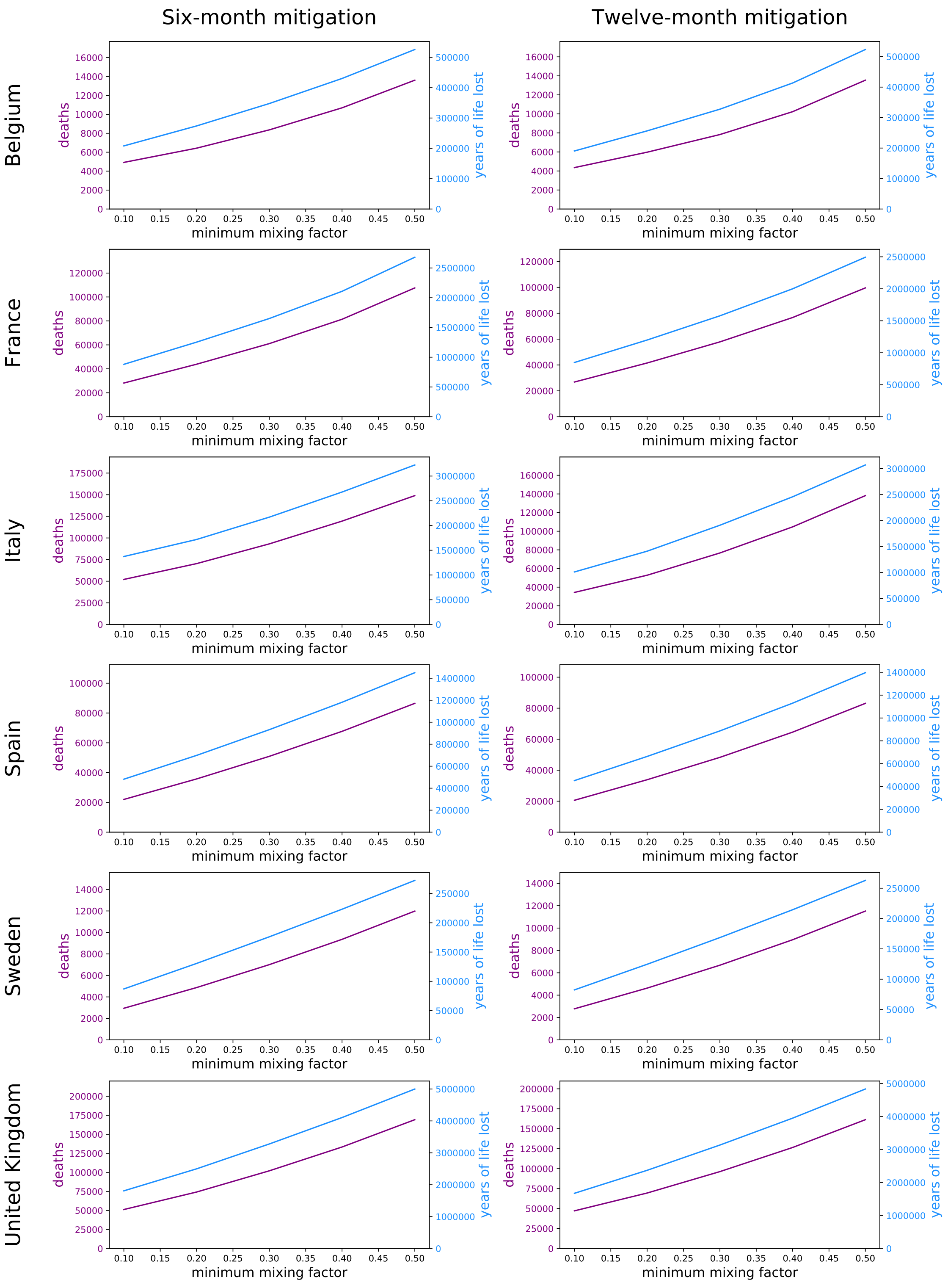

**Figure S15. Impact of increasing mixing compared to the optimal strategies.** The minimum mixing factor is the variable $b$ described above. The purple line represents the total number of COVID-19-related deaths occurring after 1 Oct 2020 using the reference solution of the optimisation by age minimising deaths. The blue line represents the total number of COVID-19-related YLLs occurring after 1 Oct 2020 using the reference solution of the optimisation by age minimising YLLs.

#### 5.3 Contact matrices resulting from the optimisation

The main text presents the optimisation results in terms of age-specific or location-specific mixing variables. Here we present the mixing matrices resulting from the optimised mixing factors. Figures S16 and S17 present the mixing matrices obtained under the different configurations when running the optimisation processes by ages and by location, respectively.

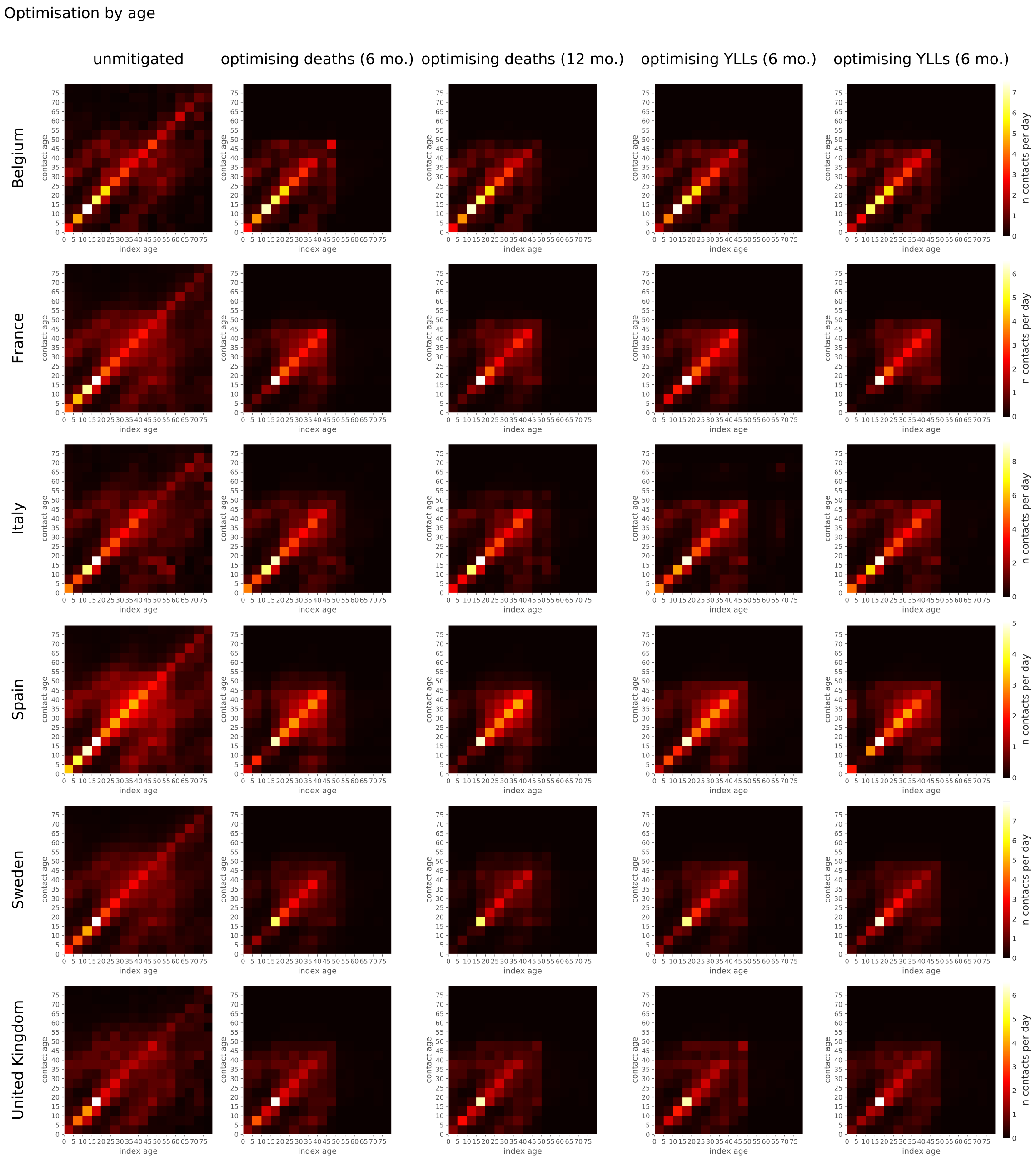

**Figure S16. Age-specific contact matrices obtained from the optimisations by age.**

Optimisation by location

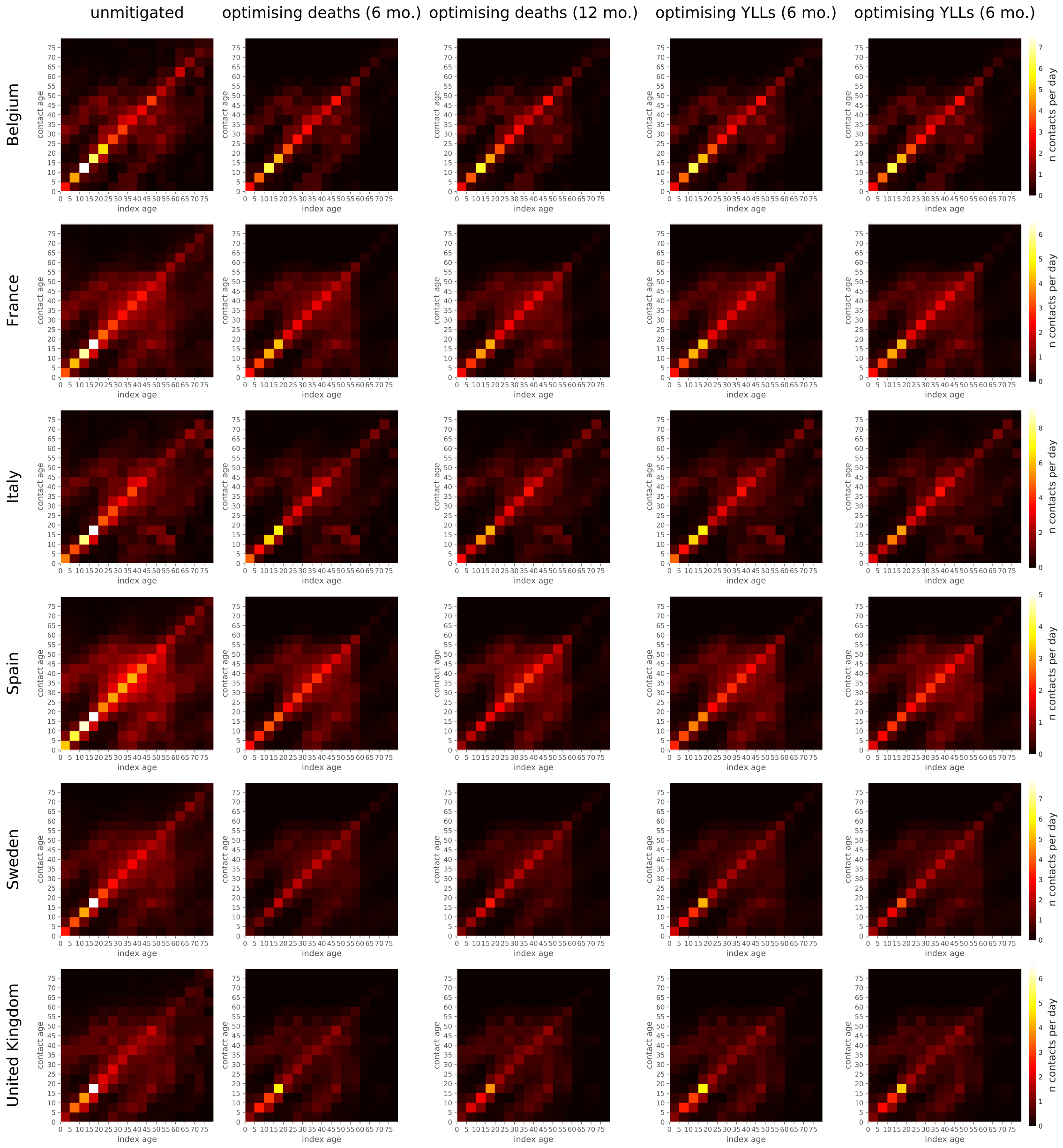

**Figure S17. Age-specific contact matrices obtained from the optimisations by location.**

#### 5.4 Estimates of proportions of recovered individuals under optimised scenarios

Figures S18 and S19 present the predicted proportions of recovered individuals over time under the optimised scenarios of mitigation by age and by location, respectively. Recovered individuals were assumed to have persistent immunity in these analysis.

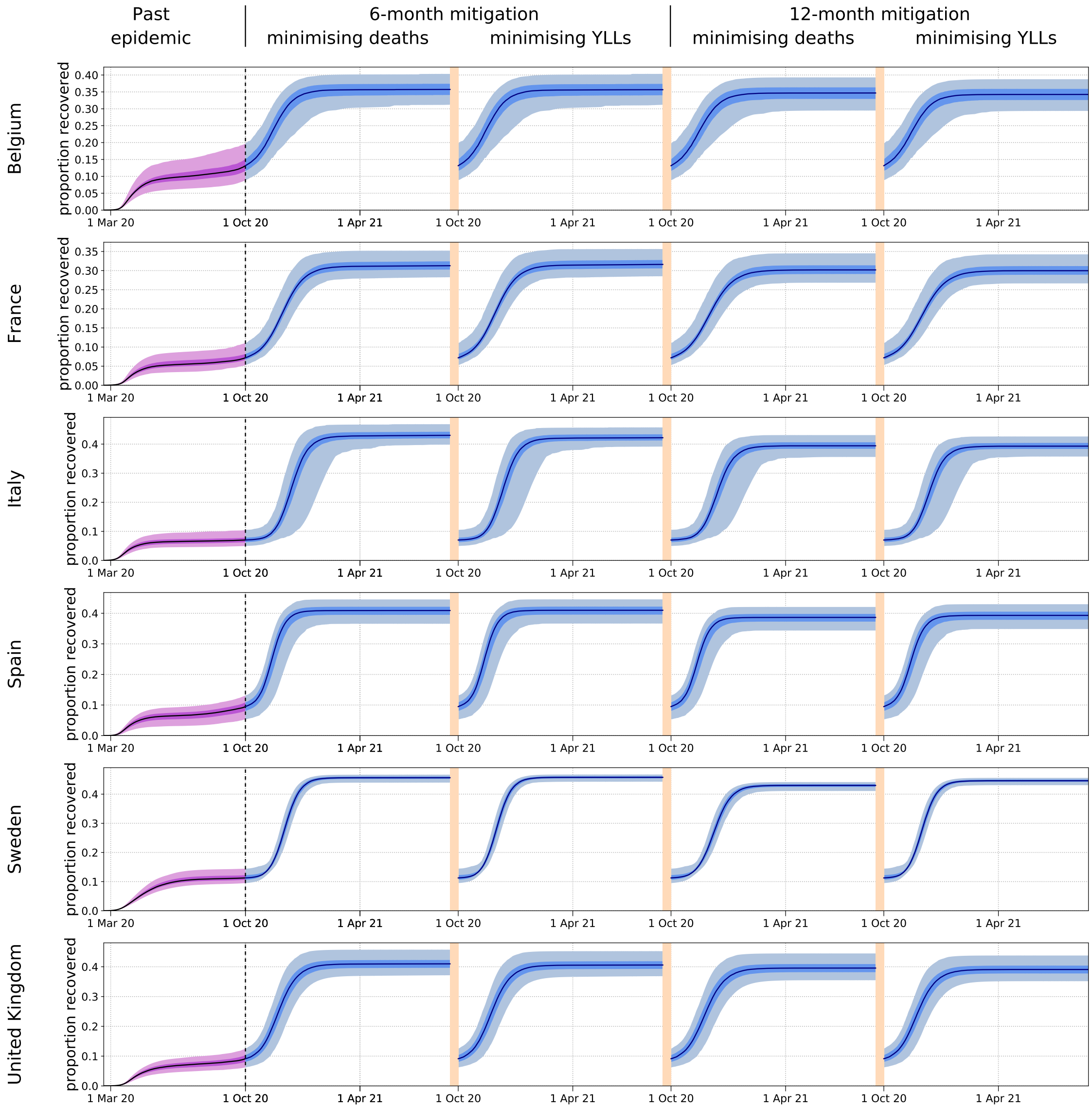

**Figure S18. Posterior estimates of proportions of recovered individuals over time (optimisation by age).**
The first waves (past epidemics) are represented in purple while the predictions of the future epidemics are represented in blue. The future epidemics are those associated with the four different optimisation configurations: six- or 12-month mitigation minimising total number of deaths or years of life lost (YLLs). The light shades show the central 95% credible intervals, the dark shades show the central 50% credible intervals and the solid lines represent the median estimates.

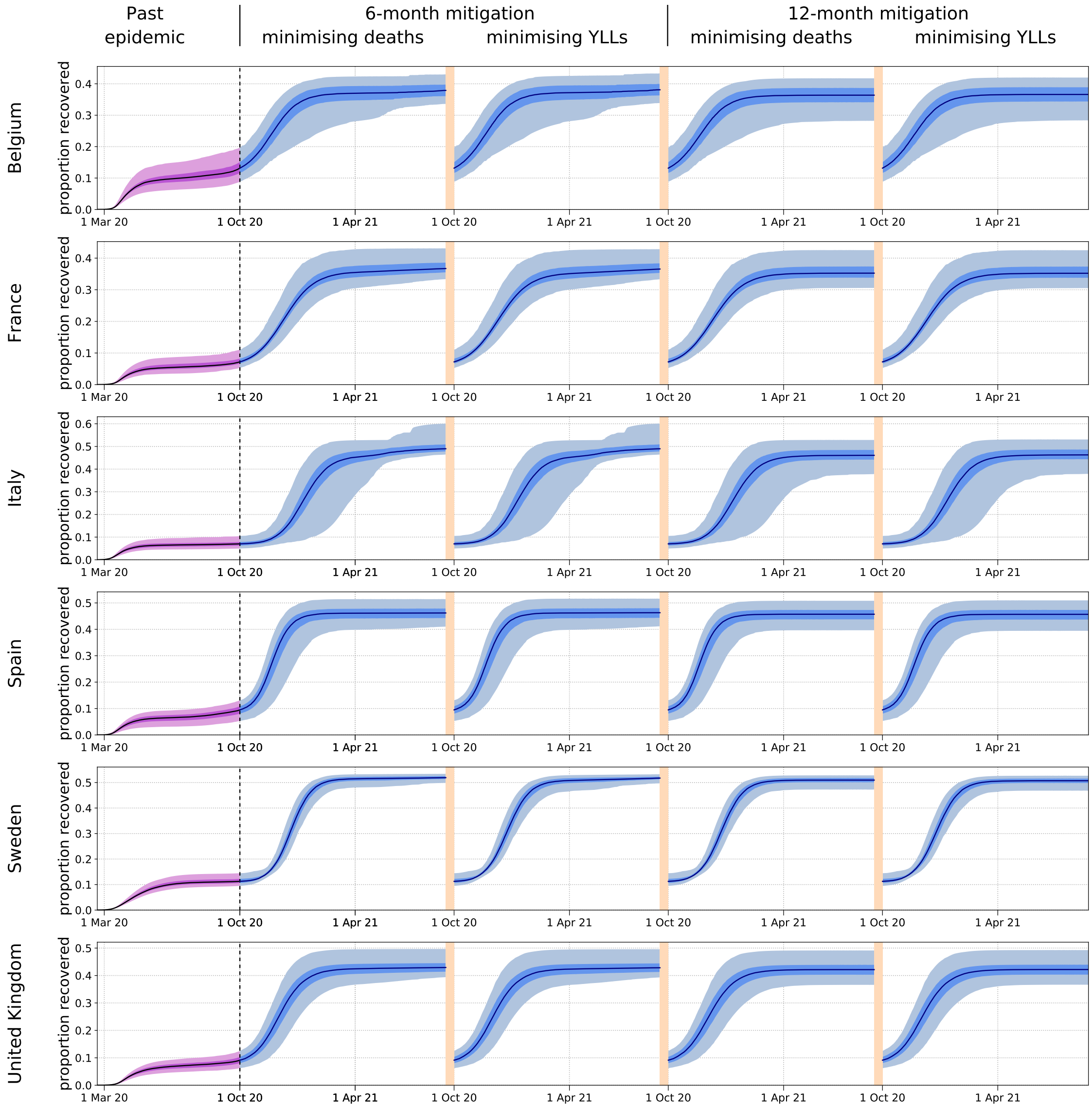

**Figure S19. Posterior estimates of proportions of recovered individuals over time (optimisation by location).**
The first waves (past epidemics) are represented in purple while the predictions of the future epidemics are represented in blue. The future epidemics are those associated with the four different optimisation configurations: six- or 12-month mitigation minimising total number of deaths or years of life lost (YLLs). The light shades show the central 95% credible intervals, the dark shades show the central 50% credible intervals and the solid lines represent the median estimates.

Figures S20 and S21 show the age-specific proportions of recovered individuals after the optimised mitigation phase when optimising by age and by location, respectively. Since the optimisation was constrained by the fact that herd immunity had to be reached by the end of the mitigation phase, these proportions could also be interpreted as age-specific vaccine coverage that would lead to herd immunity using a 100% vaccine.

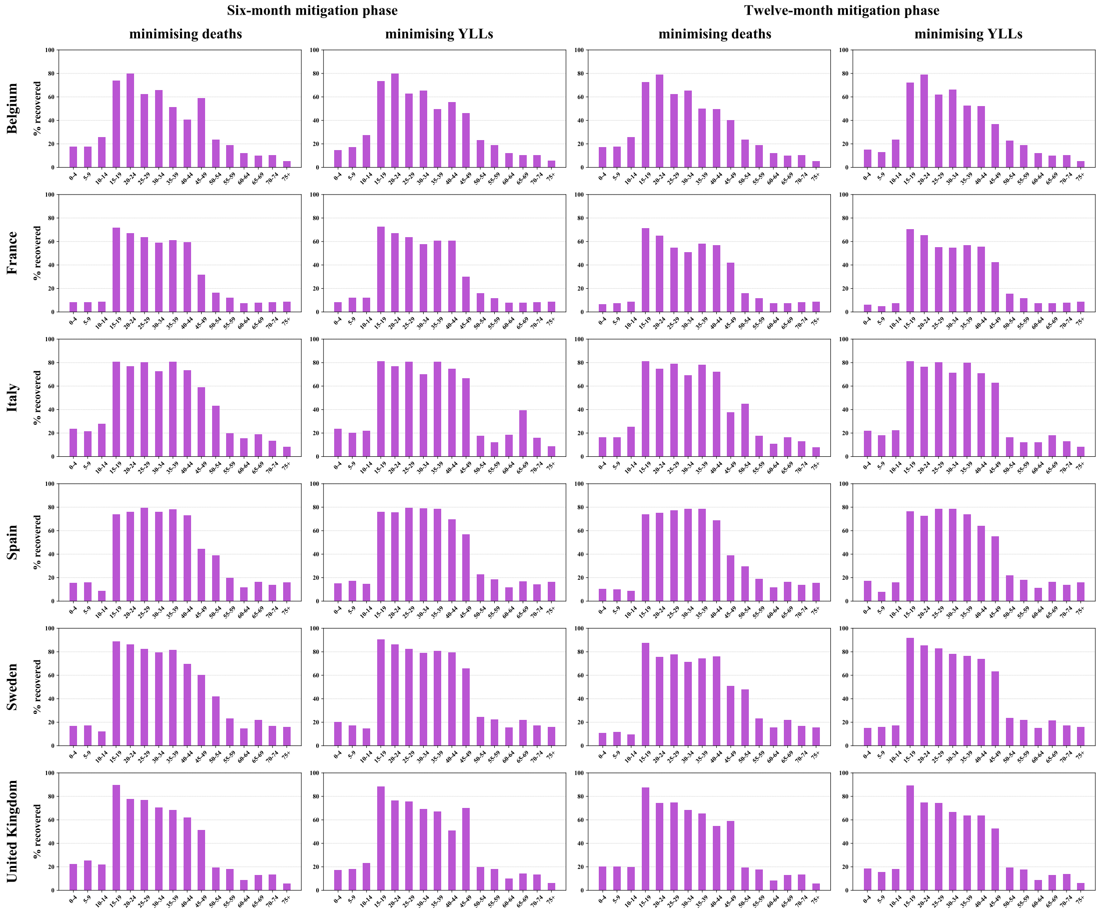

**Figure S20. Age-specific proportions of recovered individuals at the end of Phase 2 (optimisation by age).**

Recovered individuals assumed to have persistent immunity. Simulations based on the maximum-likelihood parameter sets. YLLs: years of life lost.

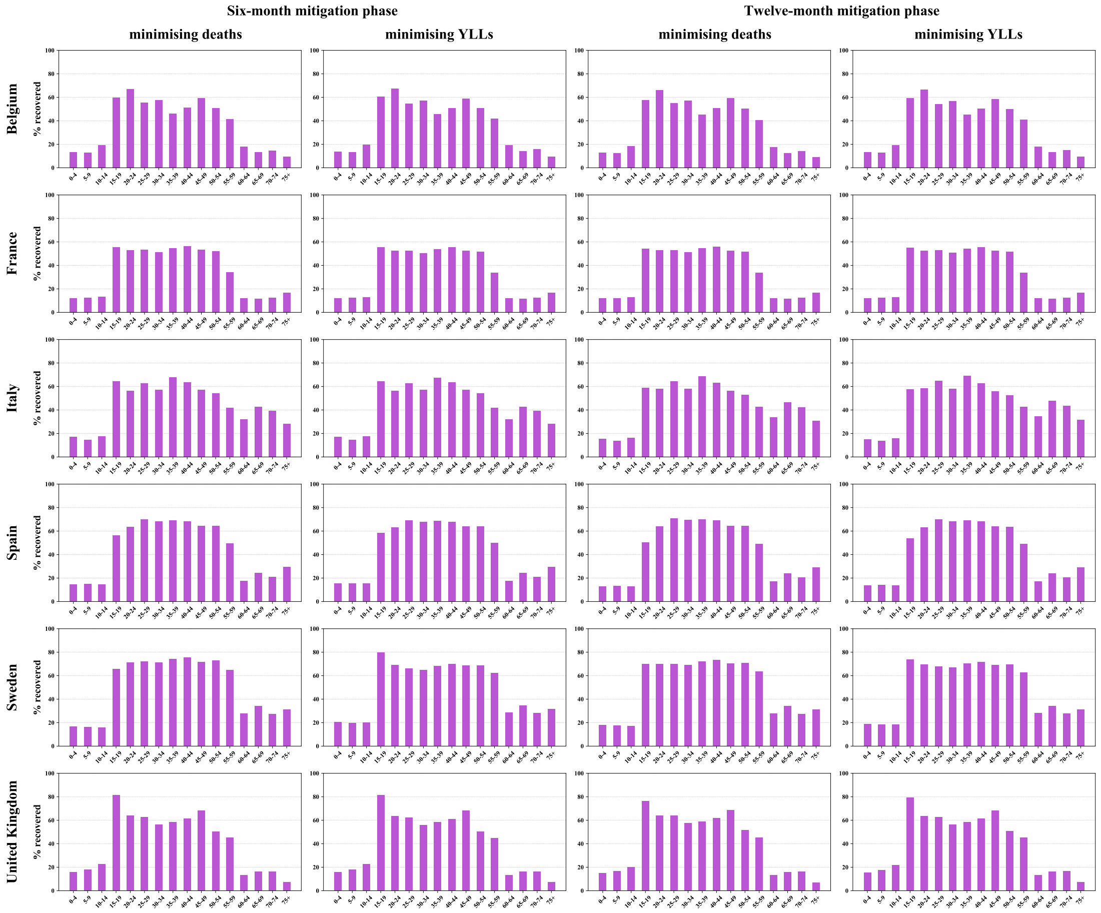

**Figure S21. Age-specific proportions of recovered individuals at the end of Phase 2 (optimisation by location).**

Recovered individuals assumed to have persistent immunity. Simulations based on the maximum-likelihood parameter sets. YLLs: years of life lost.

#### 5.5 Predicted numbers of deaths and YLLs per capita

Table S7 presents the predicted numbers of COVID-19-related deaths and YLLs per capita associated with the optimised and unmitigated scenarios. Recovered individuals were assumed to have persistent immunity in these analysis.

| Country | Optimisation mode | Mitigation phase | Deaths before 1 Oct 2020 (per million) | | Deaths from 1 Oct 2020 (per million) | | | YLLs before 1 Oct 2020 (per 10,000) | YLLs from 1 Oct 2020 (per 10,000) | | |
|---|---|---|---|---|---|---|---|---|---|---|---|
| | | | Model prediction | WHO report | Unmitigated | Optimised | | Model prediction | Unmitigated | Optimised | |
| | | | | | | Minimising deaths | Minimising YLLs | | | Minimising deaths | Minimising YLLs |
| **Belgium** | by age | 6 mo. | 752 (519-1090) | 880 | 4317 (2929-4658) | 418 (314-480) | 424 (315-486) | 274 (181-402) | 1292 (936-1357) | 182 (137-213) | 179 (135-211) |
| | | 12 mo. | | | | 371 (208-436) | 376 (209-441) | | | 167 (95-199) | 163 (93-195) |
| | by location | 6 mo. | | | | 1043 (840-1401) | 1104 (888-1485) | | | 520 (405-603) | 508 (394-589) |
| | | 12 mo. | | | | 952 (478-1080) | 996 (498-1132) | | | 485 (256-555) | 478 (252-546) |
| **France** | by age | 6 mo. | 584 (373-808) | 485 | 5079 (3756-6321) | 428 (326-580) | 430 (329-582) | 120 (69-181) | 1059 (746-1356) | 136 (91-189) | 134 (90-187) |
| | | 12 mo. | | | | 406 (288-565) | 411 (292-573) | | | 130 (84-189) | 127 (82-184) |
| | by location | 6 mo. | | | | 1315 (1054-1803) | 1319 (1065-1805) | | | 422 (294-590) | 421 (295-586) |
| | | 12 mo. | | | | 1213 (871-1766) | 1219 (875-1773) | | | 390 (262-576) | 389 (261-574) |
| **Italy** | by age | 6 mo. | 616 (458-900) | 593 | 6482 (5350-7475) | 744 (524-1069) | 913 (637-1215) | 114 (72-193) | 1213 (792-1465) | 294 (171-420) | 239 (141-334) |
| | | 12 mo. | | | | 597 (431-735) | 599 (415-753) | | | 262 (151-355) | 175 (103-234) |
| | by location | 6 mo. | | | | 3302 (2709-4886) | 3304 (2710-4880) | | | 690 (465-1056) | 690 (465-1057) |
| | | 12 mo. | | | | 3075 (2225-3871) | 3172 (2294-3989) | | | 656 (420-830) | 646 (413-819) |
| **Spain** | by age | 6 mo. | 712 (325-1027) | 692 | 4970 (3507-7102) | 482 (357-767) | 483 (358-784) | 99 (45-187) | 742 (511-1526) | 129 (93-294) | 107 (78-236) |
| | | 12 mo. | | | | 443 (329-704) | 450 (335-706) | | | 109 (79-243) | 100 (73-219) |
| | by location | 6 mo. | | | | 1464 (1075-2145) | 1473 (1081-2163) | | | 304 (216-670) | 303 (216-668) |
| | | 12 mo. | | | | 1420 (1055-2031) | 1425 (1057-2034) | | | 296 (214-648) | 295 (213-646) |
| **Sweden** | by age | 6 mo. | 574 (486-662) | 584 | 3105 (2347-3692) | 302 (205-388) | 304 (203-391) | 96 (77-115) | 557 (400-690) | 107 (69-142) | 88 (57-116) |
| | | 12 mo. | | | | 285 (192-368) | 295 (193-385) | 96 (77-115) | | 111 (69-147) | 83 (53-110) |
| | by location | 6 mo. | | | | 1068 (868-1260) | 1096 (924-1287) | 96 (77-115) | | 278 (205-349) | 274 (210-342) |
| | | 12 mo. | | | | 1038 (733-1248) | 1041 (731-1253) | 96 (77-115) | | 269 (178-342) | 266 (175-339) |
| **United Kingdom** | by age | 6 mo. | 636 (449-833) | 620 | 6921 (6014-7857) | 704 (599-848) | 772 (659-925) | 192 (133-262) | 1682 (1569-1818) | 273 (238-324) | 259 (223-308) |
| | | 12 mo. | | | | 685 (564-839) | 716 (590-874) | 192 (133-262) | | 255 (213-307) | 252 (210-305) |
| | by location | 6 mo. | | | | 1341 (1226-1679) | 1346 (1229-1682) | 192 (133-262) | | 639 (594-769) | 638 (592-767) |
| | | 12 mo. | | | | 1249 (1055-1627) | 1269 (1071-1655) | 192 (133-262) | | 620 (537-769) | 616 (533-766) |

**Table S7. Predicted numbers of deaths and years of life lost per capita.** Optimisations realised under the assumption of persistent immunity. Numbers are presented per million population for deaths and per 10,000 population for YLLs as median and central 95% credible intervals. YLLs: Years of life lost.

#### 5.6 Hospital occupancy with optimised mitigation by location

Figure S22 presents the predicted daily number of beds occupied by COVID-19 patients over time under the optimised scenarios of mitigation by location. Recovered individuals were assumed to have persistent immunity in this analysis.

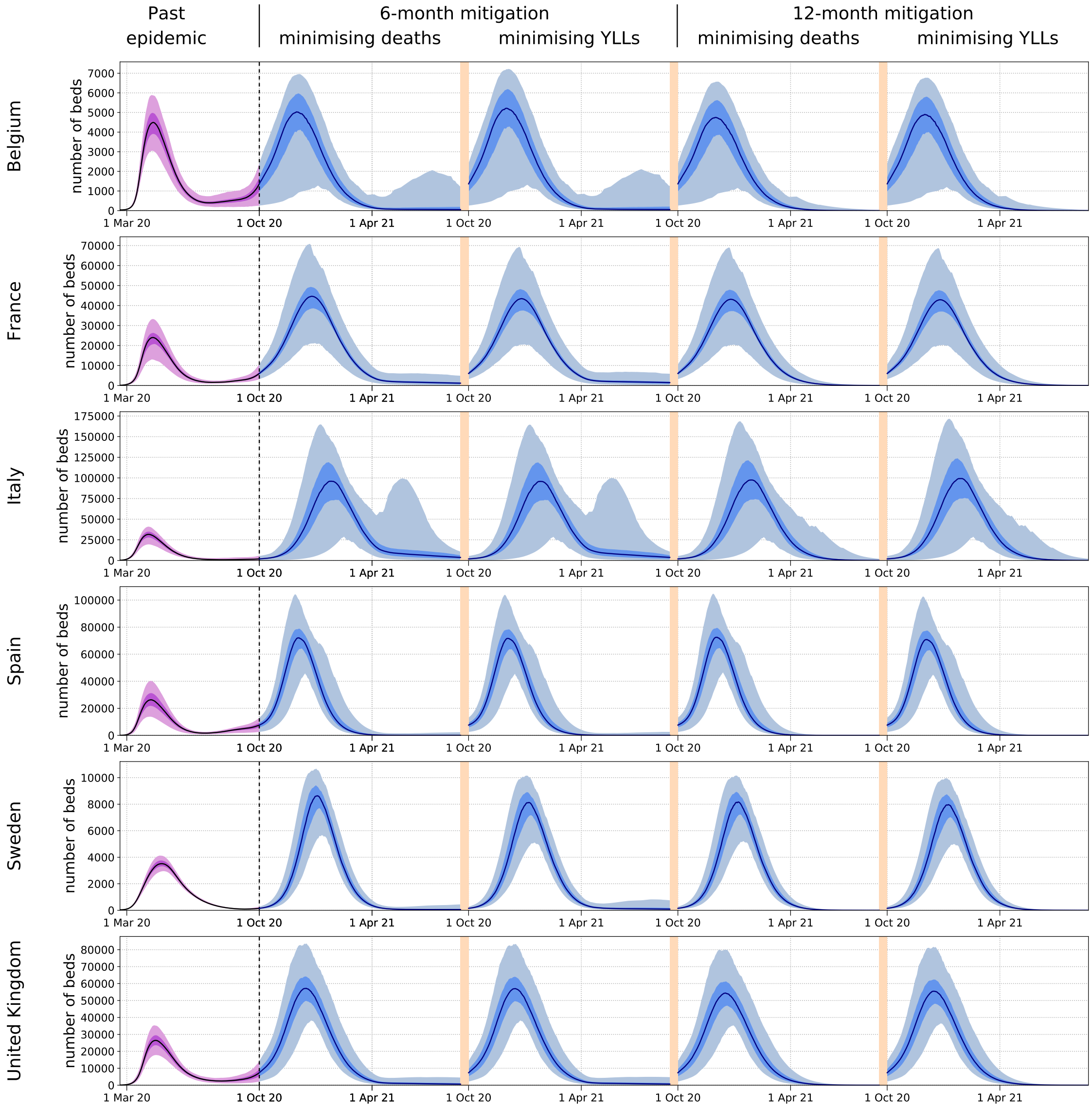

**Figure S22. Hospital occupancy with mitigation optimised by location (assuming persistent immunity).**
The first waves (past epidemics) are represented in purple while the predictions of the future epidemics are represented in blue. The future epidemics are those associated with the four different optimisation configurations: six- or 12-month mitigation minimising total number of deaths or years of life lost (YLLs). The light shades show the central 95% credible intervals, the dark shades show the central 50% credible intervals and the solid lines represent the median estimates.

#### 5.7 Age-specific profiles of disease indicators over time

The following figures are the equivalent of Figure 3 (main text) under different configurations of optimisation. The configurations are indicated at the top of each figure and describe the type of mitigation (by age or by location), the duration of the mitigation phase (6 or 12 months) and the minimised indicator (deaths or YLLs).

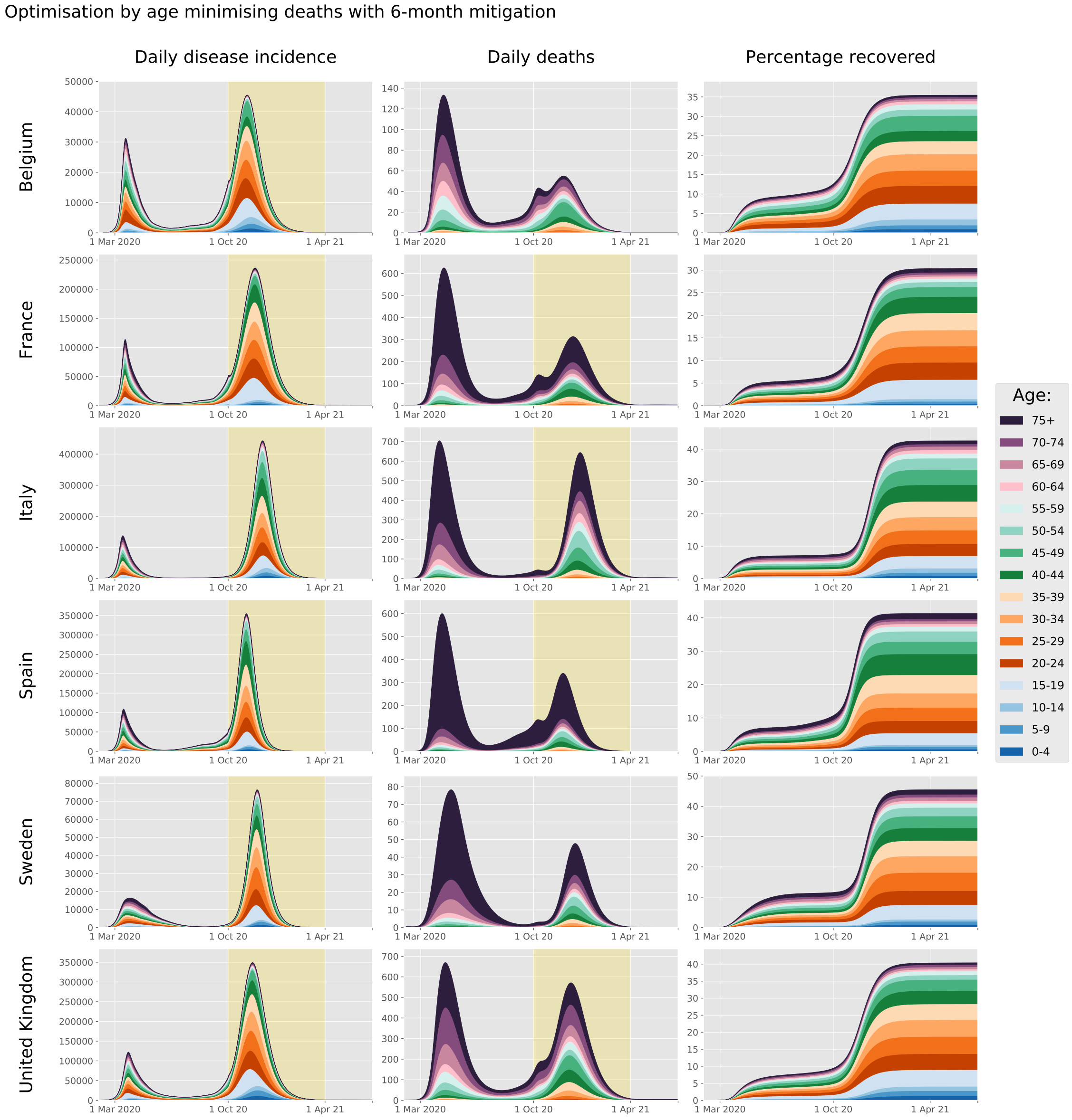

**Figure S23. Age-specific profile of disease incidence, COVID-19-related deaths and proportion recovered over time**
The yellow background indicates the intervention phase during which age-specific contacts were optimised. These projections were produced assuming that recovered individuals have persistent immunity against SARS-CoV-2 reinfection and using the maximum a posteriori estimates.

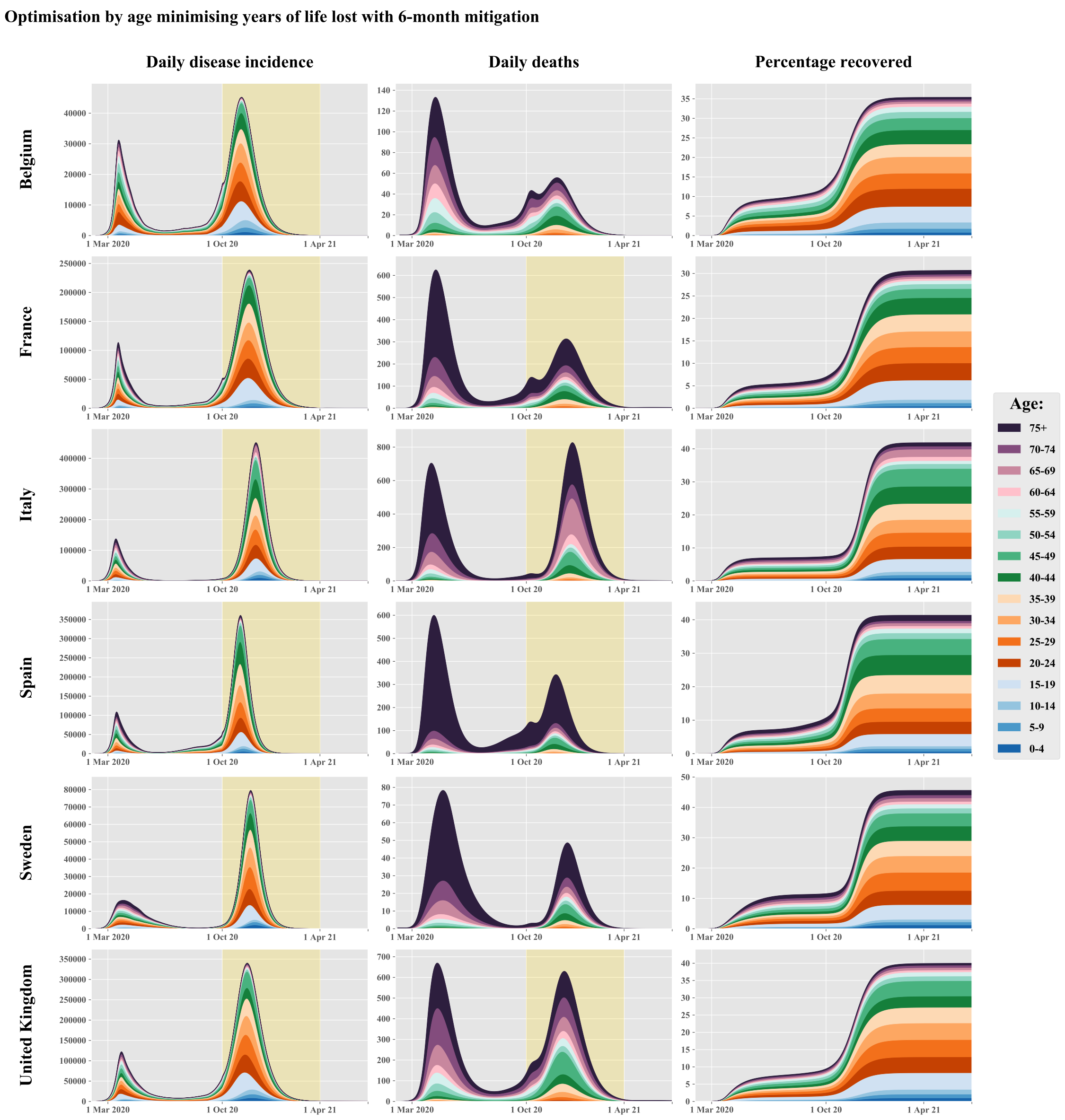

**Figure S24. Age-specific profile of disease incidence, COVID-19-related deaths and proportion recovered over time**
The yellow background indicates the intervention phase during which age-specific contacts were optimised. These projections were produced assuming that recovered individuals have persistent immunity against SARS-CoV-2 reinfection and using the maximum a posteriori estimates.

Optimisation by age minimising deaths with 12-month mitigation

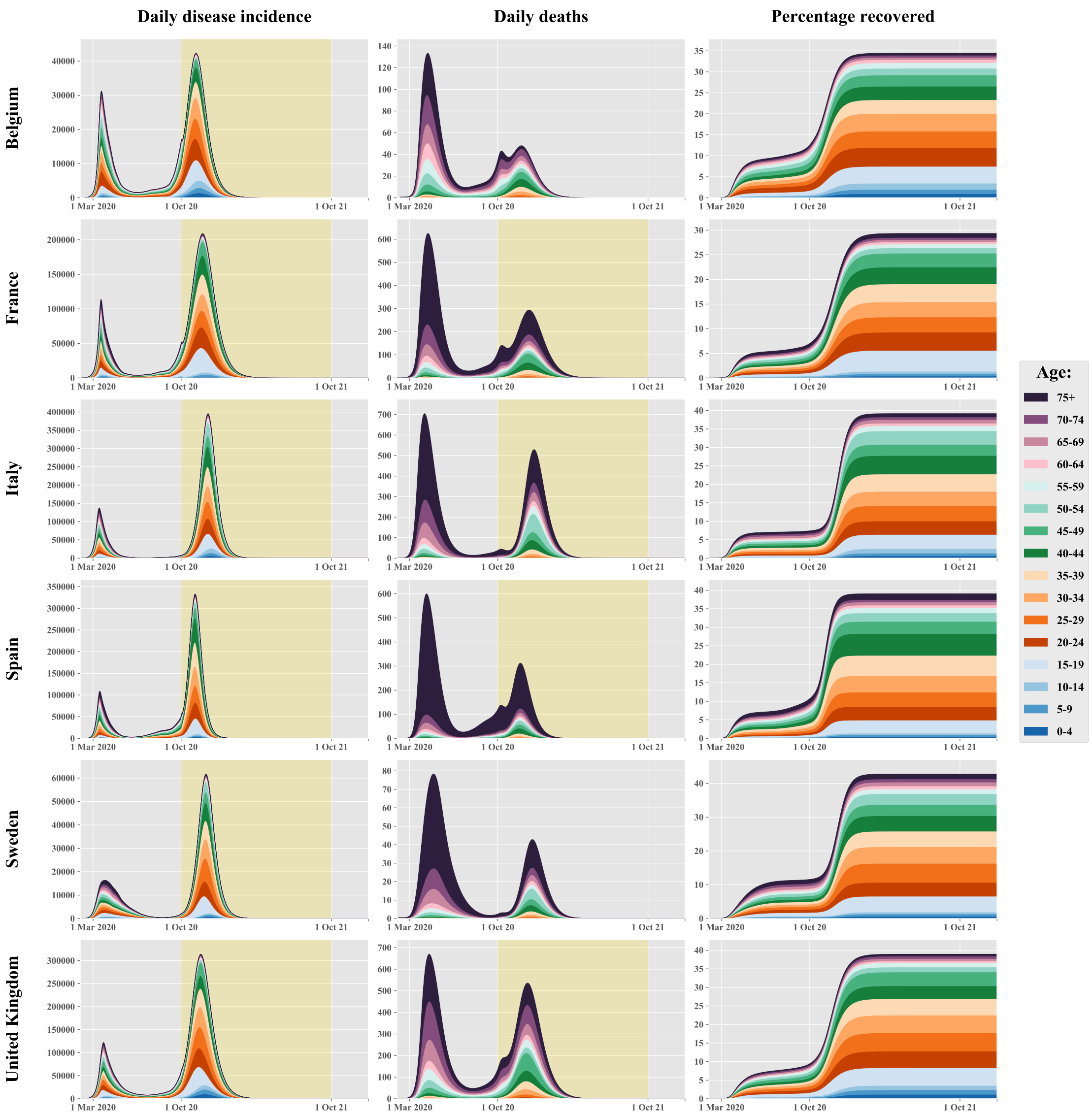

**Figure S25. Age-specific profile of disease incidence, COVID-19-related deaths and proportion recovered over time**

The yellow background indicates the intervention phase during which age-specific contacts were optimised. These projections were produced assuming that recovered individuals have persistent immunity against SARS-CoV-2 reinfection and using the maximum a posteriori estimates.

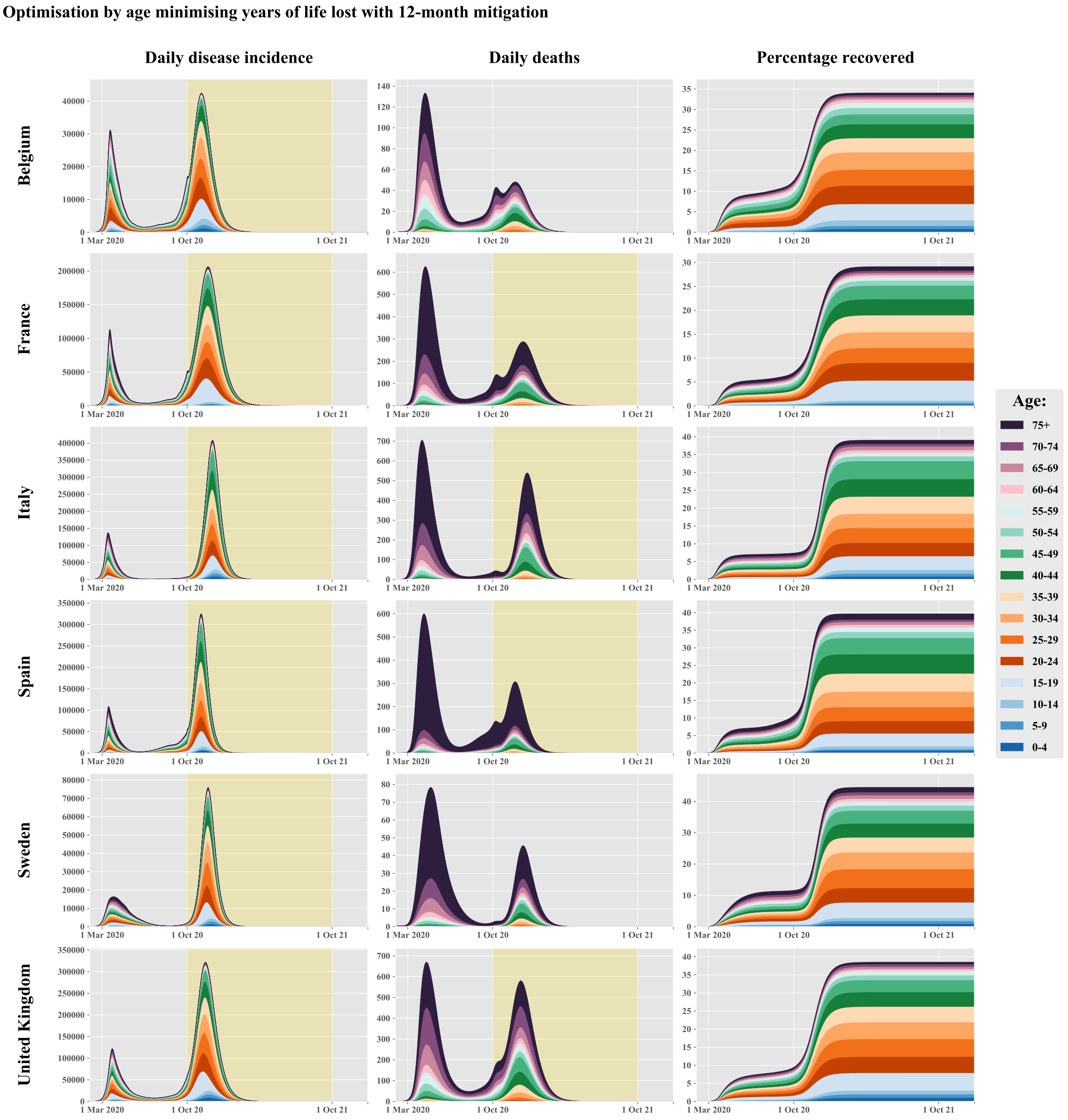

**Figure S26. Age-specific profile of disease incidence, COVID-19-related deaths and proportion recovered over time**

The yellow background indicates the intervention phase during which age-specific contacts were optimised. These projections were produced assuming that recovered individuals have persistent immunity against SARS-CoV-2 reinfection and using the maximum a posteriori estimates.

Optimisation by location minimising deaths with 6-month mitigation

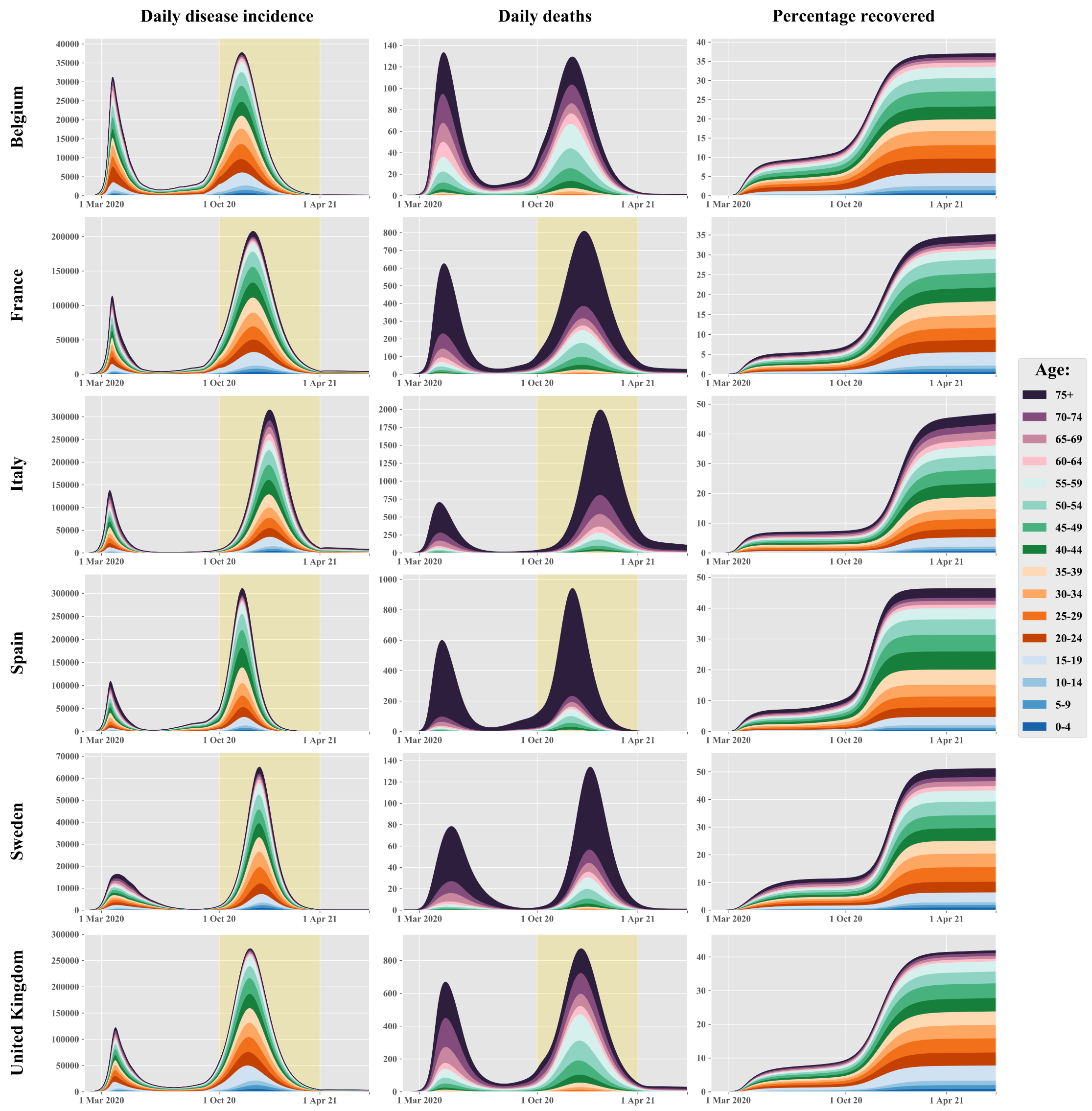

**Figure S27. Age-specific profile of disease incidence, COVID-19-related deaths and proportion recovered over time**

The yellow background indicates the intervention phase during which age-specific contacts were optimised. These projections were produced assuming that recovered individuals have persistent immunity against SARS-CoV-2 reinfection and using the maximum a posteriori estimates.

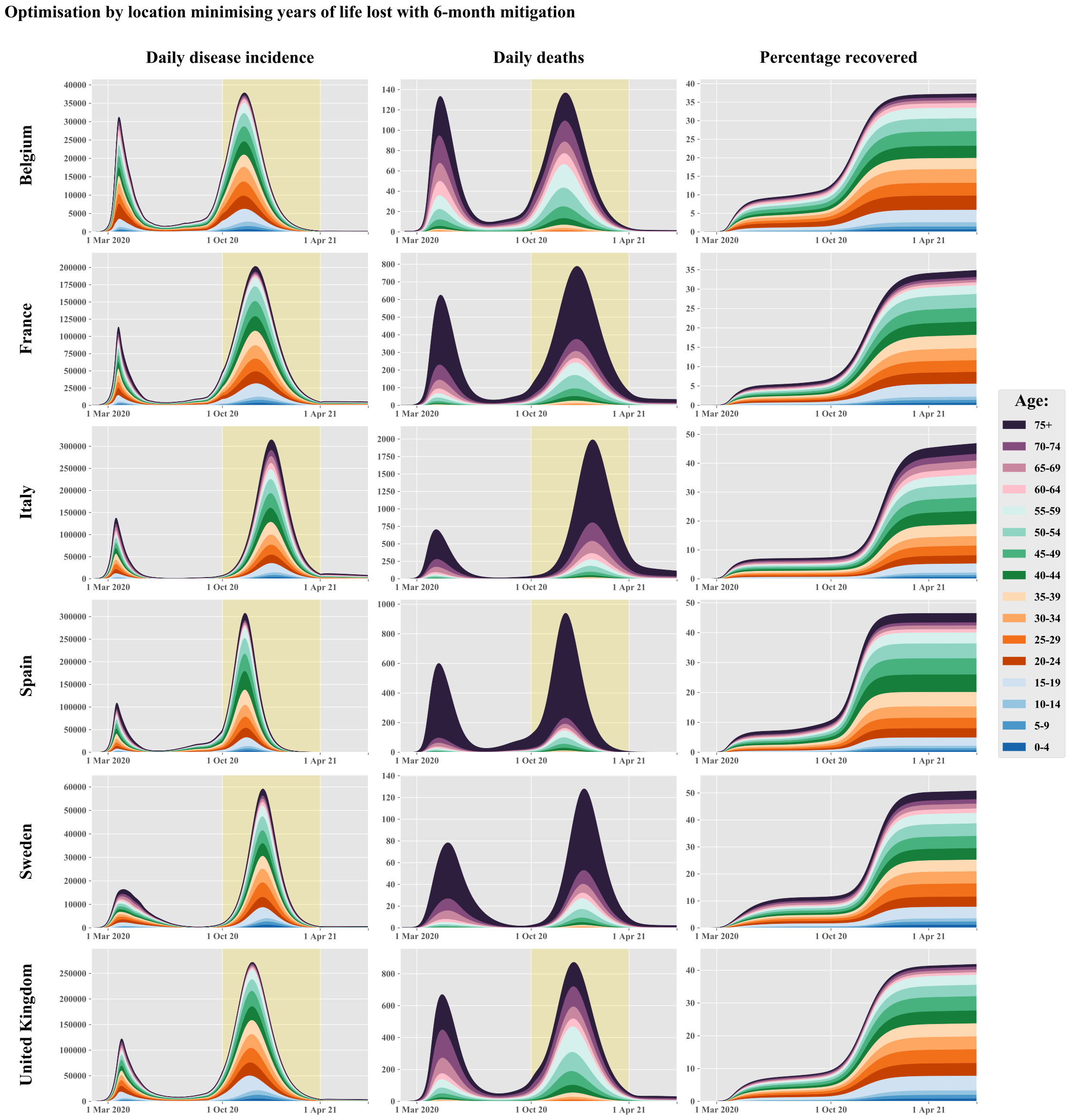

**Figure S28. Age-specific profile of disease incidence, COVID-19-related deaths and proportion recovered over time**

The yellow background indicates the intervention phase during which age-specific contacts were optimised. These projections were produced assuming that recovered individuals have persistent immunity against SARS-CoV-2 reinfection and using the maximum a posteriori estimates.

Optimisation by location minimising deaths with 12-month mitigation

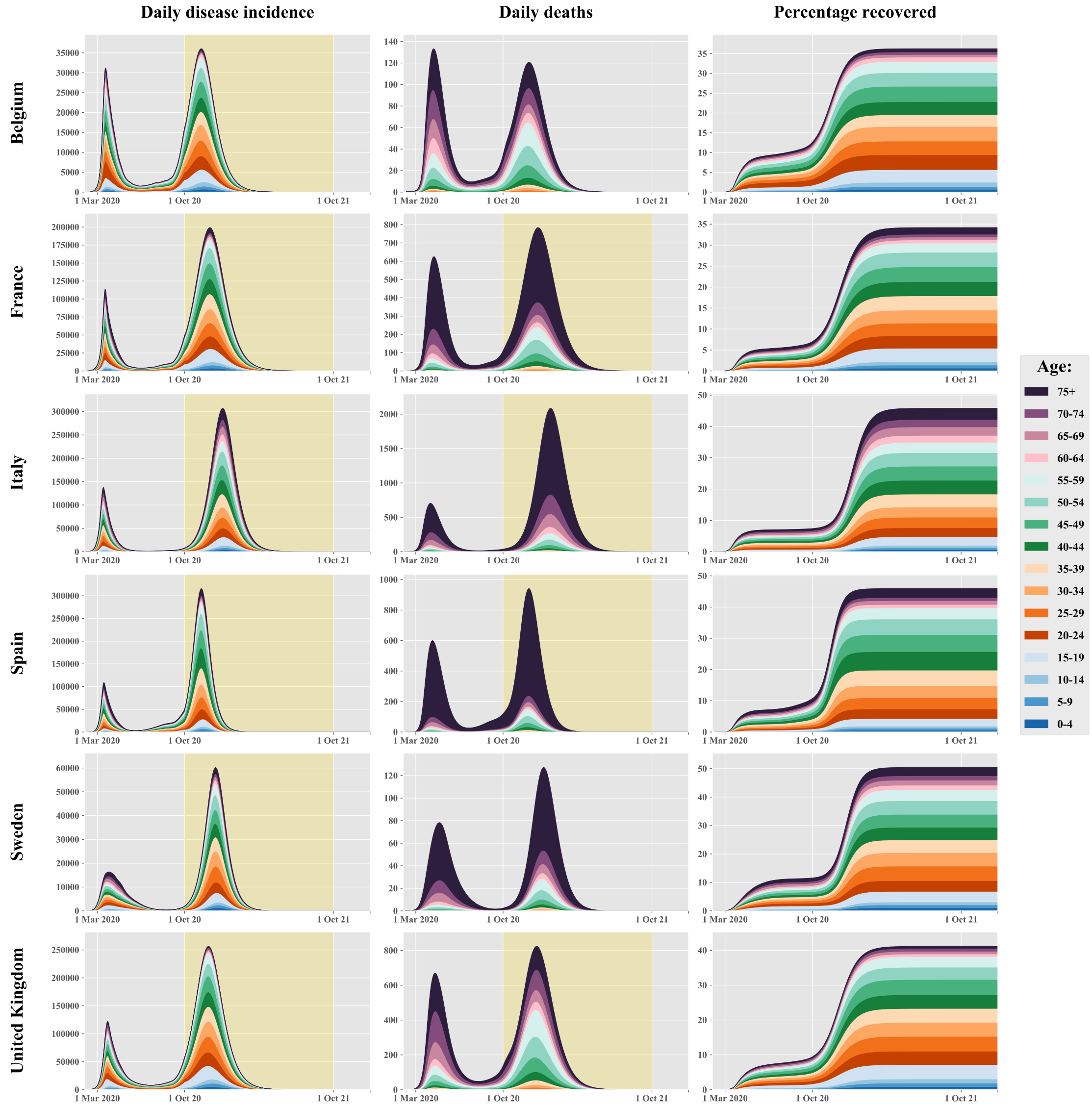

**Figure S29. Age-specific profile of disease incidence, COVID-19-related deaths and proportion recovered over time**

The yellow background indicates the intervention phase during which age-specific contacts were optimised. These projections were produced assuming that recovered individuals have persistent immunity against SARS-CoV-2 reinfection and using the maximum a posteriori estimates.

**Figure S30. Age-specific profile of disease incidence, COVID-19-related deaths and proportion recovered over time**

The yellow background indicates the intervention phase during which age-specific contacts were optimised. These projections were produced assuming that recovered individuals have persistent immunity against SARS-CoV-2 reinfection and using the maximum a posteriori estimates.

##### 5.8 Epidemic trajectory using different assumptions for the profile of waning immunity.

The optimisation searches were all performed based on the assumption of persistent immunity for recovered individuals. However, we also ran epidemic simulations considering various assumptions of waning immunity while using the optimal mitigation plans obtained when assuming full immunity. We considered 5 different scenarios regarding post-infection immunity:

1. immunity against reinfection for an average of 6 months
2. immunity against reinfection for an average of 6 months and 50% reduction in disease severity
3. immunity against reinfection for an average of 24 months
4. immunity against reinfection for an average of 24 months and 50% reduction in disease severity
5. persistent immunity against reinfection

The 50% reduction in disease severity was modelled as a 50% reduction in the probability of presenting symptoms during repeat SARS-CoV-2 infections. The structure of the model implied that the same reduction also applies to the risk of hospitalisation and death. Figures S31, S32, S33 and Figure 7 (main text) show the predicted COVID-19 incidence, mortality and hospital occupancy over time for the different assumptions and considering the optimal plan obtained using different optimisation configurations.

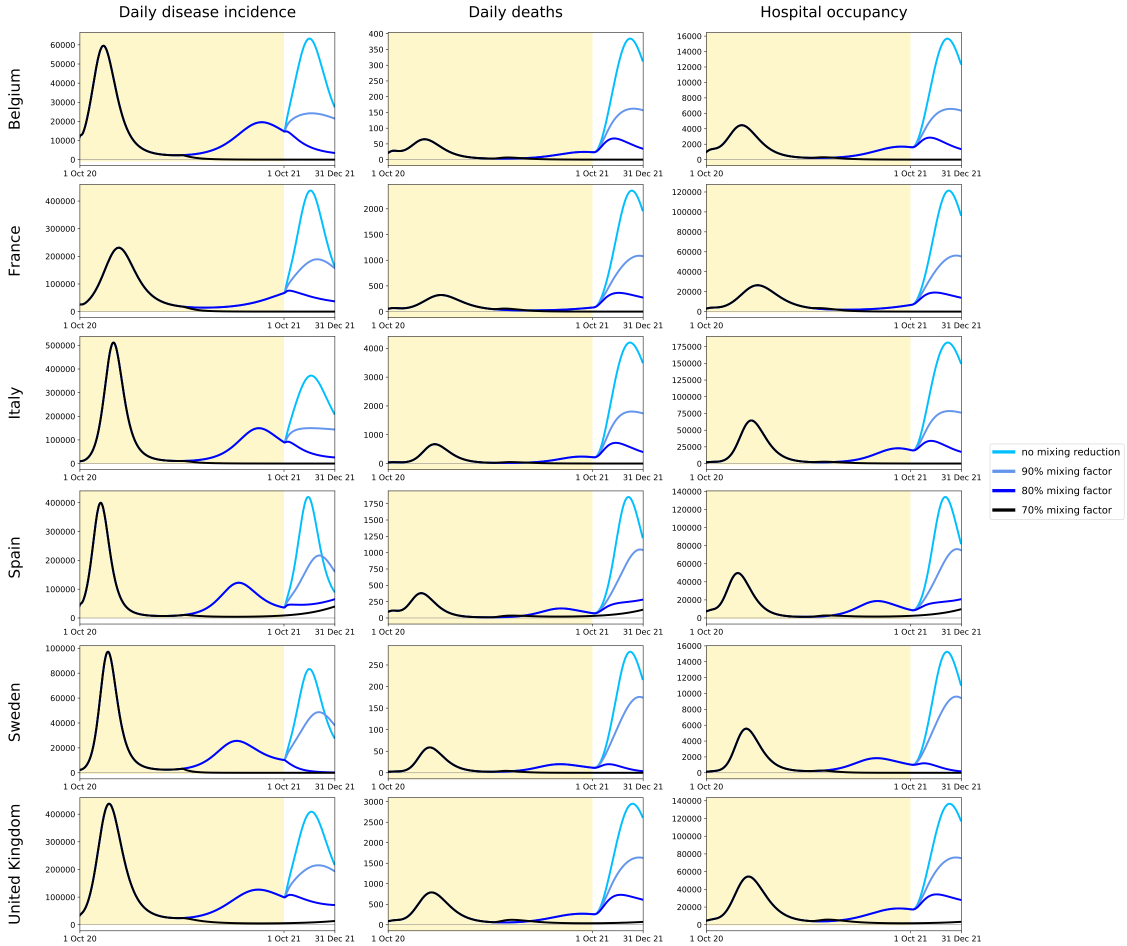

**Figure S31. Predicted COVID-19 incidence, mortality and hospital occupancy over time under various assumptions of waning immunity.**
The predictions were obtained using the maximum-likelihood parameter estimates and based on the 12-month contact mitigation by age minimising years of life lost (YLLs). The yellow background indicates the mitigation phase during which age-specific contacts were optimised. Five different assumptions were used to project the disease indicators: persistent immunity (black), 24-month immunity with and without 50% reduction in risk of symptoms for repeat infections (red and coral, respectively), 6-month immunity with and without 50% reduction in risk of symptoms for repeat infections (blue and turquoise, respectively).

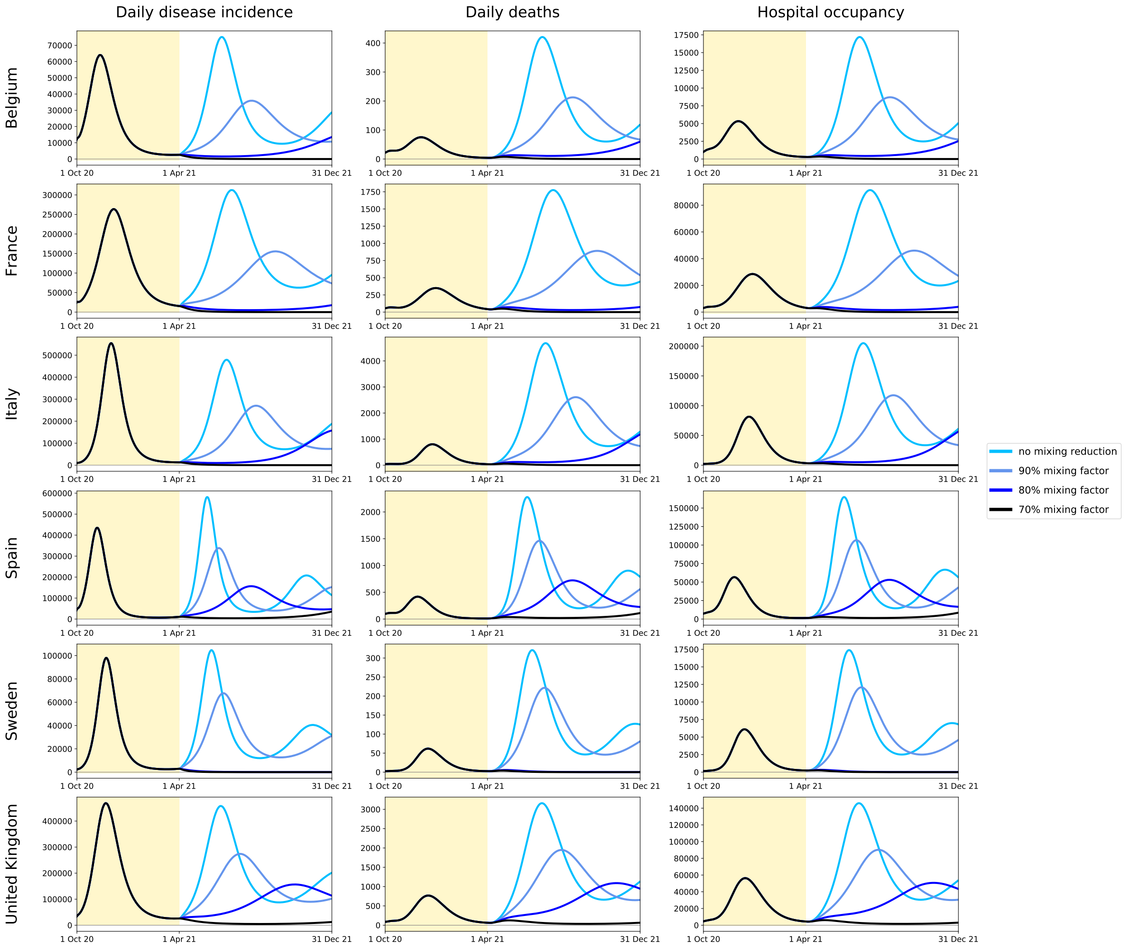

**Figure S32. Predicted COVID-19 incidence, mortality and hospital occupancy over time under various assumptions of waning immunity.**
The predictions were obtained using the maximum-likelihood parameter estimates and based on the 6-month contact mitigation by age minimising deaths. The yellow background indicates the mitigation phase during which age-specific contacts were optimised. Five different assumptions were used to project the disease indicators: persistent immunity (black), 24-month immunity with and without 50% reduction in risk of symptoms for repeat infections (red and coral, respectively), 6-month immunity with and without 50% reduction in risk of symptoms for repeat infections (blue and turquoise, respectively).

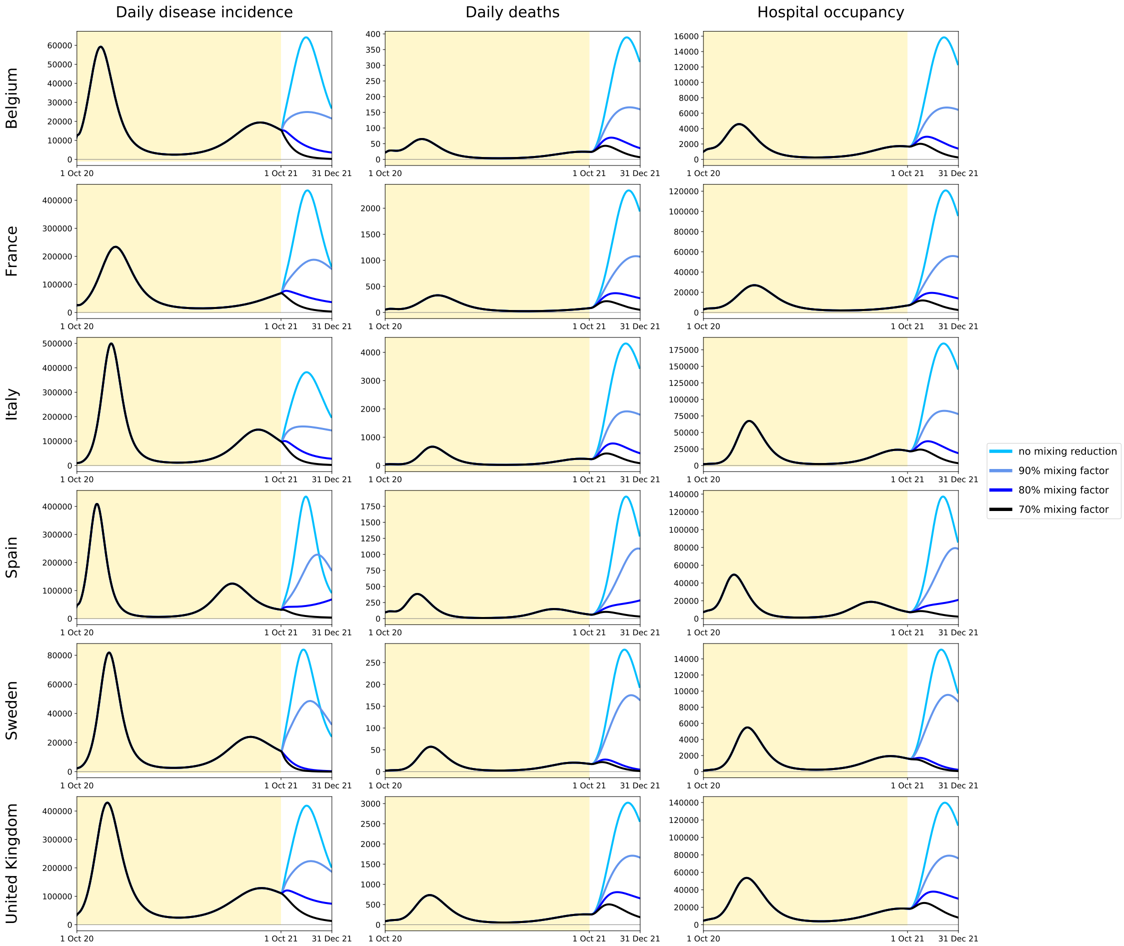

**Figure S33. Predicted COVID-19 incidence, mortality and hospital occupancy over time under various assumptions of waning immunity.**
The predictions were obtained using the maximum-likelihood parameter estimates and based on the 12-month contact mitigation by age minimising deaths. The yellow background indicates the mitigation phase during which age-specific contacts were optimised. Five different assumptions were used to project the disease indicators: persistent immunity (black), 24-month immunity with and without 50% reduction in risk of symptoms for repeat infections (red and coral, respectively), 6-month immunity with and without 50% reduction in risk of symptoms for repeat infections (blue and turquoise, respectively).

#### 5.9 Epidemic trajectory under short-lived immunity applying mild mitigation after optimised phase.

We ran simulations under our most pessimistic assumption regarding waning immunity (6 month average duration and no effect on repeat disease severity), considering that mild contact mitigation was applied after the optimised phase. Figure 8 (main text) presents the predictions associated with the optimal age-mitigation plan obtained by minimising YLLs over a period of 6 months. The results of the other optimisation configurations are presented below.

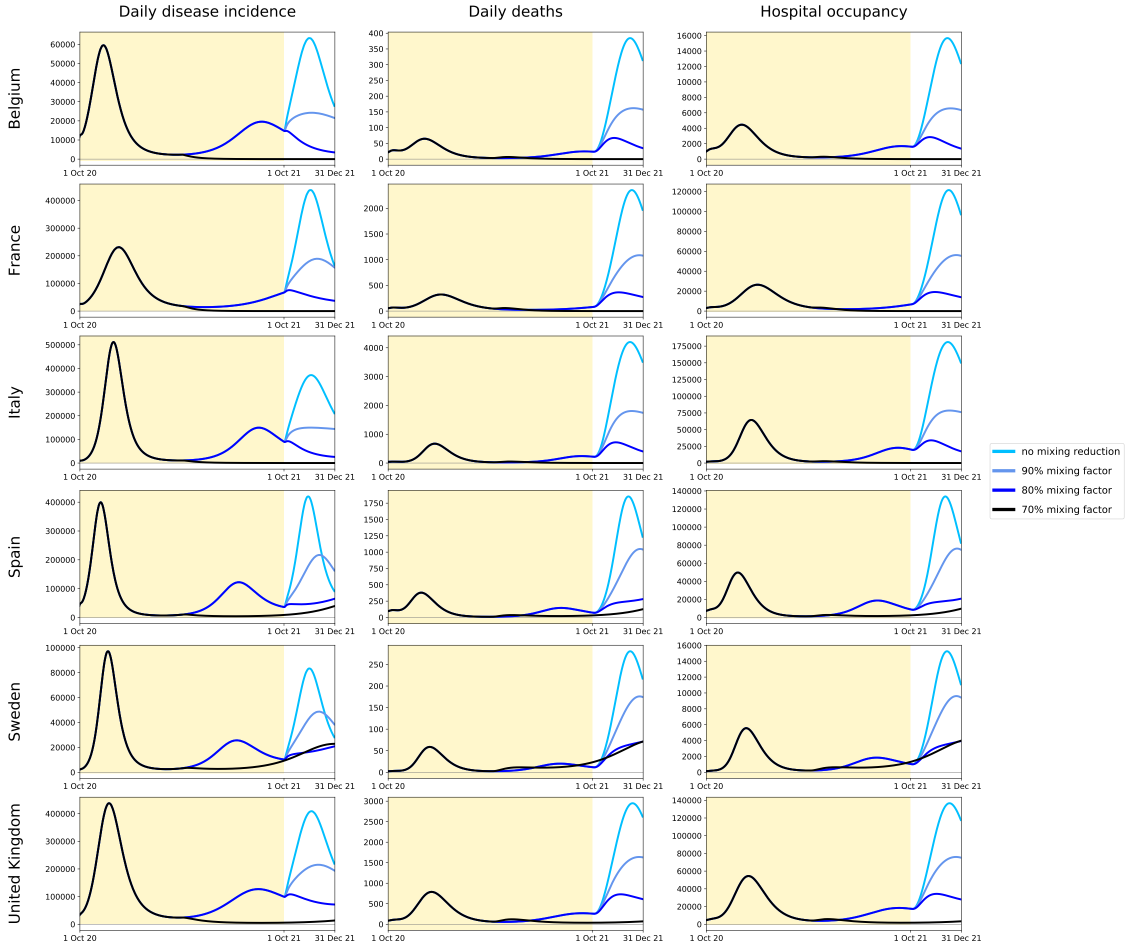

**Figure S34. Predicted COVID-19 incidence, mortality and hospital occupancy over time with short-lived post-infection immunity and applying mild mixing reductions after the optimised phase.**
The predictions were obtained using the maximum-likelihood parameter estimates and based on the 12-month contact mitigation by age minimising years of life lost (YLLs). The yellow background indicates the mitigation phase during which age-specific contacts were optimised. These predictions were obtained assuming 6-month average duration of immunity with no effect on the severity of repeat SARS-CoV-2 infections. The mixing factors were defined in the same way as during optimisation except that the same factor was applied to all age-groups. That is, a 90% mixing factor corresponds to a situation where every individual reduces their opportunity of contact by 10%.

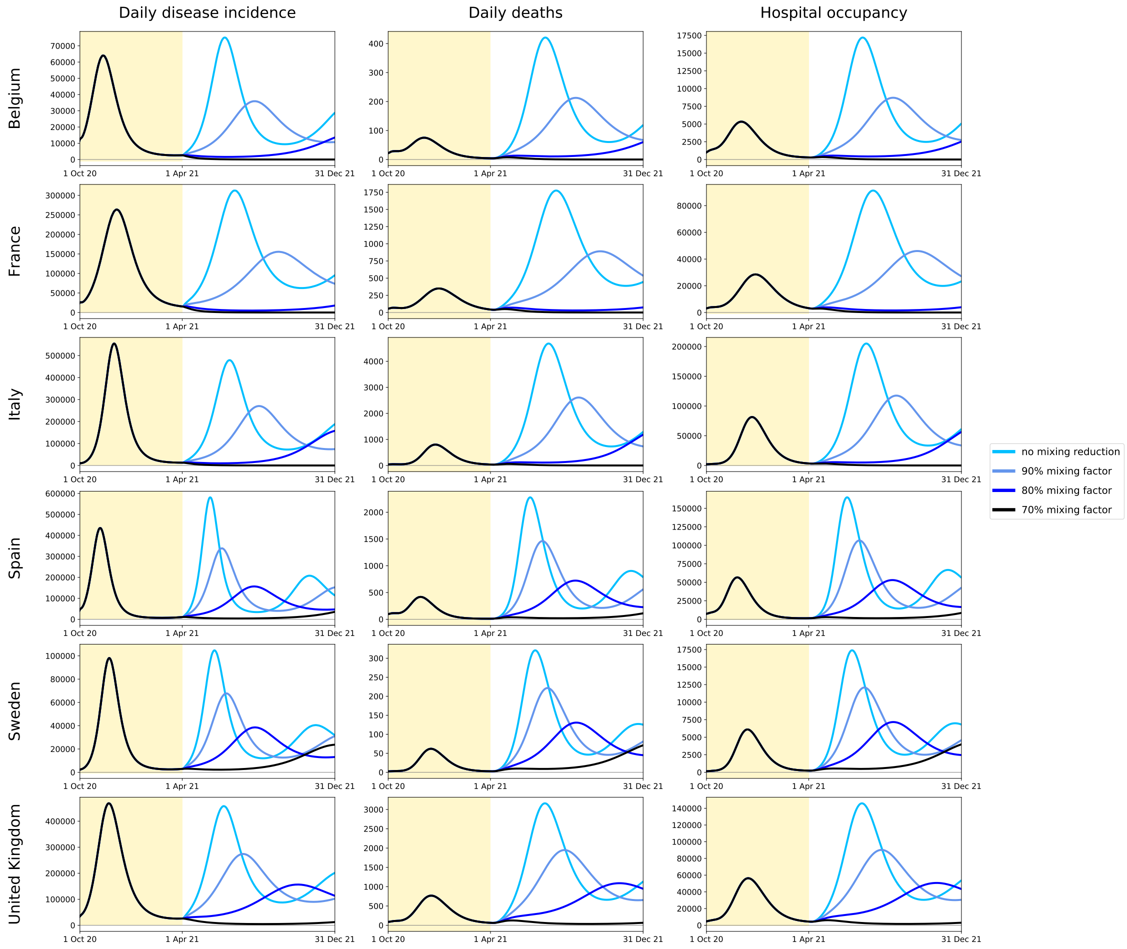

**Figure S35. Predicted COVID-19 incidence, mortality and hospital occupancy over time with short-lived post-infection immunity and applying mild mixing reductions after the optimised phase.**
The predictions were obtained using the maximum-likelihood parameter estimates and based on the 6-month contact mitigation by age minimising deaths. The yellow background indicates the mitigation phase during which age-specific contacts were optimised. These predictions were obtained assuming 6-month average duration of immunity with no effect on the severity of repeat SARS-CoV-2 infections. The mixing factors were defined in the same way as during optimisation except that the same factor was applied to all age-groups. That is, a 90% mixing factor corresponds to a situation where every individual reduces their opportunity of contact by 10%.

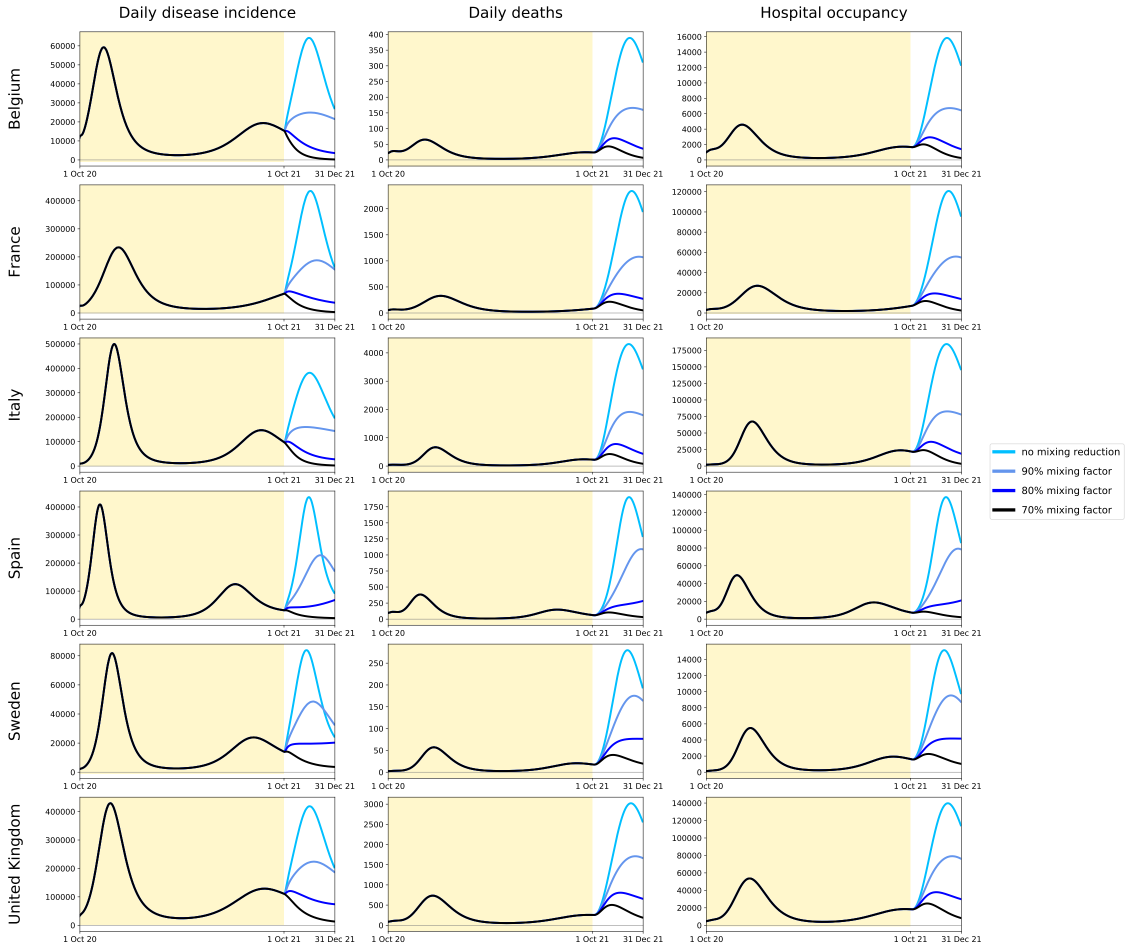

**Figure S36. Predicted COVID-19 incidence, mortality and hospital occupancy over time with short-lived post-infection immunity and applying mild mixing reductions after the optimised phase.**
The predictions were obtained using the maximum-likelihood parameter estimates and based on the 12-month contact mitigation by age minimising deaths. The yellow background indicates the mitigation phase during which age-specific contacts were optimised. These predictions were obtained assuming 6-month average duration of immunity with no effect on the severity of repeat SARS-CoV-2 infections. The mixing factors were defined in the same way as during optimisation except that the same factor was applied to all age-groups. That is, a 90% mixing factor corresponds to a situation where every individual reduces their opportunity of contact by 10%.

[6] UNESCO, "COVID-19 Impact on Education," 2020.

[7] Google, "COVID-19 Community Mobility Reports."

[8] J. Zhang, M. Litvinova, W. Wang, Y. Wang, X. Deng, X. Chen, M. Li, W. Zheng, L. Yi, X. Chen, Q. Wu, Y. Liang, X. Wang, J. Yang, K. Sun, I. M. Longini, M. E. Halloran, P. Wu, B. J. Cowling, S. Merler, C. Viboud, A. Vespignani, M. Ajelli, and H. Yu, "Evolving epidemiology and transmission dynamics of coronavirus disease 2019 outside Hubei province, China: a descriptive and modelling study," *The Lancet Infectious Diseases*, vol. 20, no. 7, 2020.

[9] S. A. Lauer, K. H. Grantz, Q. Bi, F. K. Jones, Q. Zheng, H. R. Meredith, A. S. Azman, N. G. Reich, and J. Lessler, "The Incubation Period of Coronavirus Disease 2019 (COVID-19) From Publicly Reported Confirmed Cases: Estimation and Application.," *Annals of Internal Medicine*, vol. 172, pp. 577–582, may 2020.

[10] Q. Li, X. Guan, P. Wu, X. Wang, L. Zhou, Y. Tong, R. Ren, K. S. Leung, E. H. Lau, J. Y. Wong, X. Xing, N. Xiang, Y. Wu, C. Li, Q. Chen, D. Li, T. Liu, J. Zhao, M. Liu, W. Tu, C. Chen, L. Jin, R. Yang, Q. Wang, S. Zhou, R. Wang, H. Liu, Y. Luo, Y. Liu, G. Shao, H. Li, Z. Tao, Y. Yang, Z. Deng, B. Liu, Z. Ma, Y. Zhang, G. Shi, T. T. Lam, J. T. Wu, G. F. Gao, B. J. Cowling, B. Yang, G. M. Leung, and Z. Feng, "Early transmission dynamics in Wuhan, China, of novel coronavirus-infected pneumonia," mar 2020.

[17] "Wekelijkse update epidemiologische situatie COVID-19 in Nederland — RIVM."

[18] H. Haario, E. Saksman, and J. Tamminen, "An adaptive Metropolis algorithm," *Bernoulli*, 2001.

[19] WHO, "WHO COVID-19 dashboard."

[20] Sciensano, "COVID-19 Belgium Epidemiological Situation."

[21] Santé Publique France, "Données hospitalières relatives à l'épidémie de COVID-19."

[22] Dipartimento della Protezione Civile, "Emergenza Coronavirus: la risposta nazionale."

[23] Ministerio de Sanidad (Spanish Ministry of Health), "Actualización nº115: enfermedad por SARS-CoV-2 (COVID-19) 24.05.2020."

[24] Svenska Intensivvårdsregistret (Swedish Intensive Care Registry), "Covid-19 på svenska intensivvårdsavdelningar."

[25] UK Government, "Coronavirus (COVID-19) in the UK."

[26] S. Herzog, J. D. Bie, S. Abrams, I. Wouters, E. Ekinci, L. Patteet, A. Coppens, S. D. Spiegeleer, P. Beutels, P. V. Damme, N. Hens, and H. Theeten, "Seroprevalence of IgG antibodies against SARS coronavirus 2 in Belgium: a prospective cross-sectional study of residual samples," *medRxiv*, 2020.

[27] S. L. Vu, G. Jones, F. Anna, T. Rose, J.-B. Richard, S. Bernard-Stoecklin, S. Goyard, C. Demeret, O. Helynck, C. Robin, V. Monnet, L. P. de Facci, M.-N. Ungeheuer, L. Léon, Y. Guillois, L. Filleul, P. Charneau, D. Lévy-Bruhl, S. van der Werf, and H. Noel, "Prevalence of SARS-CoV-2 antibodies in France: results from nationwide serological surveillance," *medRxiv*, p. 2020.10.20.20213116, jan 2020.

[28] M. Pollán, B. Pérez-Gómez, R. Pastor-Barriuso, J. Oteo, M. A. Hernán, M. Pérez-Olmeda, J. L. Sanmartín, A. Fernández-García, I. Cruz, N. Fernández de Larrea, M. Molina, F. Rodríguez-Cabrera, M. Martín, P. Merino-Amador, J. León Paniagua, J. F. Muñoz-Montalvo, F. Blanco, R. Yotti, R. Gutiérrez Fernández, S. Mezcua Navarro, J. F. Muñoz-Montalvo, M. Salinero Hernández, J. L. Sanmartín, M. Cuenca-Estrella, J. León Paniagua, P. Fernández-Navarro, A. Avellón, G. Fedele, J. Oteo Iglesias, M. T. Pérez Olmeda, M. E. Fernandez Martinez, F. D. Rodríguez-Cabrera, M. A. Hernán, S. Padrones Fernández, J. M. Rumbao Aguirre, J. M. Navarro Marí, B. Palop Borrás, A. B. Pérez Jiménez, M. Rodríguez-Iglesias, A. M. Calvo Gascón, M. L. Lou Alcaine, I. Donate Suárez, O. Suárez Álvarez, M. Rodríguez Pérez, M. Cases Sanchís, C. J. Villafáfila Gomila, L. Carbo Saladrigas, A. Hurtado Fernández, A. Oliver, E. Castro Feliciano, M. N. González Quintana, J. M. Barrasa Fernández, M. A. Hernández Betancor, M. Hernández Febles, L. Martín Martín, L. M. López López, T. Ugarte Miota, I. De Benito Población, M. S. Celada Pérez, M. N. Vallés Fernández, T. Maté Enríquez, M. Villa Arranz, M. Domínguez-Gil González, I. Fernández-Natal, G. Megías Lobón, J. L. Muñoz Bellido, P. Ciruela, A. Mas i Casals, M. Doladé Botías, M. A. Marcos Maeso, D. Pérez del Campo, A. Félix de Castro, R. Limón Ramírez, M. F. Elías Retamosa, M. Rubio González, M. S. Blanco Lobeiras, A. Fuentes Losada, A. Aguilera, G. Bou, Y. Caro, N. Marauri, L. M. Soria Blanco, I. del Cura González, M. Hernández Pascual, R. Alonso Fernández, N. Cabrera Castro, A. Tomás Lizcano, C. Ramírez Almagro, M. Segovia Hernández, N. Ascunce Elizaga, M. Ederra Sanz, C. Ezpeleta Baquedano, A. Bustinduy Bascaran, S. Iglesias Tamayo, L. Elorduy Otazua, R. Benarroch Benarroch, J. Lopera Flores, and A. Vázquez de la Villa, "Prevalence of SARS-CoV-2 in Spain (ENE-COVID): a nationwide, population-based seroepidemiological study," *The Lancet*, 2020.

[29] T. S. P. H. Agency, "Detection of antibodies after the covid-19 review in blood samples from outpatient care (Sub-report 1)," tech. rep., 2020.

[30] Biobank, "UK Biobank SARS-CoV-2 Serology Study," tech. rep., 2020.

[31] E. G. Talbi, *Metaheuristics: From Design to Implementation.* Wiley Series on Parallel and Distributed Computing, Wiley, 2009.

[32] G. Briffoteaux, R. Ragonnet, M. Mezmaz, N. Melab, and D. Tuyttens, "Evolution control for parallel ann-assisted simulation-based optimization application to tuberculosis transmission control," *Future Generation Computer Systems*, vol. 113, pp. 454 – 467, 2020.

[33] R. Bolze *et al.*, "Grid'5000: A large scale and highly reconfigurable experimental grid testbed," *The International Journal of High Performance Computing Applications*, vol. 20, no. 4, pp. 481–494, 2006.

[34] H. Ward, C. Atchison, M. Whitaker, K. Ainslie, J. Elliott, L. Okell, R. Redd, D. Ashby, C. Donnelly, W. Barclay, A. Darzi, G. Cooke, S. Riley, and P. Elliott, "Antibody prevalence for SARS-CoV-2 following the peak of the pandemic in England: REACT2 study in 100,000 adults," *Preprint*, 2020.
